# Barriers and Facilitators for the Use of Telehealth by Healthcare Providers (HCP) in India - A Scoping Review

**DOI:** 10.1101/2023.10.28.23297653

**Authors:** Parth Sharma, Shirish Rao, Padmavathy Krishna Kumar, Aiswarya R Nair, Disha Agrawal, Siddhesh Zadey, S Gayathri, Rachna George Joseph, Girish Dayma, Liya Rafeekh, Shubhashis Saha, Sitanshi Sharma, SS Prakash, Venkatesan Sankarapandian, Preethi John, Vikram Patel

**Affiliations:** Association for Socially Applicable Research (ASAR), Pune, Maharashtra, India; Department of Community Medicine, Maulana Azad Medical College, Delhi, India; Seth GS Medical College and KEM Hospital, Mumbai, Maharashtra, India; Adichunchanagiri Institute of Medical Sciences, BG Nagara, Karnataka, India; Travancore Medical College, Kollam, Kerala, India; Dr D. Y. Patil Medical College, Hospital, and Research Centre Pune, Dr. D. Y. Patil Vidyapeeth, Pune, Maharashtra, India; Global Emergency Medicine Innovation and Implementation (GEMINI) Research Center, Duke University School of Medicine, Durham North Carolina USA; Department of Epidemiology, Columbia University Mailman School of Public Health, New York City New York USA; Christian Medical College and Hospital, Vellore, Tamil Nadu, India; KEM Hospital Research Centre, Pune, Maharashtra, India; Indian Institute of Technology, Kharagpur, West Bengal, India; Centre for Health Research and Development, Society for applied studies, Delhi, India; Global Business School for Health, University College London UK; Harvard T.H. Chan School of Public Health, Boston MA USA

**Keywords:** Digital health, Telehealth, mHealth, Human resources for health

## Abstract

**Background:** It is widely assumed that telehealth tools like mHealth, telemedicine, and tele-education can supplement the efficiency of Healthcare Providers (HCPs). We conducted a scoping review of evidence on the barriers and facilitators associated with the use of telehealth by HCPs in India.

**Methods:** A systematic literature search following a pre-registered protocol (https://doi.org/10.17605/OSF.IO/KQ3U9 [PROTOCOL DOI]) was conducted in PubMed. The search strategy, inclusion, and exclusion criteria were based on the World Health Organization’s action framework on Human Resources for Health (HRH) and Universal Health Coverage in India with a specific focus on telehealth tools. Eligible articles published in English from 1st January 2001 to 17th February 2022 were included.

**Results:** One hundred and six studies were included in the review. Of these, 53 studies (50%) involved mHealth interventions, 25 (23.6%) involved telemedicine interventions whereas the remaining 28 (26.4%) involved the use of tele-education interventions by HCPs in India. In each category, most of the studies followed a quantitative study design and were mostly published in the last 5 years. The study sites were more commonly present in states present in south India. The facilitators and barriers related to each type of intervention were analyzed under the following sub-headings-1) Human resource related, 2) Application related 3) Technical, and 4) Others. The interventions were most commonly used for improving the management of mental health, non-communicable diseases, and maternal and child health.

**Conclusions:** Use of telehealth has not been uniformly studied in India. The facilitators and barriers to telehealth use need to be kept in mind while designing the intervention. Future studies should focus on looking at region-specific, intervention-specific, and health cadre-specific barriers and facilitators for the use of telehealth.

## Introduction

Telehealth is defined “as the delivery and facilitation of health and health-related services including medical care, provider and patient education, health information services, and self-care via telecommunications and digital communication technologies.”(1) Even though used interchangeably, telehealth and telemedicine are not the same. Telehealth covers a wide range of services like telemedicine, mHealth, and remote patient monitoring. Telemedicine refers to the delivery of diagnostic or treatment services to a patient using telecommunication technology remotely. (1) mHealth on the other hand refers to applications or programs used on smartphones or tablets. (1) These interventions could be used to address the shortage of human resources for health (HRH), for education and training of HCPs, or for supporting the functioning of the existing health workforce.

Access to healthcare of adequate quality is inequitable in India which disproportionately affects(2) rural and low-resourced states, where a majority of the Indian population resides. (3) Access is worse for those belonging to vulnerable groups like the elderly, and people with disability. (4) A major driver for this inequitable access is the inequitable distribution of human resources for health (HRH). (5) These barriers have resulted in the rapid privatization of healthcare in India,(6) thus making healthcare a leading cause of out-of-pocket expenditure. (7) It is widely assumed that inequitable access to quality care could be addressed by telehealth interventions like mHealth and telemedicine and also help in cutting the costs of healthcare. (8–10)

To enhance the uptake of digital health interventions, the World Health Organization (WHO) published its Global Strategy on Digital Health for 2020-2025. (11) In India, the National Health Policy (NHP) 2017 recommended the use of Information and Communications Technologies (ICT) to improve access to health services. In recent years, there has been a mushrooming of a range of telehealth interventions in India, for example, mSakhi and ASHA Kirana(12,13) in antenatal and postnatal maternal care through patient monitoring and behavior change communication; for the care of people with non-communicable diseases(14); the eSanjeevani telemedicine portal to improve access to care in remote areas(15), and to train HRH. (16)

In the context of the significant place occupied by telehealth in national health policy and the multitude of telehealth interventions being piloted in different sectors and regions of the country, this review was conducted with the primary objective of understanding the facilitators, and barriers associated with the use of telehealth tools, like telemedicine, tele-education and mHealth, by HR in India. We also aimed to look at the role of telehealth in various aspects of the health system from service delivery, education, and training of HRHs, to its impact on their functioning and also their attitude toward the intervention.

## Methods

### Overview

This study is a scoping review conducted as one of the components of a larger evidence synthesis exercise undertaken by the Lancet Citizens’ Commission on Reimagining India’s Health System (www.citizenshealth.in). The protocol for evidence synthesis for the entire HRH workstream was registered on 16th June 2022. (17) It is in compliance with the Arksey and O’Malley methodological framework(18) and the PRISMA-ScR guidelines (19) and can be accessed here-https://doi.org/10.17605/OSF.IO/KQ3U9 [PROTOCOL DOI].

### Search Strategy

The review was a part of the larger evidence synthesis work on HRH for UHC in India. The search was conducted for published literature between 1st January 2001 and 17th February 2022 in the PubMed database. The search strategy focused on the WHO action framework on HRH and diverse categories of medical professional cadres along with universal health care in India **(S1 Panel)**.

### Screening and Selection

All the articles identified through the search strategy using the above-mentioned database were added to the Distiller SR software and duplicates were removed. A multi-level screening of articles using DistillerSR software was carried out by the team as described in the PRISMA-ScR 2020 diagram (**Fig 1**). Inclusion criteria included studies conducted in India and reported in English that focused on the use of telehealth by healthcare providers. Studies only evaluating clinical outcomes but not related to HRH cadre or management strategies or practices and study protocols, editorials, viewpoints, commentaries, letters, and correspondences were excluded.

**Fig 1:**
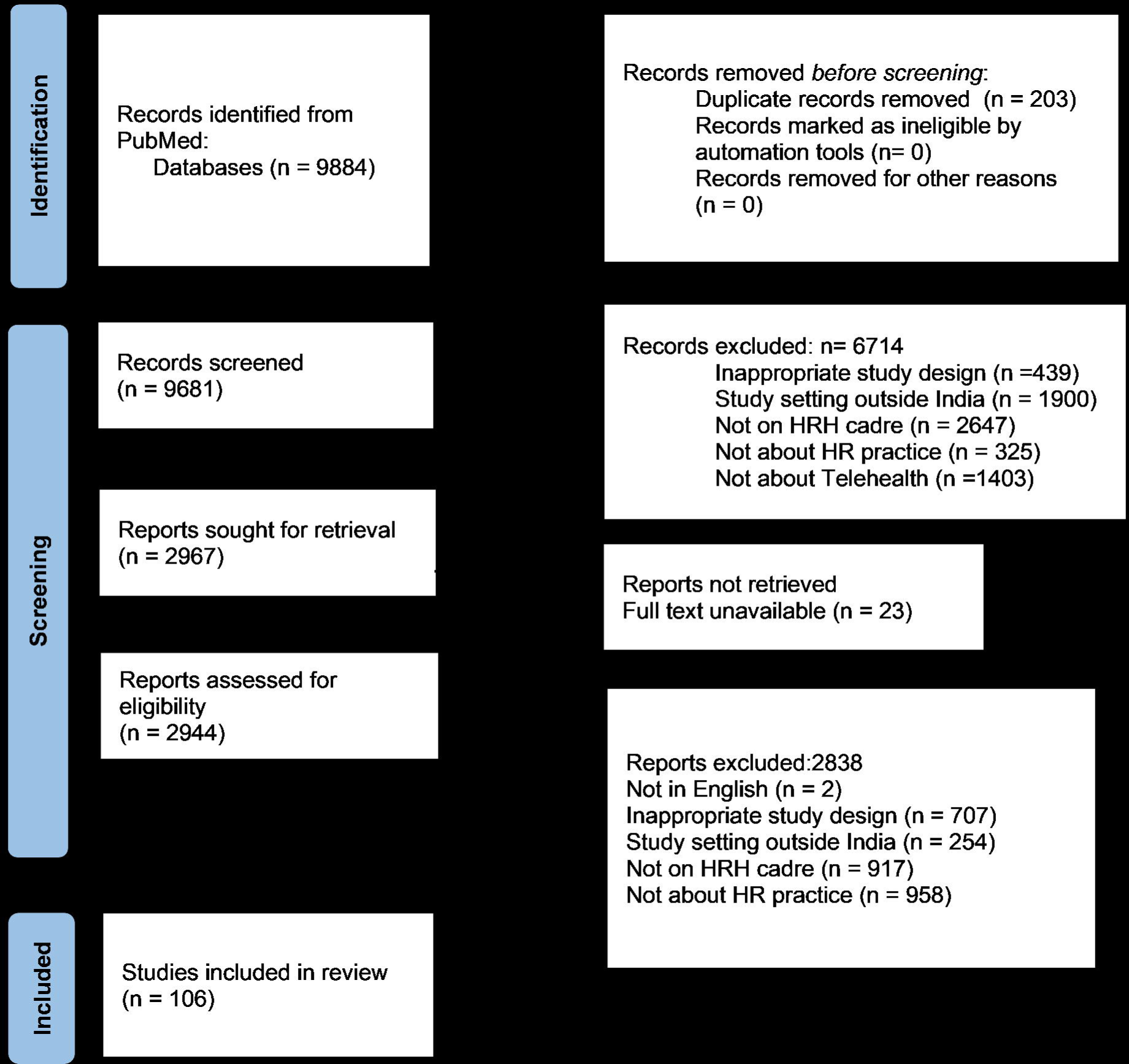
PRISMA flowchart showing selection and inclusion of the studies in the review.

The articles were divided into a team of two reviewers. At Level 1, the articles were screened only based on the title and abstract. The articles included by any one reviewer at Level 1 screening were moved to Level 2. The full text of all the articles in Level 2 was reviewed independently by two reviewers. After the full-text screening, articles were finally excluded or included only if both reviewers were in agreement. Conflicts about the eligibility criteria were resolved either through consensus between the two reviewers or by consulting one more reviewer.

### Data extraction and analysis

At Level 3, data charting for all the included full-text articles was done. Charting done by one author was verified by the other author. The data extracted included the following variables: Authors, Year of Publication, Study Design, Study Setting, Study Location, HR cadre, HR practice, Sample Size, Primary Objectives, Primary Outcomes, Impact, Challenges and barriers, and Study limitations. Articles were then classified based on the type of telehealth intervention into mHealth, telemedicine, and tele-education. A descriptive analysis of the included articles was conducted to understand barriers and facilitators of telehealth use.

## Results

One hundred and six studies were included in the review. Of these, 53 studies (50%) involved mHealth interventions (13,20–71), 25 (23.6%) involved telemedicine interventions (72–96) whereas the remaining 28 (26.4%) involved the use of tele-education interventions by HCPs in India. (97–124)

### mHealth

Of the total 53 studies, nearly half the studies (45%) were quantitative(21,26–28,31,33– 36,38,39,42,43,45,50,51,53,55,58,59,63,64,66,70), 14 (26%) were qualitative (13,20,22,29,32,37,44,46–48,60,65,67,69), 12 (23%) were mixed-methods (23–25,30,40,41,52,54,56,57,62,68) and 3 (6%) were review studies.(49,61,71) No study on the use of mHealth was published before 2013 and 64% (13,20–51,55) of the studies were published after 2018 with the maximum (n=12, 23%)(13,22,24–31,33,34) number of studies being published in 2021. The studies were conducted in tertiary care or teaching hospital settings (n=28)(22,23,26–29,31–38,46,47,49,55–59,61,63,65,68,69,71), community health centers (n=6)(20,21,42,44,60,66), primary health centers (n=16)(13,30,40,43,48,50–53),(39,54,62,64,66,67,70), and other settings (n=4). (24,25,41,45) The use of mHealth was most studied in Karnataka (n=7)(13,28,41,52,66,67,71), followed by Gujarat (21,38,44,58,65), Maharashtra (26,28,32,49,66), and Tamil Nadu (26,36,37,51,53) (5 studies each) (**Fig 2**). Findings from all included studies have been summarized in the S1 Table.

**Fig 2:**
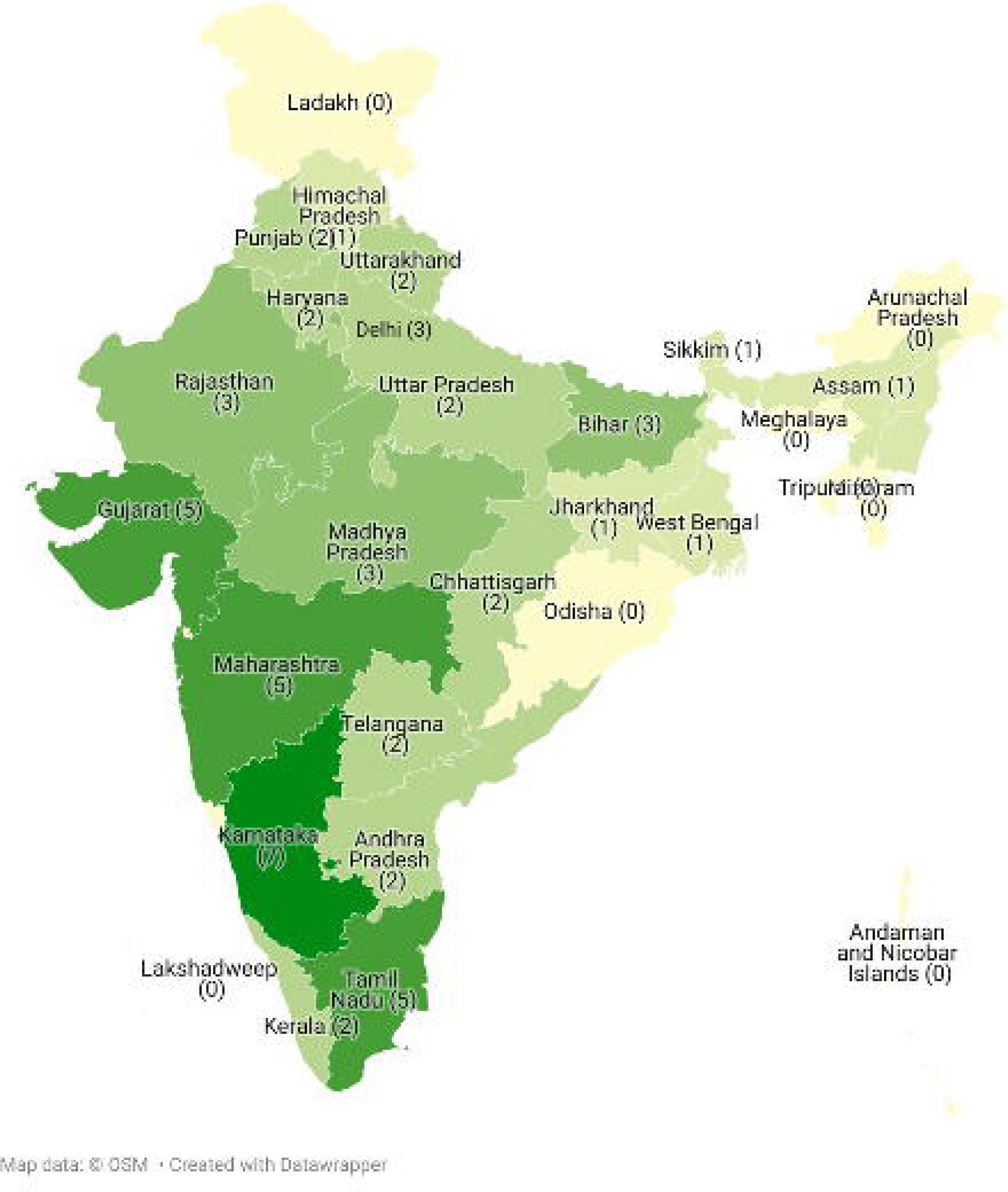
The number of study sites per state in India for mHealth.

mHealth interventions were most commonly used by doctors (n=38)(21–23,25–27,30,31,35– 38,40–45,47–50,52–54,56–62,64,65,68–71), followed by community healthcare workers (n=18)(22,28–30,32–34,36,39,41,44,46,49,51,58,63,66,67), nurses (n=8)(13,20,22,41,42,55,56,68), allied health professionals (n=4)(43,54,68,70), auxiliary midwife nurses (n=3)(22,36,41), and others (n=8). (24,29,36,46,54,62,64,68)

#### 1. Facilitators and barriers to the use of mHealth

Prior training to use the mHealth intervention (n=19)(13,20,23,25,26,32,37,41,42,44,46,49,50,52,55,57,60,62,63), interactive intervention with the use of videos and images (n=14)(21,23,34,39–41,44–46,49,60,67–69), and availability of the device to use the intervention (n=6)(22,23,48,49,58,61) were the most common human resource-related, application-related, and technical facilitators respectively. Formative research prior to designing the intervention (n=4)(29,32,44,48) and government support for the intervention (n=2)(29,30) were other facilitators that were identified. Other facilitators are mentioned in **Fig 3**.

**Fig 3:**
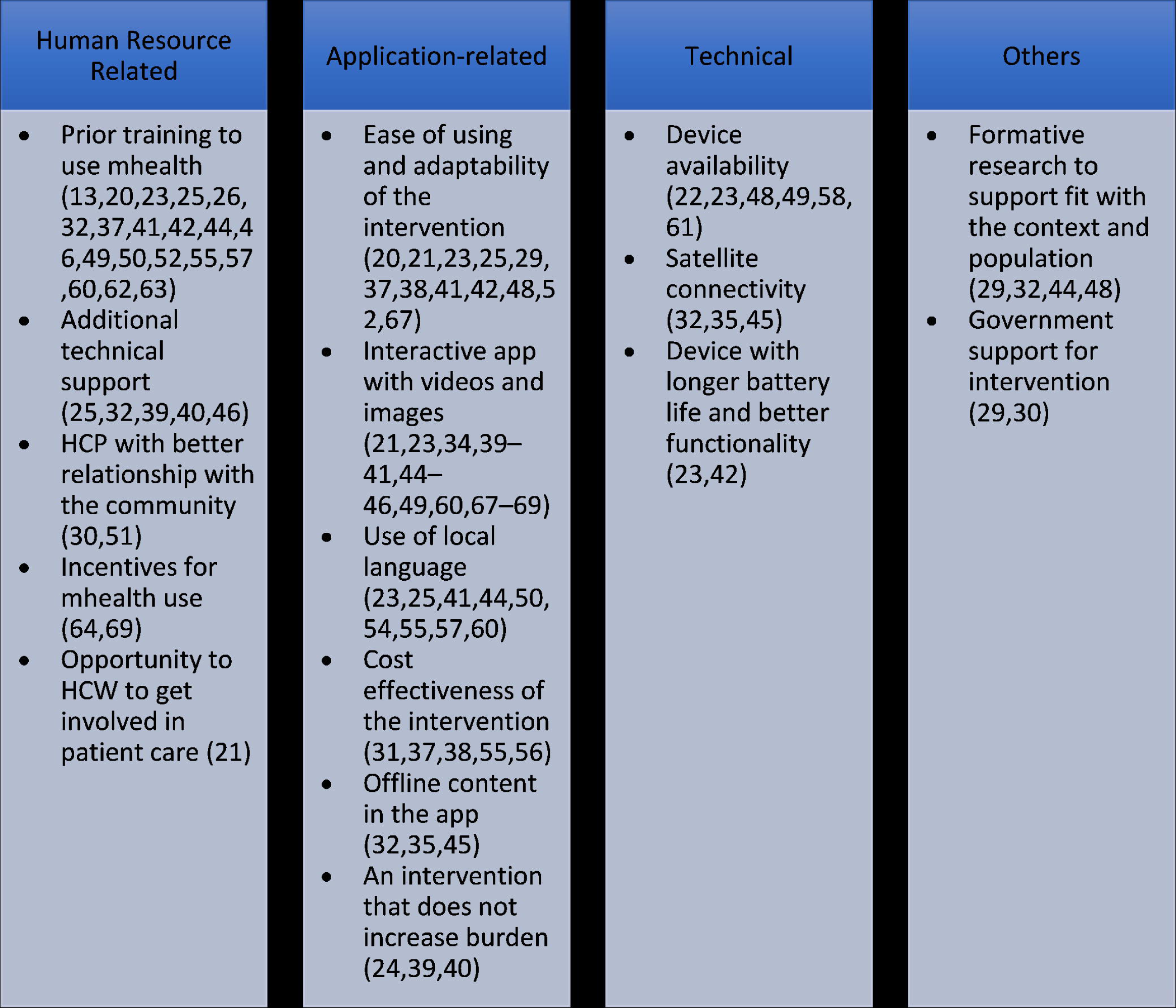
Facilitators of use of mHealth.

Low digital literacy (n=10)(13,22,26,32,38,41,44,46,65,69), malfunctioning of the software (n=13)(13,20–22,24,25,29,31,37,43,66,67,69), and poor network connectivity (n=14)(20–22,24,32,34,38,41,44,47,53,55,67,69) were the most common human resource-related, application-related, and technical barriers respectively. Stigma related to technology (n=4)(13,38,68,69), worsening of disease-related stigma due to the use of technology (n=3)(41,55,62), lack of formative research (n=1)(69), and lack of human touch due to the use of mhealth (n=1)(34) were other barriers that were identified. Other barriers are mentioned in **Fig 4**.

**Fig 4:**
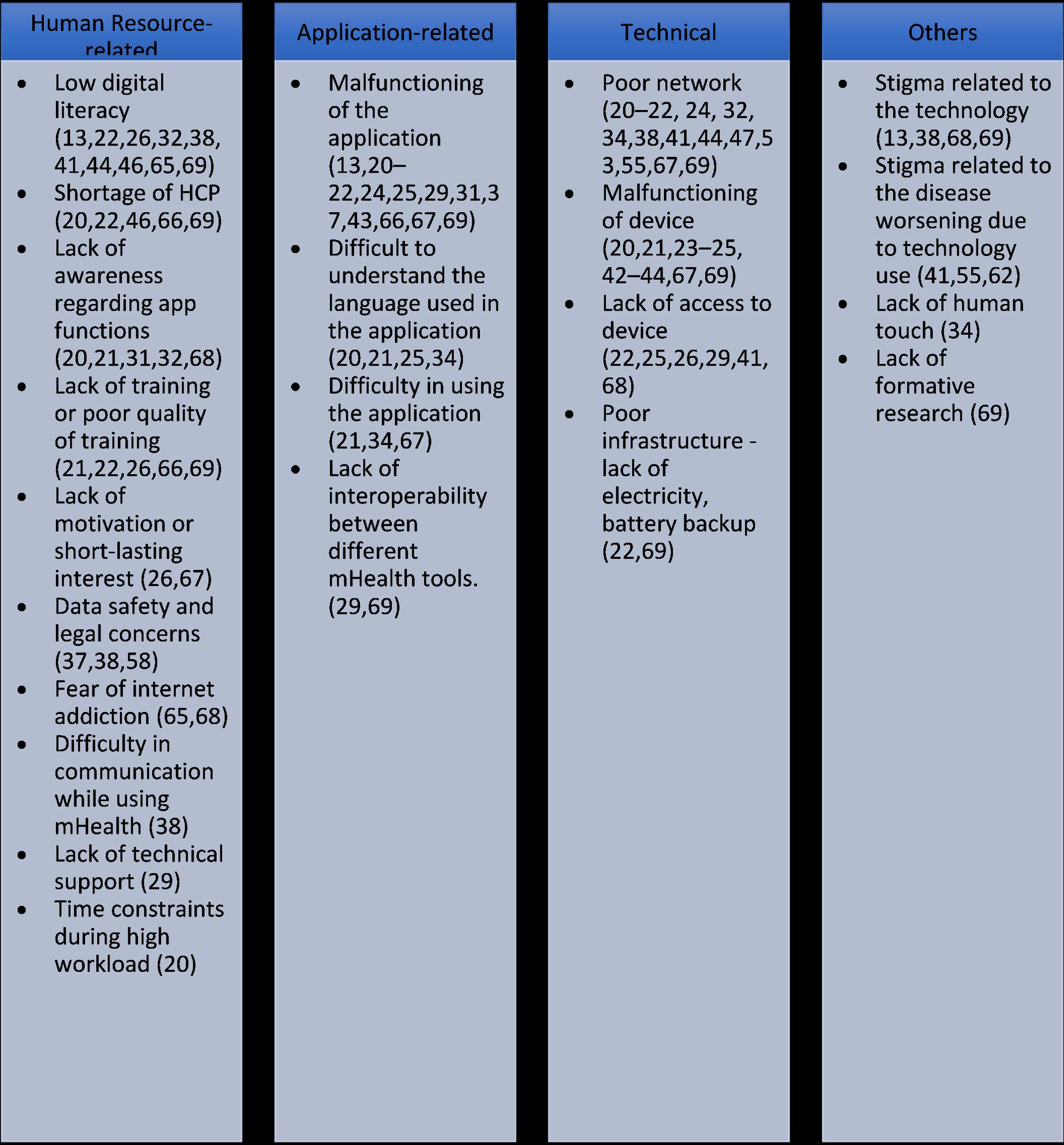
Barriers to the use of mHealth.

#### 2. Role of mHealth

The mHealth interventions were most commonly used for improving maternal and child healthcare (n=24)(13,20,22–25,27,33,34,36,39–43,53,54,59,60,63,64,66–68), followed by non-communicable diseases like diabetes, hypertension, and cancer (n=12)(26,28,30,31,46,47,50–52,55,57,65) and mental health (n=6).(21,32,44,49,61,62) Based on the WHO action framework on HRH, 29 (55%) studies focused on Human Resource (HR) management and aimed at improving the efficiency of available human resources.(13,22–24,28–30,33,35–40,42,43,46,51,54,55,57,58,61,64–67,69,71) Twenty-three (43%) studies involved mHealth interventions that aimed at the education and training of HCPs.(20,21,25–27,31,32,34,41,44,45,47–50,52,53,59,60,62,63,68,70) Only one study looked at the financial aspect of the intervention’s use by the HCPs.(56)

#### 3. Impact of mHealth Interventions and Attitude of HCPs towards Them

The use of mHealth impacted the practice of HCPs in various ways. Improvement in patient outcome was reported in 22 studies (20,24,28,33,36,39–43,46,47,49,51,53,54,60–63,69,71), improvement in knowledge of HCP in 18 studies (13,20,21,23,27,29,36,41,44,45,53,55,59,62,65,68–70), and improvement in work performance of HCP in 24 studies (13,20,22,23–25,29,33,38,40,41–43,45,46,48,52– 54,59,65,67,69,71). Studies also reported an improvement in confidence (n=7)(13,20,23,42,46,52,68) and communication (n=7)(13,40,41,43,50,54,58) while using mHealth interventions. The other impacts are mentioned in **Table 2**.

**Table 2:** Impact of telehealth interventions.

|  | **mHealth** | **Telemedicine** | **Tele-education** |
| --- | --- | --- | --- |
| **Variable** | **Number of studies**<br>**(n=53)** | **Number of studies**<br>**(n=25)** | **Number of studies**<br>**(n=28)** |
| Improvement in work performance | 24 (13,20,22,23–25,29,33,38,40,41–43,45,46,48,52– | 3 (76,85,92) | 5 (97,101,105,114,124) |
|  | 54,59,65,67,69,71) |  |  |
| Improvement in patient outcome | 22 (20,24,28,33,36,39–43,46,47,49,51,53,54,60–63,69,71) | 16 (74–78,81,82,85–87,89,91–94,96) | 2 (102,110) |
| Improvement in knowledge of HCP | 18 (13,20,21,23,27,29,36,41,44,45,53,55,59,62,65,68–70) | 3 (76,85,89) | 17 (97,100,101,104,105,107,108,110,111,114–118,120,122,124) |
| Increases social status/ recognition of work/care seeking/trust/reliability of HCP | 11 (13,22,23,25,36,39–41,43,52,69) | 2 (76,82) | - |
| Promotes better communication and relationship between HCP-HCP/HCP - patient | 7 (13,40,41,43,50,54,58) | 2 (76,95) | 1 (119) |
| Increase in confidence | 7 (13,20,23,42,46,52,68) | 1 (76) | 5 (104,107,110,111,115) |
| Flexibility to learn offered by the intervention | 7 (21,23,29,31,44,65,70) | 1 (83) | 4 (116,118,122,123) |
| Saves time | 6 (20,41,44,48,52,71) | 3 (87,92,94) | - |
| No diagnostic difference as compared to conventional techniques | 3 (47,57,61) | 3 (79,88,92) | 1 (124) |
| Decrease in workload/stress | 3 (20,39,46) | 2 (76,94) | - |
| Decreases travel | 2 (44,56) | 4 (78,91,92,94) | 1 (101) |
| Increased motivation of HCP due to the intervention | 2(52,69) | 1(77) | 1(122) |

Out of the 53 studies, 26 studies reported positive attitudes of HCPs toward mHealth interventions (13,22,25,26,29,31,32,34–38,40–46,49,52,53,55,58,68,69) whereas 1 study reported a negative attitude (65) and the remaining did not mention the attitude of the HCP toward the intervention. Nine studies reported that the HCP was satisfied with the intervention (24,29,31,32,43,54,55,61,68) and in two studies HCP mentioned that they would recommend the intervention to others. (23,31)

#### 4. Limitations of studies assessing the use of mHealth

Studies evaluating the use of mHealth interventions commonly cited inadequate sample size (n=10)(21,29,32,38–40,44,62,64,69), poor sampling techniques (n=9)(26,36,38– 40,44,62,64,69), and incomplete data (n=7)(33,39,40,42,47,50,63) as limitations. Desirability bias, as mentioned in 8 studies (20,21,23,25,32,36,39,64), could have resulted in a more positive outcome of the interventions being studied. Other limitations of studies are mentioned in **Table 3**.

**Table 3:** Limitations of studies included in the review.

|  | **mHealth** | **Telemedicine** | **Tele-education** |
| --- | --- | --- | --- |
| **Variable** | **Number of studies (n=53)** | **Number of studies (n=25)** | **Number of studies (n=28)** |
| Inadequate sample size | 10 (21,29,32,38–40,44,62,64,69) | 4 (74,81,85,86) | 4 (107,110,111,114) |
| Poor sampling technique | 9 (26,36,38–40,44,62,64,69) | 2 (72,73) | 3 (114,121,123) |
| Incomplete data/other data related constraints | 7 (33,39,40,42,47,50,63) | 2 (74,87) | 2 (106,108) |
| Poor study design | 6 (28,49,51,56,63,65) | 4 (80–82,96) | 2 (109,123) |
| Assessed perception only and not hard outcomes | 6 (20,26,31,36,42,62) | - | - |
| Short-term effect only assessed | 2 (31,68) | - | 3 (106,107,116) |
| Short duration of study | 3 (30,36,43) | - | 1 (115) |
| Resource constraints | 2 (23,35) | - | - |
| Desirability bias | 8 (20,21,23,25,32,36,39,64)/ | 1 (83) | - |
| Recall bias | 2 (43,56) | 1 (73) | - |
| Hawthorne bias | 1 (33) | 0 | - |
| Inappropriate study setting | 1 (44) | - | - |

### Telemedicine

Twenty-one studies (84%) were quantitative (72–77,79–82,84–89,91,93–96), 2 (8%) were qualitative (78,83), 1 (4%) followed mixed methodology (90) and 1 (4%) was a review study.(92) No study on the use of telemedicine by human resources for health (HRH) was published before 2011. A majority (64%)(74–78,82,84–87,89,91–93,95,96) of the studies were published after 2017 with the maximum (n=5, 20%)(75,77,84,86,91) number of studies being published in 2020. Nearly all the studies were conducted in tertiary care settings or teaching hospital settings (92%)(72,73,75–88,90–96), and only 1 study each was conducted in primary health centers (81), community health centers (73), HIV clinics (74), and non-governmental organization clinics. (89) The use of telemedicine interventions was most studied in Karnataka (n=5)(72,75,77,94,96) followed by Andhra Pradesh (n=3)(73,76,93) and Bihar (n=2).(86,87) (**Fig 5**) Findings from all included studies have been summarized in the S2 Table.

**Fig 5:**
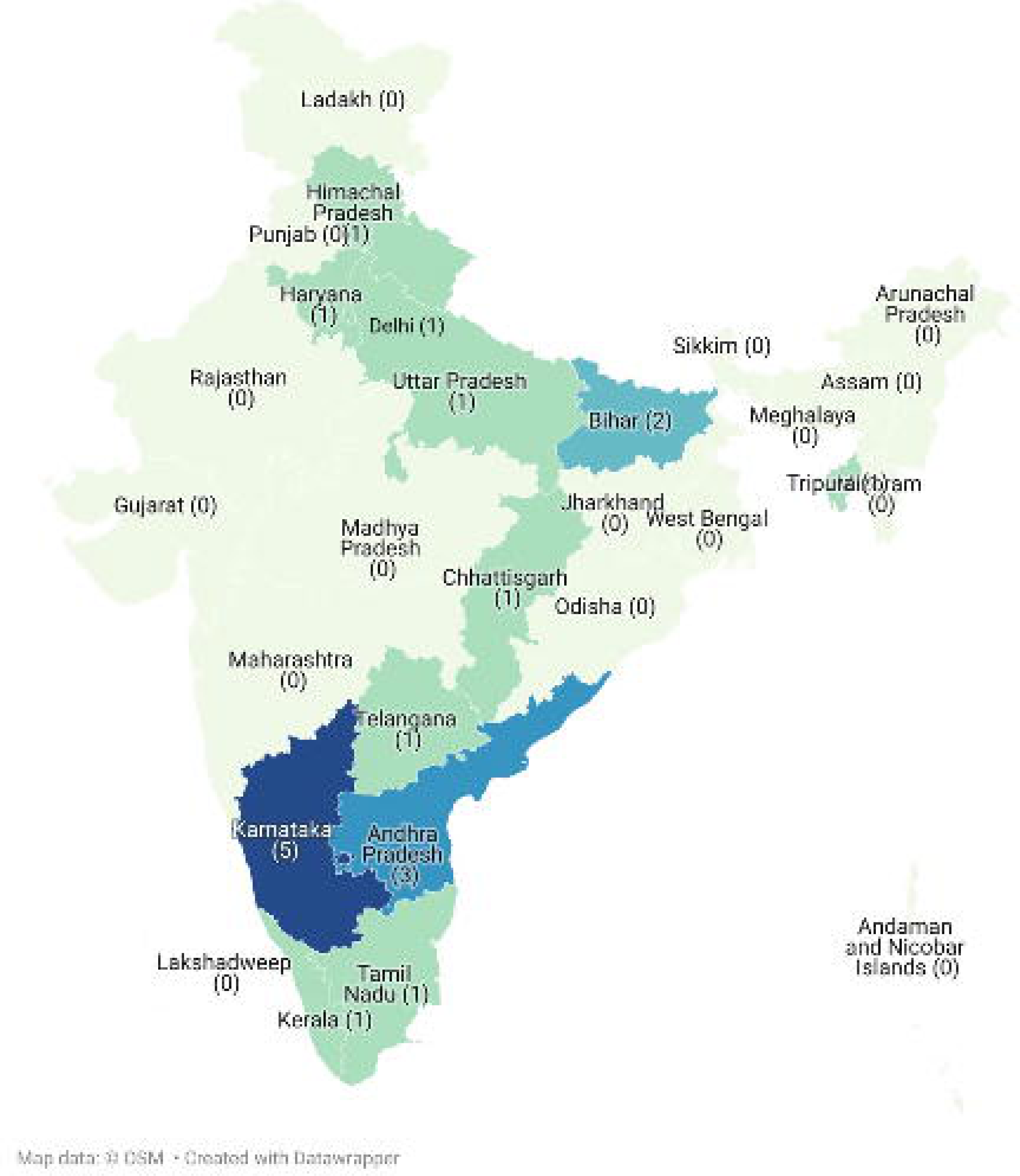
The number of study sites per state in India for telemedicine.

Telehealth was most commonly used by doctors (n=19)(72–77,79,80,82,83,85– 88,90,91,93,94,96) and focussed more on nurses (n=5)(82,84,86,89,96) than community healthcare workers (n=3)(72,77,95), allied health professionals (n=2)(72,89), and auxiliary midwife nurses (n=2). (72,81)

#### 1. Facilitators and barriers to the use of telemedicine

Prior training to use the telemedicine intervention (n=2)(75,93), use of local language(n=1)(89), and additional technical support (n=1)(76) were identified to be the human resource-related facilitators. The availability of satellite connectivity (n=1)(88) was a technical facilitator that improved the uptake of telemedicine. Cost-effectiveness (n=7)(74,75,78,85,87,93,94), and ease of use of the intervention (n=1)(92) were the application-related facilitators. (**Fig 6**)

**Fig 6:**
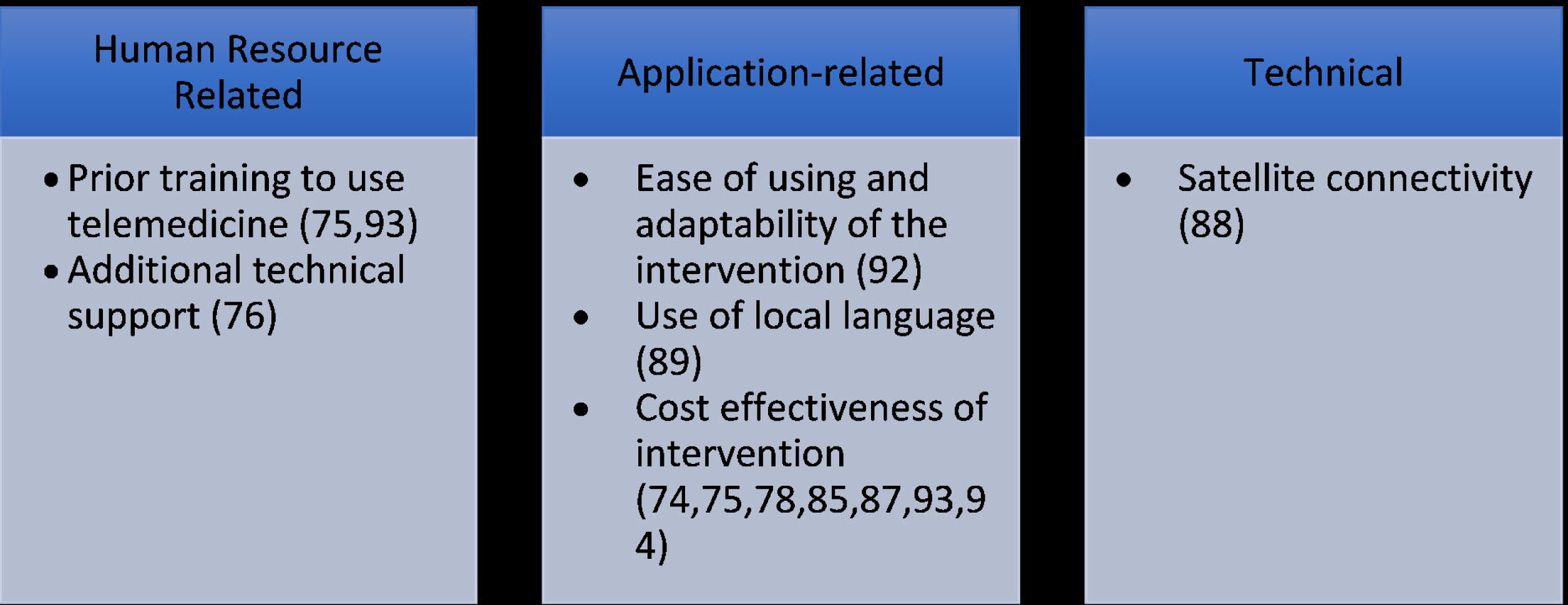
Facilitators of use of telemedicine.

Poor network connectivity (n=8)(73,76,81,84,87,92,94,96), difficulty in understanding English, the language used in the application (n=5)(84,86,87,92,93), and difficulty in communicating while using telemedicine (n=6)(76,83,86,87,93,95) were the most common technical, application-related, and human resource-related barriers respectively. Lack of human touch (n=5)(77,80,83,91,95) and stigma related to technology (n=1)(94) also acted as barriers to the uptake of telemedicine. Other barriers are mentioned in **Fig 7**.

**Fig 7:**
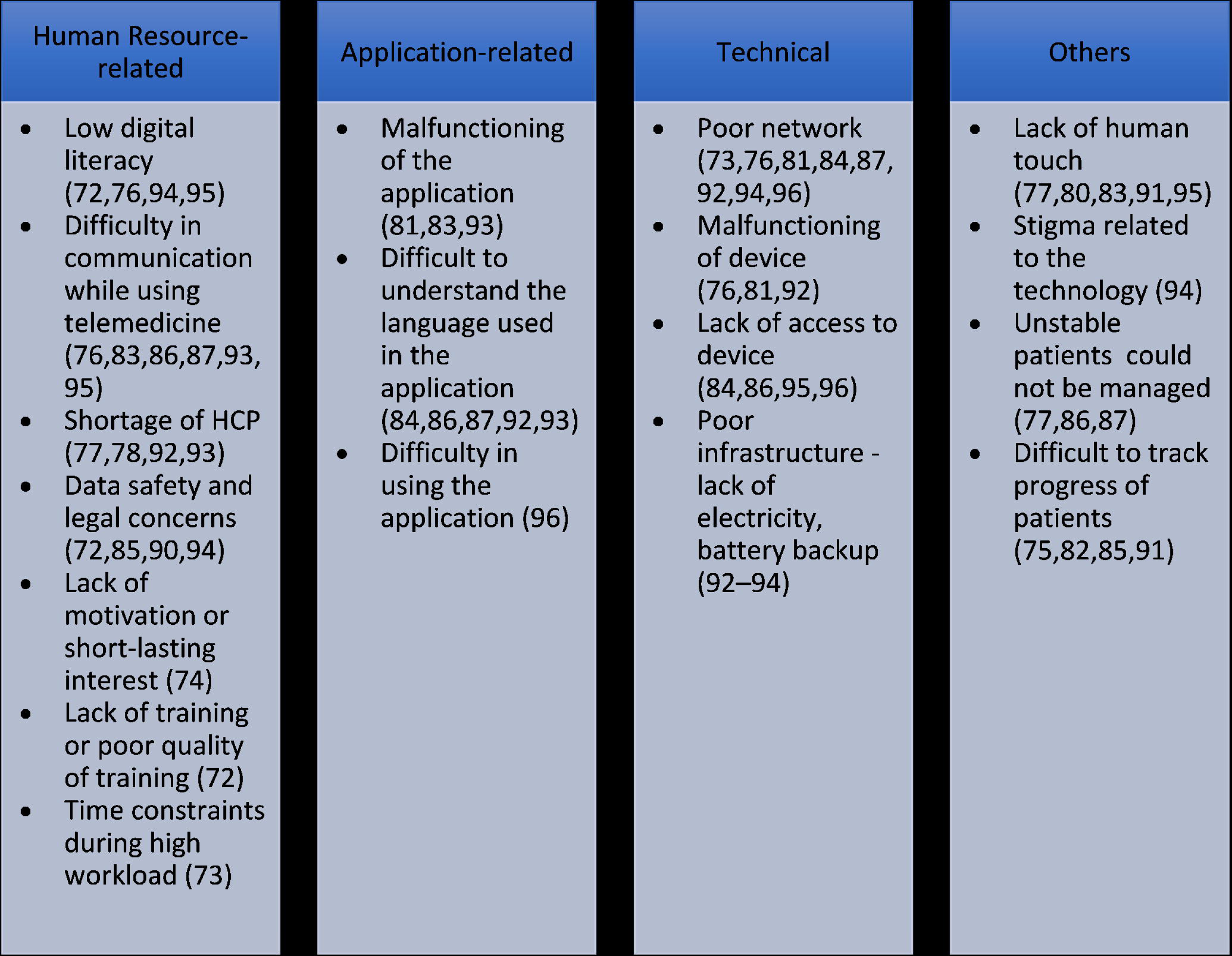
Barriers to the use of telemedicine.

#### 2. Role of telemedicine

Telemedicine was most commonly used for providing treatment for conditions related to maternal and child health (n=5)(76,80,82,84,86), non-communicable diseases (n=3)(88,91,94) like diabetes, hypertension, and cancer, and mental health (n=3).(77,78,95) While most studies focused on improving the efficiency and performance of the HRH (n=23)(73–89,91–96), 1 study focused on the knowledge and awareness regarding telemedicine in the HCPs(72) and 1 study addressed the policy and financial aspects of telemedicine.(90)

#### 3. Impact of telemedicine interventions and attitude of HCP towards them

Improvement in patient outcome (n=16)(74–78,81,82,85–87,89,91–94,96), improvement in knowledge of HCP (n=3)(76,85,89), and improvement in work performance (n=3)(76,85,92) were associated with the use of telemedicine. It also helped in reducing travel (n=4)(78,91,92,94) and when used for remote diagnosis, telemedicine showed no significant diagnostic difference when compared with conventional diagnostic modalities (n=3)(79,88,92). The other impacts are mentioned in **Table 2**.

Out of the 25 studies, 13 studies reported positive attitudes of HCPs toward telemedicine interventions (76–78,83,85–87,91–96) whereas 1 study reported a negative attitude (90) and the remaining did not mention the attitude of the HCP toward the intervention. Twelve studies reported that the HCP was satisfied with the intervention (76,77,82–84,86,87,91– 94,96) and in 2 studies HCPs mentioned that they would recommend the intervention to others. (86,87)

#### 4. Limitations of studies assessing the use of telemedicine

Inadequate sample size (n=4)(74,81,85,86), poor study design (n=4)(80–82,96), and poor sampling techniques (n=2)(72,73) were the most commonly cited limitations in the studies included. Other limitations of studies are mentioned in **Table 3**.

### Tele-education

Twenty-four studies (85.7%) were quantitative (97–102,105–109,111,112,115–124), 2 (7.1%) were qualitative (104,113), 1 (3.6%) followed mixed methodology (114) and 1 (3.6%) was a review study.(103) No study on the use of tele-education was published before 2009. A majority (72%)(97,98,100–102,104–109,111–113,116–118,121–123) of the studies were published after 2017 with the maximum (n=10, 36%)(97,98,101,106,108,109,117,121– 123) number of studies being published in 2021. Nearly all the studies were conducted in tertiary care settings or teaching institutes (86%)(98–100,102–107,109–117,119–124), and only 3 studies were conducted in primary health centers (101,108,118) and 2 in community health centers. (101,108) The use of tele-education was most studied in Karnataka (n=4)(100,104,111,112) and Delhi (n=4). (102,111,117,120) (**Fig 8**) Findings from all included studies have been summarized in the S3 Table.

**Fig 8:**
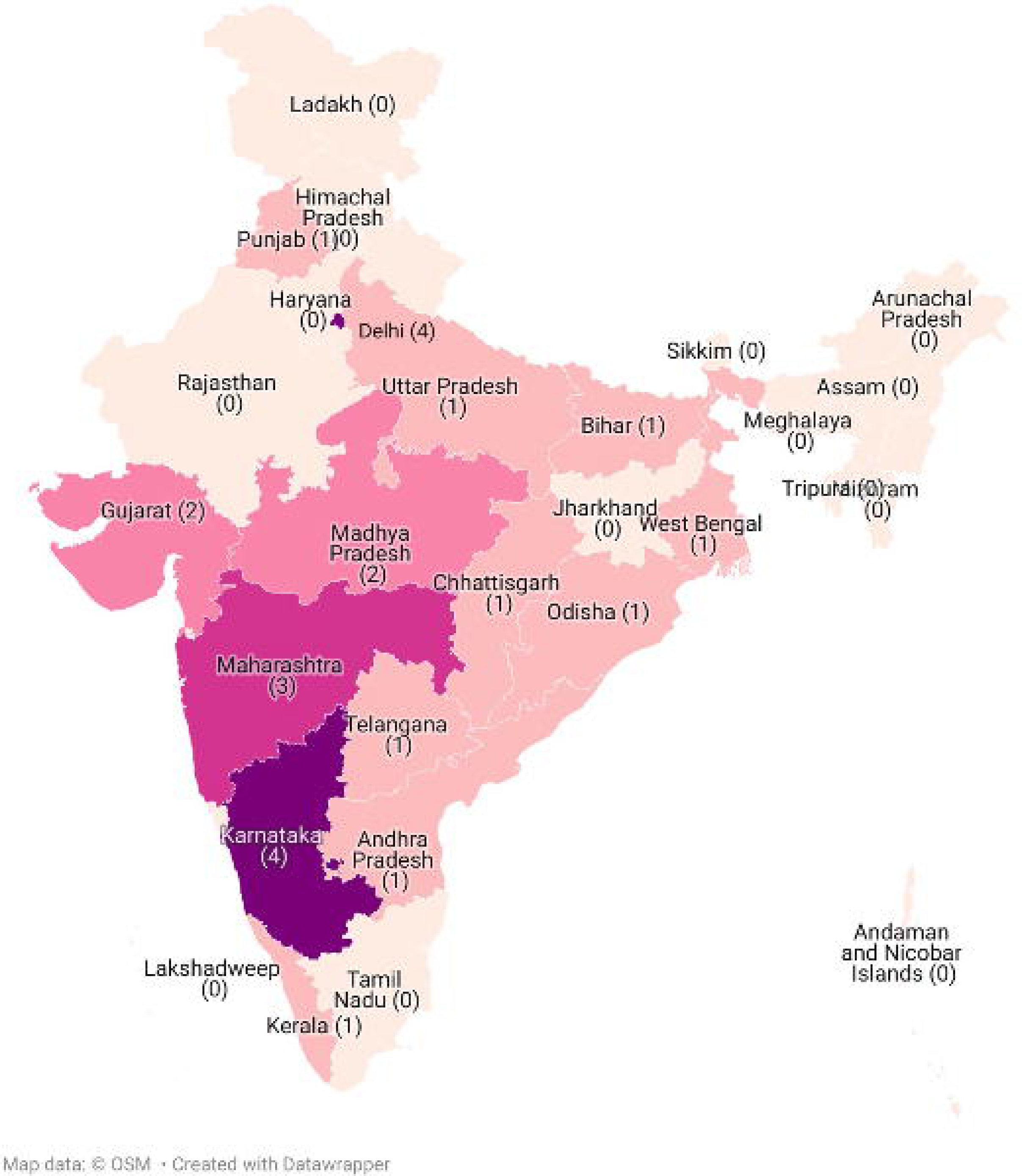
The number of study sites per state in India for tele-education.

Tele-education services were most commonly meant for doctors (n=16)(97–103,106,109– 111,117–119,121,124) followed by nurses (n=9)(98,105,106,109,111,117,121,122,124), community healthcare workers (n=8)(103–105,107,114–116,123), allied health professionals (n=5)(98,109,114,121,124), and auxiliary midwife nurses (n=4).(108,111,112,121)

#### 1. Facilitators and barriers to the use of tele-education

Similar to telemedicine, prior training to use the tele-education intervention (n=2)(102,118) and ease of using the intervention (n=2)(112,122) were the most common human resource-related, and application-related facilitators respectively. Availability of a device (n=2)(110,118) was identified to be a technical facilitator. Formative research prior to designing the intervention (n=1)(104) also helped in increasing its uptake as the formative research helped in addressing the needs of the participants. (**Fig 9**)

**Fig 9:**
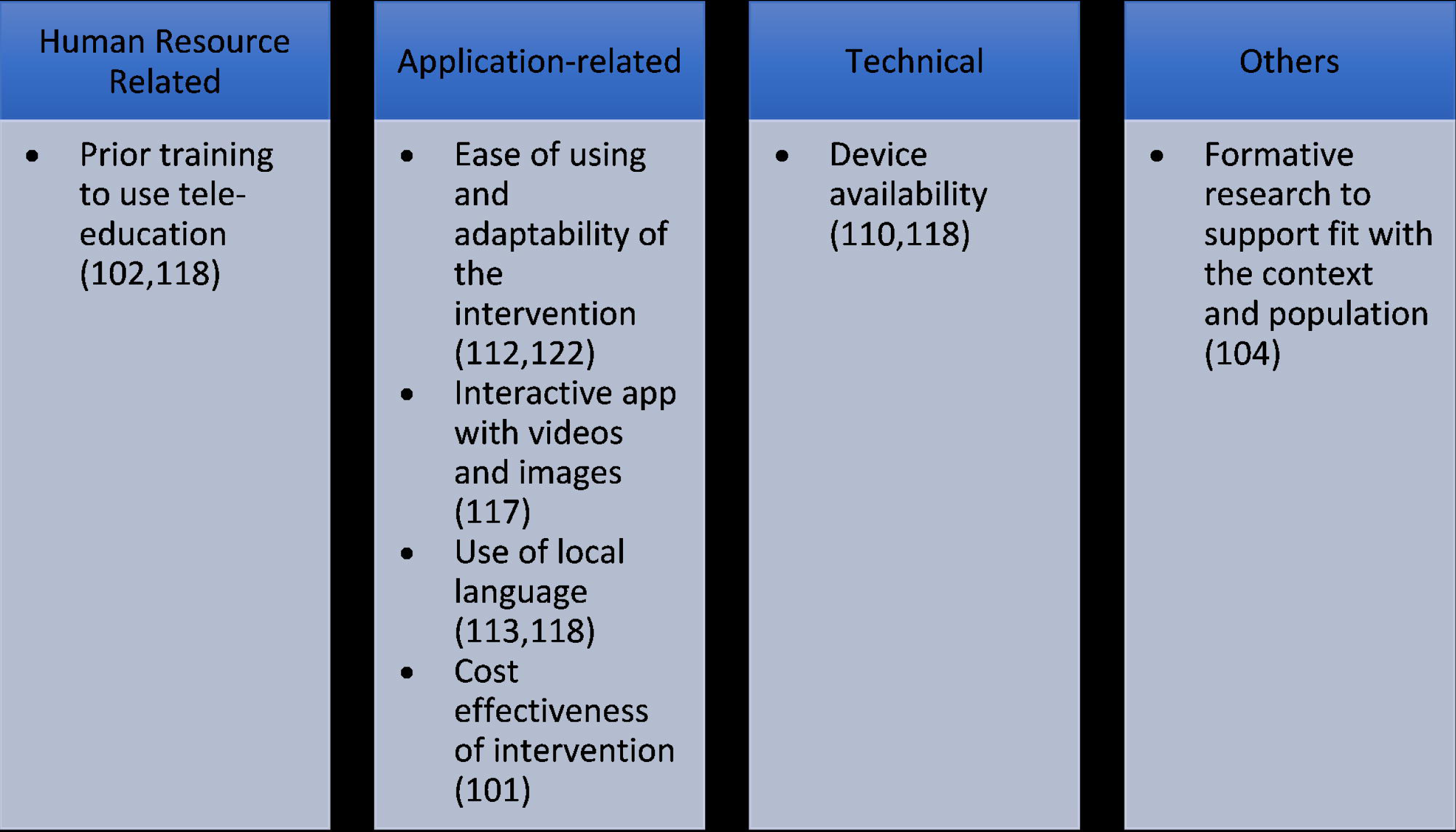
Facilitators of use of tele-education.

Similar to telemedicine, low digital literacy (n=2)(104,115), and poor network connectivity (n=11)(98–100,103,104,109,111,113,117,120,122) were the most common human resource-related, and technical barriers respectively. Difficulty in understanding English (n=2)(111,114), the language commonly used for the applications, and malfunctioning of the software (n=2)(111,113) were application-related barriers. Other barriers are mentioned in **Fig 10**.

**Fig 10:**
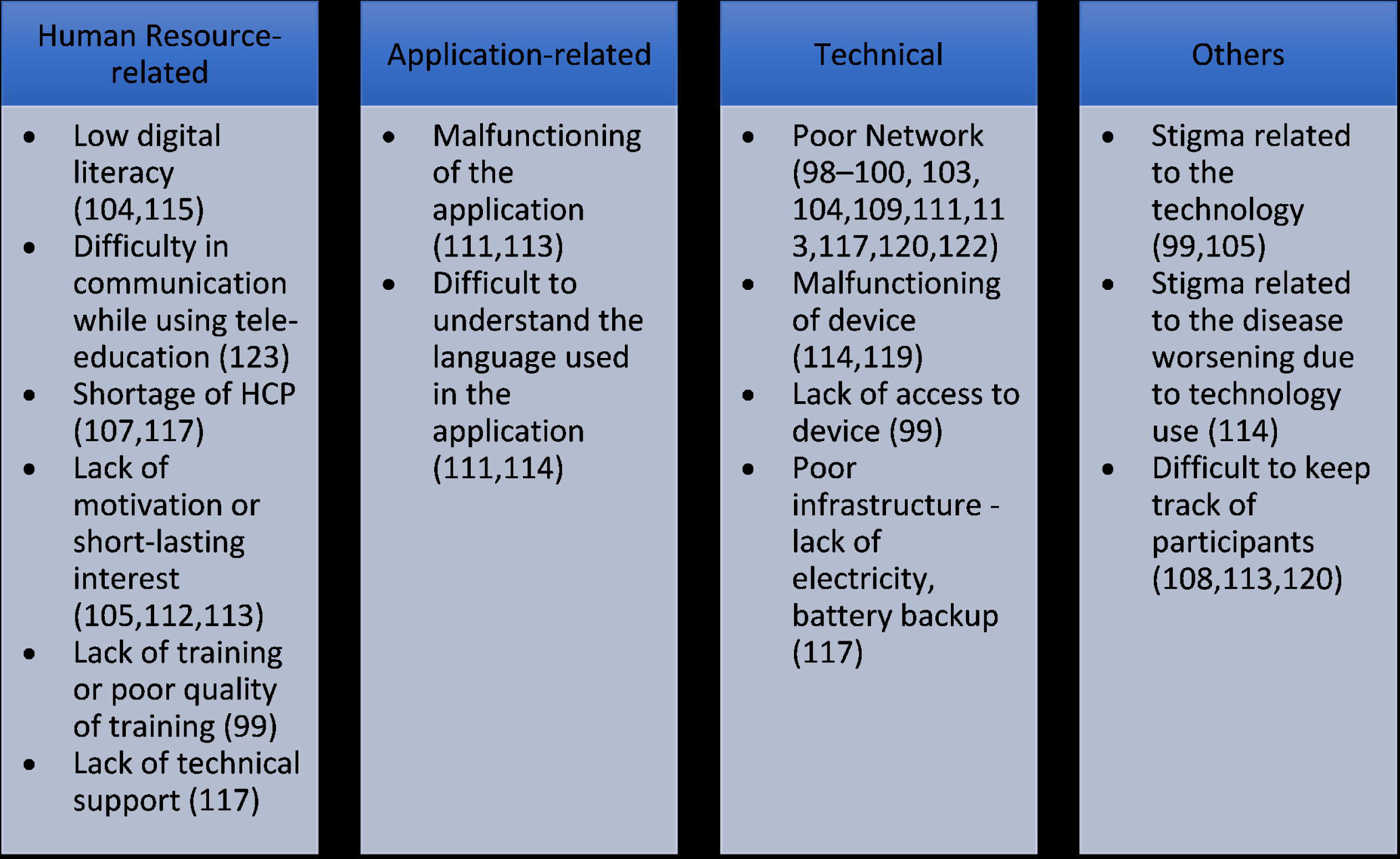
Barriers to the use of tele-education.

#### 2. Role of tele-education interventions

Tele-education services were most commonly used for educating about mental health disorders (n=9)(97,98,101,102,104,107,109,111,121) followed by non-communicable diseases like diabetes, hypertension, and cancer (n=4)(100,113,116,118) and maternal and child healthcare (n=4).(104,114,115,120) Two studies each focused on educating about oral health problems (105,106) and HIV (110,114) and one study addressed teleteaching for orthopedics (112), critical care (108), COVID-19 (117), palliative care (122) and cardiology.(124) Four studies did not mention what tele-education was used for.(99,103,119,123)

#### 3. Impact of tele-education Interventions and Attitude of HCP towards Them

Tele-education resulted in an improvement in knowledge of HCP (n=17)(97,100,101,104,105,107,108,110,111,114–118,120,122,124) and an improvement in the work performance of HCP (n=5).(97,101,105,114,124) Its use also resulted in improvement in the confidence (n=5)(104,107,110,111,115) and communication (n=1)(119) of HCPs. The other impacts are mentioned in **Table 2**.

Out of the 28 studies, 15 studies reported positive attitudes of HCPs toward tele-education interventions (97,100,104,105,108–112,114–116,122–124) whereas 1 study reported a negative attitude (99) and the remaining did not mention the attitude of the HCP toward the intervention. Nine studies reported that the HCP was satisfied with the intervention (97,107,110,111,114,115,119,120,124) and in 3 studies HCP mentioned that they would recommend the intervention to others.(110,115,122)

#### 4. Limitations of studies assessing the use of tele-education

Limitations of studies included were similar to the limitations cited by studies that assessed telemedicine with inadequate sample size (n=4)(107,110,111,114), poor study design (n=2)(109,123), and poor sampling techniques (n=3)(114,121,123) being the most commonly cited limitations. Other limitations of studies are mentioned in **Table 3**.

## Discussion

Following the rapid digitalization of healthcare, mostly following the COVID-19 pandemic, this scoping review looks at the facilitators and barriers to the application of telehealth for various health issues in the Indian health system. Even though a wide variety of interventions in the form of mHealth, telemedicine, and tele-education have been explored, only 8 states/union territories were the sites for most of the interventions. The use of telehealth by doctors, nurses, and community health workers was commonly addressed and literature on the use of the same by allied health professionals and non-medical healthcare workers was limited. Telehealth was most commonly used for HRH management aiming to improve the efficiency of available human resources. Maternal and child health, non-communicable diseases like diabetes, hypertension, obstructive airway disease, and cancer, and mental health were common areas of focus for the use of telehealth. Few studies looked at the use of telehealth for the provision of acute medical care, follow-up of patients after discharge, provision, and monitoring of home-based palliative care, and improvement in treatment compliance of patients with HIV and tuberculosis. Studies conducted globally, have also assessed the utility of telemedicine, mHealth, and tele-education for similar diseases and conditions as done by the studies in India. (125–127)

This review brings to light multiple facilitators and barriers to telehealth adoption and use. The findings could help in the modification of national policies and guidelines which currently are not very robust. (128) Moreover the facilitators and barriers identified for mHealth, telemedicine, and tele-education are similar. An understanding of the facilitators and barriers emphasizes the need for understanding the same at multiple dimensions especially focusing on the facilitators and barriers related to Human resources for health (HRH), Infrastructure, and Technology. The facilitators and barriers identified in our review are similar to those reported previously in the context of telehealth in LMICs. Technical and infrastructural barriers in the form of internet access, device access, connectivity issues, poor battery life, and unstable electricity supply contribute to a major road back in implementing telehealth services in LMICs.(129–131) This is especially important in the context of India, where over 70% of the population resides in rural areas which are highly vulnerable to the aforementioned barriers. (132) An increase in network coverage should also be associated with a push for gender equality as it is seen that women have lesser access to mobile phones and other technologies. (133) In terms of HRH barriers, previous studies have identified HRH shortage, insufficient training, and skills, additional workload, lack of motivation lack of technical support, lack of integration with other government systems, and data safety and legal concerns.(129–131,134,135) Additionally our study provides deeper insights into barriers faced by the provider like fear of internet addiction, language barriers, and malfunction of applications. Barriers concerning the lack of human touch and stigma related to subpar patient care have also been previously raised by a systematic review conducted by Kruse C. S. et al.(136) Previous studies have also reported financial barriers in the form of sponsorships and funding, capital expenses for technology start-up and maintenance, and budget constraints. (129,131)

In the Indian context, Government support and funding for telehealth interventions have been found to be an important facilitator for their implementations as reported by 3 studies included in our review. However, funding towards health overall is still largely limited in India as only 2.1% of the Gross Domestic Product (GDP) is invested in the public healthcare sectors. (137) Initiatives like the ‘G-20 Digital Innovation Alliance’ show promise in encouraging digital health startups by providing grants, sponsorship, and collaboration opportunities in order to strengthen the telehealth scenario. (138) Previous studies have also shown that a strong commitment from the governments towards supporting and financing telehealth has been one the major facilitators. (135) HRH & Application related facilitators in the form of prior training, technical support, use of local language, and better user interface which have been shown to be important facilitators (135), were also reported in over one-third of the studies from our review. Additionally, providing incentives for telehealth use, use of offline material, balanced overload, and the relationship of CHW with the community were also found to be other important facilitators in our review. Formative research to support fit with the context and population was seen as an important facilitator for telehealth in India; this emphasizes the need for regional research as well as customizing the intervention as per the setting. Fifteen studies also emphasized the cost-effectiveness of telehealth interventions, which serve as a vital facilitator in resource-constrained settings like India.

Our review reported a strong impact of telehealth on patient care in terms of better patient outcomes, treatment compliance, and disease knowledge. It reduced travel constraints and improved accessibility for both patients and healthcare providers which has also been shown to improve the previously mentioned outcomes. (139,140) Specifically for healthcare workers, a greater number of studies showed that the use of telehealth improved their performance, confidence, and patient communication. Globally as well, multiple studies have reported similar positives. (141,142) However, a few studies also highlight contradicting findings which are multifactorial and scenario-dependent. (143,144) Studies assessing the use of telehealth diagnostics have also shown promising results in India which are similar to other studies conducted globally. (127,145–147) Our review also highlights the utility of digital health interventions in the overall education and skill training of HRH personnel. As shown by multiple studies conducted during the COVID-19 pandemic, remote learning facilitated by tele-education has proven to be an effective tool that can be harnessed even after the pandemic in order to make education and training more convenient and accessible. (145,148–150)

The need for decentralized healthcare planning was identified following the Covid-19 pandemic. (7) Our review identifies that with respect to telehealth, the generation of scientific literature on facilitators and barriers has been concentrated in a few states only. As the government is pushing for the digitization of healthcare through the Ayushman Bharat Digital Health Mission (151), it is important to understand the barriers and facilitators not just at the national level but also at the community level. More comparable evidence needs to be generated in order to understand local factors affecting the implementation of telehealth in India.

### Strengths and Limitations

A few of the strengths of our studies are the use of a robust search strategy and the inclusion of a large number of studies. While previous reviews have assessed the overall utility of telemedicine, our review specifically looks at telehealth, which covers broader interventions, and its utility in the context of HRH providers. In the Indian context use of mHealth, telemedicine, and tele-education by community health workers has been an important highlight of our review. However, the findings of this review must be interpreted in the context of the following limitations. Firstly, studies included mainly assess the utility of public health, mHealth, telemedicine, and tele-education portals, while there are multiple private applications that are usually accessible to upper socioeconomic strata, whose utility hasn’t been assessed. Secondly, since this is a scoping review we only provide a brief overview of the facilitators and barriers, and an in-depth analysis of study outcomes, meta-analysis, and critical appraisal of the risk of bias was not performed for the studies included. Thirdly, while analyzing the number of studies from each state, data was not available for 28 studies, and 8 studies were conducted in multiple states with no mention of the names of the states involved. Finally, our search strategy, though comprehensive, is limited only to PubMed, which might have led to the exclusion of a few studies available on other databases like Scopus and EMBASE.

## Conclusion

Use of telehealth has not been studied uniformly across India. Systematic efforts need to be taken to anticipate and address barriers and implement telehealth intervention in ways to facilitate its uptake. Future studies should focus on looking at region-specific, intervention-specific, and health cadre-specific barriers and facilitators for the use of telehealth in order to promote decentralized decision-making for successfully implementing telehealth interventions in India.

## Supporting information

S1 Panel

S2 Panel

PRISMA-ScR Checklist

## Data Availability

All data produced in the present study are available upon reasonable request to the authors.

## Abbreviations

HCP: Health Care Provider
UHC: Universal Health Coverage HRH - Human Resources for Health

## Acknowledgments

We want to thank Dr. Dipanwita Sengupta, Lancet Citizens’ Commission Member for her support during the review process. We also want to thank Ms. Vashumathi Sriganesh from QMed Knowledge Foundation, Mumbai for providing her support for building the search strategy.

