## Supplementary material for "Barriers and Facilitators for the Use of Telehealth by Healthcare Providers (HCP) in India - A Scoping Review": S1 Panel

**S1 Panel: PubMed search strategy**

**PubMed search strategy**

The search terms comprise of all concepts and terminologies related to the six main components (human resource management systems, policy, finance, education, partnership and leadership) in the HRH action framework AND all terms related to different cadres of

healthcare workers AND combined with search terms related to universal health coverage, healthcare AND India.

(((("gender equity"[MeSH Terms] OR "health care facilities, manpower, and services"[MeSH Terms] OR "health services administration"[MeSH Terms] OR "health care economics and organizations"[MOSH Terms] OR "job satisfaction"[MeSH Terms] OR "professional competence"[MeSH Terms] OR "professional role"[MeSH Terms] OR "work performance"[MeSH Terms] OR "career development"[Title/Abstract] OR "career path*"[Title/Abstract] OR "career progression"[Title/Abstract] OR "career trajectory"[Title/Abstract] OR "deployment"[Title/Abstract] OR "employee motivation"[Title/Abstract] OR "employee relation*"[Title/Abstract] OR (("employee s" [All Fields] OR "occupational groups"[MeSH Terms] OR ("occupational" [All Fields] AND "groups"[All Fields]) OR "occupational groups"[All Fields] OR "Employee" [All Fields] OR "employees" [All Fields]) AND "remediation"[Title/Abstract]) OR "employee retention"[Title/Abstract] OR "employee satisfaction"[Title/Abstract] OR "employee selection"[Title/Abstract]) AND "employee wellbeing"[Title/Abstract]) OR "gender disparity"[Title/Abstract] OR "gender equity"[Title/Abstract] OR "gender inequality"[Title/Abstract] OR "gender inequity"[Title/Abstract] OR "grievance*"[Title/Abstract] OR "health administration"[Title/Abstract] OR "health care facilit*"[Title/Abstract] OR "health resource*"[Title/Abstract] OR "health service*"[Title/Abstract] OR "health care economics"[Titleabstract] OR "health care organization*"[Title/Abstract] OR "hiring"[Title/Abstract] OR "human resource management"[Title/Abstract] OR "information system*"[Title/A_bstract] OR "job expectation*"[Title/Abstract] OR "job satisfaction"[Title/Abstract] OR "performance appraisal" [Title/ Abstract] OR "personnel system*"[Title/Abstract] OR "Productivity"[Title/Abstract] OR "professional competence"[Title/Abstract] OR "professional rolelTitle/Abstract] OR "promotion"[Title/Abstract] OR "recruitment"[Title/Abstract] OR "skill-mix" [Title/Abstract] OR "staffing norm*"[Title/Abstract] OR "Supervision"[Title/Abstract] OR "work environment"[Title/Abstract] OR "work performance"[Title/Abstract] OR "workforce

plan*" Title/Abstract] OR "working condition*"[Title/Abstract] OR "workload assessment"[Title/Abstract] OR "workplace safety"[Title/Abstract[ OR ("accreditation"[MeSH Terms] OR "Certification"[MeSH Terms] OR "health care reform"[MeSH Terms] OR "health policy"[MeSH Terms] OR "accredit*"[Title/Abstract] OR "Certification"[Title/Abstract] OR "employment law*"[Title/Abstract] OR "employment preference"[Title/Abstract] OR "employment rule*"[Title/Abstract] OR "financial decision*"[Title/Abstract] OR "health polic*"[Title/Abstract] OR "healthcare reform*"[Title/Abstract] OR "licensing"[Title/Abstract] OR "policy decision*"[Title/Abstract] OR "political decision*" [Title/ Abstract] OR "professional standard*"[Title/Abstract] OR scope of practice[tiab]) OR ("healthcare financing"[MeSH Terms] OR "Income"[MeSH Terms] OR "workers compensation"[MeSH Terms] OR "allowance*"[Title/Abstract] OR "budget"[Titleabstract] OR "Compensation"[Title/Abstract] OR "employee benefit*"[Title/Abstract] OR "expenditure*"[Title/Abstract] OR "financial resource*"[Title/Abstract] OR "fund*"[Title/Abstract] OR "healthcare financ*"[Title/Abstract] OR "incentive*"[Title/Abstract] OR "Income"[Title/Abstract] OR "package*"[Title/Abstract] OR "perk*"[Title/Abstract] OR "perquisite*"[Title/Abstract] OR "reward*"[Title/Abstract] OR "salar*"[Title/Abstract] OR "workers compensationlTitle/Abstractl) OR ("health facilities"[MeSH Terms] OR "schools, health occupations"[MeSH Terms] OR "education, professional"[MeSH Terms] OR "schools, public health"[MeSH Terms] OR "competenc*"[Title/Abstract] OR "continuing professional education"[Title/Abstract] OR "Curriculum"[Title/Abstract] OR "Employability"[Title/Abstract] OR "health facilit*ITitle/Abstract] OR ((("Health"[MeSH Terms] OR "Health"[All Fields] OR "health s"[All Fields] OR "healthful"[All Fields] OR "healthfulness" [All Fields] OR "healths"[All Fields]) AND "profession*"[All Fields]) AND "education"[Title/Abstract]) OR "professional education" [Title/ Abstract] OR "public health school*"[Title/Abstract] OR "scholarship*"[Title/Abstract] OR "training institution*"[Title/Abstract] OR "Training"[Title/Abstract]} OR ("public private sector partnerships"[MeSH Terms] OR "committee"[Title/Abstract] OR "community involvement"[Title/Abstract] OR "Cooperation"[Title/Abstract] OR "Coordination"[Title/Abstract] OR "Governance"[Title/Abstract] OR "multi-stakeholder" [Title/Abstract] OR "partnership*"[Title/Abstract] OR "private sector"[Title/Abstract] OR "public sector"[Title/Abstract]) OR ("accountability"[Title/Abstract] OR "collaborat*"[Title/Abstract] OR "empowerment"[Title/Abstract] OR "health advocates"[Title/Abstract] OR "Leadership"[Title/Abstract] OR "professional association*"[Title/Abstract] OR "strateg*"[Title/Abstract] OR "teamwork"[Title/Abstract])) AND ("administrative personnel"[MeSH Terms] OR ("counselors"[MeSH Terms] OR "counseling"[MeSH Terms]) OR "Employment"[MeSH Terms] OR "foreign professional personnel"[MeSH Terms] OR "health occupations"[MeSH Terms] OR "health personnel"[MeSH Terms] OR "housekeeping, hospital"[MeSH Terms] OR "occupations"[MeSH Terms] OR "Workforce"[MeSH Terms] OR "accredited social health activist*"[Title/Abstract] OR "administrative assistant*"[Title/Abstract] OR "administrative personnel"[Title/Abstract] OR "allied professional*"[Title/Abstract] OR "Allopathic"[Title/Abstract] OR "alternative medicine"[Title/Abstract] OR "ambulance drivers"[Title/Abstract] OR "Ayurvedic"[Title/Abstract] OR "community worker*"[Title/Abstract] OR "complementary medicine"[Title/Abstract] OR "compounder*"[Title/Abstract] OR "counselor*"[Title/Abstract] OR "dentist*"[Title/Abstract] OR "dietician*"[Title/Abstract] OR "doctor*"[Title/Abstract] OR "Employment"[Title/Abstract] OR "faith healer*"[Title/Abstract] OR "foreign professional personnel"[Title/Abstract] OR "frontline health workforce"[Title/Abstract] OR "healers" [Title/ Abstract] OR "health care practitioner*"[Title/Abstract] OR "health manager*"[Title/Abstract] OR "health occupation*"[Title/Abstract] OR "health personnel"[Title/Abstract] OR "health worke&"[Title/Abstract] OR (("delivery of health care"[MeSH Terms] OR rdelivery"[All Fields] AND "Health"[All Fields] AND "Care"[All Fields]) OR "delivery of health care" [All Fields] OR "Healthcare" [All Fields] OR "healthcare s" [All Fields] OR "healthcares"[All Fields]) AND "manpower"[Title/Abstract]) OR "healthcare worker"[Title/Abstract] OR "horneopath*"[Title/Abstract] OR "homoeopath*"[Title/Abstract] OR "hospital

housekeeping"[Title/Abstract] OR "human resource*"[Title/Abstract] OR "human resources for health"[Title/A_bstract] OR "integrative medicine"[Title/Abstract] OR "laboratory personnel"[Title/Abstract] OR "labour*"[Title/Abstract] OR "medical officer*"[Title/Abstract] OR "Midwives"[Title/Abstract] OR "naturopath*"[Title/Abstract] OR "nurse*"[Title/Abstract] OR "occupation*"[Title/Abstract] OR "office staff"[Title/Abstract] OR "optometrist*"[Title/Abstract] OR "pharmacist*"[Title/Abstract] OR "phlebotomist*"[Title/Abstract] OR "physician assistant*" [Title/ Abstract] OR "physician*"[Title/Abstract] OR "physiotherapist*"[Title/Abstract] OR "psychologist*"[Title/Abstract] OR "quack*"[Title/Abstract] OR "sanitary workee"[Title/Abstract] OR "secretary"[Title/Abstract] OR "security personnel"[Title/Abstract] OR "SiddhalTitle/Abstract] OR "social worker*"[Title/Abstract] OR "surgeon*"[Title/A_bstract] OR "therapist*"[Title/Abstract] OR "traditional healer*"[Title/Abstract] OR "traditional medicine practitioner*"[Title/Abstract] OR "Unani"[Title/Abstract] OR "village worker*"[Title/Abstract] OR ("ward" [All Fields] AND "attendee"[Title/Abstract]) OR "Workforce"[Title/Abstract] OR "Yoga"[Title/Abstract]) AND ("health care"[Title/Abstract] OR "Healthcare"[Title/Abstract] OR "Health"[Title/Abstract]) AND ("India"[Title/Abstract] OR "Indian"[Title/Abstract] OR

"India" [Place of Publication] OR "India" [Affiliation] OR "Indian"[Affiliation])) AND (2001:2022[pdat])
