## Supplementary material for "Barriers and Facilitators for the Use of Telehealth by Healthcare Providers (HCP) in India - A Scoping Review": S2 Panel

**S2 Panel:  HR Cadres**

All doctors and nurses (all doctors plus nurses and midwives)

All doctors (allopathic plus AYUSH doctors)

Ayurvedic, homeopathic and unani (AYUSH) doctors

Dental Specialists

All Health Professionals

Nursing Professionals

Medical Assistants

Sanitarians

Dieticians and Nutritionists

Optometrists and Opticians

Dental Assistants and Therapists

Physiotherapists and Related Associate Professionals

Pharmaceutical Assistants

Modern Health Associate Professionals (except Nursing)

Nursing Associate Professionals

Midwifery Associate Professionals

Traditional Medicine Practitioners

Faith Healers
