## Supplementary material for "Barriers and Facilitators for the Use of Telehealth by Healthcare Providers (HCP) in India - A Scoping Review": S1 Table

S1 Table: Study characteristics for studies on mHealth as mentioned by the authors.

| Study identification* | Year of Publication | Study design and setting | States Studied | Population studied | Sample size | Primary objectives | Positive findings/Facilitators | Barriers | Implications for healthcare delivery/ universal health coverage | Limitations of the study |
| --- | --- | --- | --- | --- | --- | --- | --- | --- | --- | --- |
| Usmanova et al (20) | 2020 | Qualitative (cross-sectional study) | Rajasthan, Madhya Pradesh | Medical Officers, Staff Nurses, Labor room supervisors | 44 | Acceptability and Barriers to Use of the ASMAN Provider-Facing Electronic Platform for Peripartum Care in Public Facilities in Madhya Pradesh and Rajasthan, India | ASMAN was user friendly and required minimal training for providers using it. It was effective in reducing workload in places where manual entry in registers was discontinued. | Poor internet connectivity, tablet malfunction and software glitches caused the greatest difficulties in using the ASMAN digital platform. Offline data entry suffered from occasional bugs, resulting in double entries or data loss when synced online. Database inflexibility affected some respondents: providers reported being unable to change incorrect entries using the tablet. At facilities that continued to use written registers respondents reported increased workload due to double documentation. It was difficult to enter data while providing patient care. Urgent patient needs often delayed data entry, especially amongst staff nurses. Language also acted as a barrier as there was no hindi version of the application available. Participants also complained of lack of awareness of all application features. | The intervention improved job performance, facilitated better communication, management of cases, improved reporting processes, reduced human error, facilitated timely referrals, increased confidence in clinical skills among staff. It also reduced transition time between shifts and improved continuity of care. It improved patient outcomes - decrease in obstetric complications and stillbirth rates and increase in timely referrals. | Study limitations include the categorization of facilities into “high-utilization” and “low-utilization” groups based on a proxy indicator of data field completion rates because other aspects of utilization were not quantifiable. Data quality was questionable as interviews were conducted remotely and due to social desirability bias. Lastly, the study evaluated perceptions rather than actual use behavior without assessment of ASMAN on the quality of care provided or on subsequent maternal and neonatal outcomes. |
| Muke et al (21) | 2020 | Quantitative (Randomised Controlled Trial(RCT) | Madhya Pradesh | Non specialist Health Workers | 36 | To assess the feasibility and acceptability of digital training programs and changes in competency outcomes, using a self-reported measure covering the specific knowledge and skills required to deliver the brief psychological treatment for depression. | Prior training improevd feasibility, acceptability, and adoption of the intervention. Attractive, interactive, and well-designed application also improved its acceptability. The app made learning flexibile. | Technical challenges - internet related, smartphone hardware/software related, course navigation related, application related, and smartphone handling related. Participants reported challenges in understanding the application and using smartphone. As the application was in English, the language also was a barrier in its understanding. | The intervention improved knowledge of participants who felt happy and good about having the chance of being part of training on mental health and depression. The application helped in improving access to mental healthcare. | The sample size was small and not adequately powered to detect differences in competency outcomes between groups. Desirability bias could have impacted results. |
| Scott et al (22) | 2021 | Qualitative (cross-sectional study) | Madhya Pradesh, Rajasthan | Frontline Health Workers, data entry operators, higher level officials | 59 | To understand factors underpinning the accuracy and timeliness of mobile phone numbers and other health information captured in India’s government registry for pregnant and postpartum women. | Access to mobile phones in the women and having trust in the ASHA acted as facilitators. | The intervention resulted in increase in workload. Low-literacy, slow internet connection or server capacity, lack of functioning battery backup, infrequent and inadequate training, software design issues, shortage of human resources, and shortage of time due to patient care acted as barriers in effective implementation of the intervention. | Better recording, better monitoring of service provision by FLHW and planning of resources. | - |
| Bhattacharjya et al (23) | 2022 | Randomized Controlled Trial(RCT) | Not available | Community-based Rehabilitation(CBR) workers | 12 each group | To assess the usefulness of an mHealth strategy to support assimilation of adaptive feeding interventions into daily practices of community-based rehabilitation (CBR) workers. | A 4-week, mixed-methods field test was conducted. All participants received a 1-hour training session at baseline, which introduced them to adaptive feeding interventions. In addition, CBR workers in the intervention group received smartphones pre-loaded with the video training modules, which could be reviewed on-demand throughout the 4-week intervention period. Both groups were instructed to apply the adaptive feeding techniques as appropriate over a 4-week period with families having a child with CP. Compared to the control group, the intervention group visited approximately 2.4 times the number of families having a child with CP with feeding difficulties and shared approximately 2.4 times the number of intervention strategies. Over the four weekly reporting calls, six control group participants sought advice regarding customized interventions for individual families – e.g., building a corner seat, or modifying a lap table – or sought verification from the PI that a specific recommendation to caregivers was appropriate. None of the participants in the intervention group sought intervention advice or support during the weekly check-in calls. In the post-study questionnaire, 91% (n = 10/11) of the intervention group indicated either strong or moderate agreement that the strategies were helpful; whereas, 60% (n = 6/10) of the control group reported strong or moderate agreement that the strategies were helpful. Regarding the ease with which the strategies could be assimilated into daily practice, 82% (n = 9/11) of the intervention group strongly agreed that the techniques were easy to apply in the field; in contrast, 60% (6/ 10) of the control group expressed moderate agreement that the techniques were easy to apply in the field : (a) Both groups felt that the training content had substantially improved their knowledge base of adaptive feeding strategies that were relevant to their caseloads; (b) For the intervention group, the smartphone platform was easily assimilated into practice; and (c) For the intervention group, on-demand access to the video modules improved their effectiveness as service providers. CBR workers in both groups repeatedly described how much they valued the intervention strategies they had learned. Intervention group members specifically emphasized three production features: (a) the modular nature of the videos was efficient to demonstrate only relevant content with families; (b) The video modules provided a helpful mix of still photographs and videos, simplifying explanations to caregivers; (c) Video narration in Bengali was easy to understand for workers and most caregivers. Hindi narration would have helped those caregivers who had relocated to West Bengal from another state and were not fluent in Bengali. For these families, the CBR workers translated the narration to Hindi. The intervention group members also discussed the usefulness and usability of the technical platform: (a) Bluetooth connectivity allowed CBR workers to share video modules directly with families having compatible devices in environments that lacked WiFi network access; (b) the smartphone platform was useful for capturing video documentation of children, enabling CBR workers to seek treatment advice from colleagues; The battery capacity allowed the smartphones to operate for 2 to 3 days in between charges. . Reviewing video segments enabled them to recall forgotten details, clarify ambiguities, and reinforce their existing knowledge base; (b) Clarity & specificity of intervention recommendations. The videos reinforced CBR worker suggestions for caregivers, answered caregiver questions, and decreased the potential for miscommunication; (c) Psychosocial outlook. The videos provided external affirmation that enhanced CBR worker self-confidence and increased their credibility with caregivers. Capturing photo or video of a child at an initial visit and then reviewing it with the caregiver at a later date to assess a child’s progress; (b) Capturing video of challenging cases so they could seek advice from colleagues; and (c) Graded introduction of the strategies over time to allow a child to become accustomed to the changes being asked of them All participants strongly agreed they would recommend the videos to their peers. | The small screen size and limited speaker volume were not effective for demonstrating the video modules to large families. (e) Intervention group members having a personal phone felt the burden of carrying the study phone in addition to their personal phone. (f) Others acknowledged the frustration of periodically forgetting to charge the phone. (g) Some participants encountered technical obstacles that they resolved independently without need for assistance from the study team. Control group: Control group members described their difficulty remembering details of the training content over the 4-week study period. They expressed uncertainty and lack of confidence about the correctness of their adaptive mealtime suggestions to caregivers, and felt that their explanations to families were insufficient. ” Several lamented not having continued access to the videos, which would have enabled recall of mealtime strategies with greater certainty and clarity for families. | y. mHealth strategies are a promising option for supporting geographically dispersed CBR workers implementing multi-faceted assistive technology interventions. The portability of video content reinforced learning, increased implementation of the adaptive feeding interventions, and enhanced communication with consumer. | The scale of data collection was intentionally constrained for two reasons: (a) as a relatively uncontrolled real-world test of the smartphone-based videos, it did not make sense to conduct a larger scale evaluation; and (b) given project resource constraints and time-intensive nature of the weekly phone calls, data collection with additional participants would have been logistically challenging to sustain. The possibility of response bias is another limitation of the study. Although there is potential for researcher bias in the qualitative analysis, this was minimized through independent analysis by two reviewers, as well as the use of complementary quantitative data collection tools. |
| Gupta et al (24) | 2021 | Mixed-Methods (Cross- sectional Study) | Himachal Pradesh | Auxillary Nurse Midwives, Medical Officers | 20 | To assess a digital health model for the early identification and line listing of high-risk pregnant women (PW) with appropriate referrals and increased engagement with the healthcare workers. | The intervention helped ASHA in correctly identifying high risk pregnancy and automatically send msg to pregnant women even if the ASHA forgets. The intervention did not increase their burden. An overall mean score of 4.5 was obtained, indicating that satisfaction with the App was on the higher side. | Poor connectivity, the app hanging up, inability to enter old data in the app and/or failure of app tp sync data with the existing portal acted as barriers. Lack of availability of suitable and updated hardware with the internet connectivity to the health worker and login credential being health‑worker specific became a challenge in data entry. | The SEWA application is a feasible and sustainable solution to complement the competency of the care providers for the early identification of high‑risk conditions. | - |
| Charanthimath et al (25) | 2021 | Mixed-Methods (Cross- sectional Study) | Karnataka | Community health workers(ASHAs, ANMs) | 48 | To evaluate the experiences of CHWs of using PIERS on the Move (POM) in rural India and their perceptions of acceptability and feasibility of this mHealth intervention. | The qualitative analysis highlighted that health workers who used POM reported improved interactions with women and families in their communities. POM strengthened the role of ASHA as a CHW beyond a “link-worker” accompanying women to health services. With training, the mHealth application was easy to use even for CHWs who did not have much experience with smartphones. E-training to use the mHealth application and skills required to care for pregnant women were effective. Health workers largely found the POM app easy to use, especially after the training was conducted for both ASHAs and ANMs in their local language of Kannada. Health workers shared that they felt confident using the POM app, and it helped them in identifying and advising women about pregnancy complications and guiding the management of conditions. Use of pictograms to illustrate symptoms were well-received by health workers and community members. The health workers were supported by the research project team when there were technological difficulties which improved the ease of usage of the intevention. | Relatively few health workers owned a smartphone prior to the study. Complexity of editing data and the length of the application with many app fields to fill in acted as barriers. Phone-related issues like phone date often getting reset, small size of the phone, and insufficient battery life, and application-related issues like trouble with synchronization, difficulty in entering information and changing dates, lack of more local languages, along with difficulty understanding how to ID women at first were also reported as barriers. | The intervention can strengthen health systems to reach rural, remote, and marginalized populations to reduce disparities in health. | Desirability bias could have affected the findings. Culture differences could have resulted in a better response in the likert scale. |
| Srinidhi et al (13) | 2021 | Qualitative (cohort study) | Karnataka | ASHAs | 16 | To describe the experience of implementing accredited social health activists (ASHA) Kirana, a digital technology-enabled Maternal Clinical Assessment Tool (M-CAT) and describe how the ASHAs felt empowered in the process. | It oriented ASHAs to the idea that maternal risks require prompt action and provided doctors with casesheets functioning like protocols to prompt systematic clinical assessments. Following initial training, the ASHAs were provided practical field-based training and support. Sheer practice also helped them elicit symptoms expertly. By the end of the fourth month, the ASHAs reported that the M-CAT had transformed their interactions with women and families. Repeated use of the forms reinforced their training and provided them with an opportunity to learn something new, while giving them the confidence to ride on their freshly acquired skills. The ASHAs additionally felt that greater familiarity with risk conditions had implications for their status within the community and made them feel empowered by knowing more. | Lack of experience and low literacy acted as barriers initially. ASHAs were also unsure about how women and their families would respond to the forms, because of the popular belief that surveys only benefit external organisations. The tool itself was preliminary in nature, paper-based, and faced challenges in real-time diagnosis and tracking. For example, while data on symptoms and signs pertained to the current month, laboratory test results were usually carried over from previous months, Thus an accurate current diagnosis was not possible in some cases. Additionally, when ASHAs and/or women went directly to a higher centre outside the implementation geography, the information was not communicated to the PHC Medical Officer. | Technology-driven initiatives that include empowerment as an objective can strengthen the role of front-line workers in health systems. | - |
| Bairapareddy et al (26) | 2021 | Quantitative (Cross-sectional study) | Not available | primary respiratory care physicians, rehabilitation nurses,physical therapists and respiratory therapists | 52 | To assess the feasibility of administering a smartphone-based telerehabilitation program for chronic obstructive pulmonary disease patients in India. | The majority of the healthcare professionals were found to be aware of smartphone-based telerehabilitation benefits and the overview of the functioning. We found fair amount of awareness about inclusion criteria, implementation and safety regarding telerehabilitation COPD. The majority of the participants perceived smartphones and video conferencing software to be predominant ingredients for the successful implementation of smartphone-based telerehabilitation services while the need for web cameras to be the least. The majority of respondents perceived the need for specialized training in the implementation of smartphone-based telerehabilitation programs, while few were confident in implementing PR, monitoring outcomes, and progression through information technology. Majority of the healthcare professionals supported hospital initiated telerehabilitation services followed by smartphone-based telerehabilitation training programs for healthcare professionals. Further patients found smartphone-based telerehabilitation for COPD as more comfortable and suitable. | Poor health literacy, financial burden, lack of external reward or intrinsic motivation to administer remote PR programs, lack of technology/availability of resources and lack of expertise were identified to be the predominant barriers to the successful implementation of the smartphone-based telerehabilitation programs for COPD patients. Only 49% expressed their interest in conducting research on telerehabilitation and its efficacy on COPD patients in India. | Telerehabilitation through smartphone is an innovative management strategy that addresses the disparity in healthcare utilization between rural and urban areas. | Sampling technique, homogenity of sample, and significant age difference between the two study groups were identified as limitations of this study. The potential barriers and facilitators for patients to utilize telerehabilitation were framed from the practitioner's background rather than the results of focused interviews among the patients. This might have caused some missed barriers from the patient's aspect. |
| Bashingwa et al (27) | 2021 | Quantitative (cohort study) | Assam, Bihar, Chhattisgarh, Delhi,Haryana, Himachal Pradesh,Jharkhand, Madhya Pradesh, Odisha, Rajasthan, Uttar Pradesh, Uttarakhand, West Bengal(Bihar, Uttar Pradesh and Odisha were excluded from the analysis) | ASHAs | not mentioned | To examine the reach and exposure of a mobile phone-based training programme for frontline health workers (ASHAs) in 13 states across India. | Mobile Academy completion rates for ASHAs who initiate the course were high, with an estimated 81% (128 135) of ASHAs who initiated the national version completing the course. Additionally, 11% of ASHAs who were initially anonymous users of Mobile Academy later registered their mobile numbers in the government’s databases to complete Mobile Academy. ASHAs who completed Mobile Academy, 99.7% (127 801) passed it. Once initiated, 36% of ASHAs completed the programme within 3 days; 12% within the first 24 hours. ASHAs spent an average of 5 hours over 10 calls for the first completion. Very few (1.9%) of ASHAs completed the course in one single long session, while one-third (30%) completed the course in more than 15 calls. Since January 2016 (when the database started tracking repeat attempts), 17% of ASHAs (21 225) completed the course more than once. Among ASHAs who completed Mobile Academy the first time and attained perfect (44/44) or near-perfect (43/44) scores, 21% completed the course in less than 240 min (cumulative) and 15% spent between 240 and 260 min. Overall, nearly two-thirds of ASHAs (72%) completed the course the first time with a score of 43 or 44. 15% of ASHAs spent between 240 and 260 min to complete the course and obtained a perfect or near perfect score (43 or 44 out of 44). | Nil mentioned | Understanding how ASHAs engaged with the service—defined in terms of completion, time to completion and performance on quizzes—seeks to inform future design efforts. | - |
| Deo and Singh (28) | 2021 | Quantitative (retrospective cohort study) | Telangana | CHWs | number of HR cadre not given | To assess the effectiveness of community health workers (CHWs)-led, technology-enabled programme as a large-scale, real-world solution for screening and long-term management of diabetes and hypertension | The intervention led to an improvement in patient outcome. The findings of the study demonstrate the feasibility of employing lay health workers with minimal training in screening at-risk participants and in reducing blood pressure and blood sugar level for participants who remained enrolled in the study. | The programme faced difficulties in retaining enrolled participants over longer durations, which could limit its impact on health outcomes as well as its financial sustainability. | CHW-led, technology-enabled private sector interventions can feasibly screen individuals for non-communicable diseases and effectively manage those who continue on the programme in the long run. | The study findings were not compared with a control group to obtain the incremental effect of the intervention. The determinants of programme sustainability, that is, programme cost and participant cost incurred and averted were not analysed. Long-term effect of the programme on dropped-out participants was not assessed. |
| Singh et al (29) | 2021 | Qualitative (cross-sectional study) | Delhi, Karnataka, Madhya Pradesh, Gujarat | Technology partners, implementers and technical partners, senior government stakeholders who had played key roles in commissioning, scaling and/sustaining the digital tools, funders and evaluators/academics | 19 key informants | To explore the factors underpinning scale-up of digital health solutions for FLHWs in India, and the potential implications of these factors for sustainability | The intervention was valued as an affordable, standardised and logistically simple mechanism to refresh Accredited Social Health Activist’s knowledge gained from face to face trainings and fill in knowledge gaps. The intervention streamlined data collection systems and improved the timeliness, quality, accessibility and use of data. Most key informants identified adaptability as a key enabling feature for scaling up, but one that was bounded by certain technical nuances. Although respondents agreed that data storage and data governance domains (access, privacy and consent) are key considerations for any digital tool, they did not cite them as a key factor contributing to scale or sustainability. | In order to make changes to hardcoded software a senior engineer is needed to manually rewrite the code; a process which can be time consuming and expensive. Low prioritisation of data governance domains had resulted in poor consent processes at the front lines, wherein FLHWs were ‘not really’ taking proper informed consent when collecting data using digital tools (KI10, government). Another respondent worried that failure to invest in consent processes could someday undermine sustainability by undermining public trust. Procurement, whether of handsets, maintenance contracts, server capacity, or platform services, emerged as a major challenge during the implementation process with implications for long-term sustainability. Donor commitment was key to scaling up digital tools for FLHWs because there is an ‘expensive curve’ at the beginning. India’s federated structure generates a tension between national- and state-level decision-making, ownership and control was alsoa challenge. The challenge of interoperability between digital tools within the MoHFW is enormous, but even if they all aligned with the underlying RCH platform, there are multiple other ministries building digital tools that are not interoperable with RCH. | The use of digital tools by FLHWs offers much promise for improving service delivery and health outcomes in India. | FLHWs were not included among respondents. Some components of the framework were not evenly probed in each interview due to lack of time, which may have hindered the comparability of the digital tools studied. The COVID-19 pandemic impacted the availability of some identified respondents, including additional donors, to be interviewed, which led to a smaller sample size in the study than anticipated, potentially reducing the robustness of the case studies. |
| Schierhout et al (30) | 2021 | Mixed-Methods (Cluster Randomised Control Trial) | Andhra Pradesh | ASHAs | 26 | To identify variation in outcomes and implementation of SMARTHealth India, a cluster randomised trial of an ASHA-managed digitally enabled primary healthcare (PHC) service strengthening strategy for CVD risk management, and to explain how and in what contexts the intervention was effective. | ASHAs’ delivery of screening services was consistently high with 84% population coverage or higher in all PHCs and they followed up 64%–99% (overall 85%) of those identified as at high CVD risk at least once. Support by government PHC providers for ASHAs’ new roles, trust and acceptability from the community were indentified to be important for community uptake of the intervention. Experienced ASHAs with strong community connections, and trust in ASHAs’ new roles promotes uptake of the new service, resulting in stronger community connections | Nil mentioned | Strategies for strengthening digital health enabled PHC service strategy interventions in India and elsewhere. | Period of implementation was short and themes identified potentially overemphasised the perspective of ASHAs as fewer interviews with patients and PHC doctors were conducted. The authors report that they were unlikely to identify all possible influences on cluster-level outcomes— especially if respondents were unaware of them. |
| Subramanian et al (31) | 2021 | Quantitative (Randomised Controlled Trial(RCT) | Kerala | Physicians | 125 | To educate physicians on cancer screening using a mobile application and test the acceptability, utility, and cost of two different approaches to recruit physicians. | Overall, 95.3% indicated that they would recommend the app to others, and 97.6% found the app to be a “very effective” or “effective” approach to educate physicians. In general, physicians who completed the course exhibited improvement in knowledge related to cervical and breast cancer screening. Participants provided overwhelmingly positive feedback on their experience learning via the M-OncoED app. | Slow loading of pages leading to delays, lack of videos and pictoral representation and poor instructions to use application features were perceived as barriers. | The M-OncoED platform could be an ideal tool to delivery education content to busy physicians who do not have long periods of uninterrupted time. | Only short term follow up was done and long term retention of knowledge was not assessed. Only an Android-compatible app was created, as the majority of targeted users had an Android-based phone. In the future, to allow equal participation by all eligible physicians, the app should be available for iPhone users as well. |
| Khan et al (32) | 2020 | Qualitative (cohort study) | Madhya Pradesh | Non specialist Health Workers | 19. These participants ranged in age from 24 to 49 years, with 11 (58%) having education above 8th standard, and 13 (68%) having prior experience using a smartphone. | To describe the systematic approach to designing a digital program for training non-specialist health workers to deliver an evidence-based brief psychological treatment for depression, called the Healthy Activity Program, in primary care in rural India. | The content could be accessed through the mobile app without requiring an Internet connection. Downloading of the course package onto the smartphone was enabled so that the training content could be accessed offline as well. A more comprehensive overview of the digital technology was included in the pre-training session. A protocol for remote coaching support was designed based on feedback from the participants to sustain their engagement in learning the content and successfully completing the digital training. | Technical challenges were reported due to poor connectivity. The participants unanimously expressed frustration with the slow download speed of the videos and accessing the training content. Learning to use the smartphone app for first time users was difficult. Need for additional support to finish the training was indentified. | This study illustrates a step-wise approach to combine evidence-based content with iterative feedback from stakeholders to develop a digital training program tailored to the context in a low-resource setting | The sample size was small, and the convenience sampling approach may have introduced selection bias. The research team influenced the development of the training materials and digital content and may have introduced personal biases during the design of the program. |
| Usmanova et al (33) | 2021 | Quantitative (Cross-sectional study) | Madhya Pradesh, Rajasthan | Medical Officers, Staff Nurses, ANMS | 81 public facilities. | To understand the uptake of ASMAN application and the role of CDSS in improving adherence to key clinical practices and delivery outcomes. | It was found that filling ratio was low in high delivery load facilities compare to low and medium delivery load facilities. However, filling ratio was not statistically significant by facility type and by delivery load. In univariate analysis, statistically significant improvement in adherence to key clinical practices was observed in 18 of 20 key clinical practices. There was a steady decline through the period in still birth rates and the difference between mean still birth rate of four quarters before intervention roll out and four quarters after intervention roll out, was statistically significant. The interrupted time series regression analysis on monthly data revealed that there was a decrease in number of neonatal asphyxia cases after intervention roll out. There was an increase in identification of pre-eclampsia and eclampsia cases after application roll-out, while referral out declined over the same time period. Analysis of postpartum haemorrhage cases and refer out trends in intervention sites before and after ASMAN application rollout revealed an increase in identification of postpartum haemorrhage cases and decrease in referral out. | Nil mentioned | Our study indicates that the digital intervention has a potential to improve quality of intrapartum care and delivery outcome. | Poor recording of the practices was indentified as a possible limitation. The assessment involved direct observation of provider practices, as a result potential Hawthorne effect would have happened. Lastly, due to utilization of program data in the analysis, establishing causal claim was limited. |
| Anand et al (34) | 2021 | Quantitative (cohort study) | Not available | Nurses | 4512 nurses from India | To describe an innovative interactive e-learning method to disseminate knowledge to larger group of participants over a wide geographical area. | Unlike website-based e-learning, the current method provides opportunity for interactive discussions between the faculty and participants. They expressed that the topics were relevant, course was engaging and coordination was satisfactory. The authors could retain the attention and interest of the participants till the end of 10 wk, there were more than 1000 chats and discussions till the very last day of the online course. Majority rated their overall experience as very good to excellent.. | Internet connectivity issues, lack of face-to-face interactions, and language barriers were identified as barriers. ‘Skill transfer’ and ‘skill assessment’ does remain a potential challenge in this platform, like in other online teaching methods. Few participants found it difficult to navigate through the quizbot assessments. | The authors envisage that this course methodology has a potential to reach out to a larger group of learners. | nil mentioned |
| Swathi et al (35) | 2020 | Quantitative (Cross-sectional study) | Karnataka | Doctors, nurses | 53 clinicians and 12 staff nurses | To evaluate a user‑friendly, technically less demanding, mobile App for health‑care professionals, which is accessible even without internet facility. | The evaluation of App was conducted among physicians and nursing staffs in a tertiary care referral hospital. All the study participants expressed good attitude regarding the App. | Nil mentioned | The developed App is a user‑friendly, easily accessible platform, which can help health‑care professionals in making decisions regarding rabies wound management, treatment, and prophylaxis. | In the present study, due to time as well as financial constraints, the study team had to opt for asynchronous testing. Another limitation was the absence of push alert notification function in the App. |
| Ward et al (36) | 2020 | Quantitative (Cross-sectional study) | Bihar | child bearing women | NA | To evaluate the impact of an mHealth tool implemented at scale as part of the statewide reproductive, maternal, newborn and child health and nutrition (RMNCHN) program in Bihar, India. | Multiple health behaviours were significantly higher in those who were exposed to Mobile Kunji as compared to those who were unexposed, including odds of pregnancy registration, birth preparedness activities such as saving money for delivery and arranging transport to the facility, receipt and consumption of IFA tablets, as well as immediate breastfeeding and exclusive breastfeeding. These results were similar for those who had been exposed to the audio component alone, as well as those who had been exposed to both audio and the cards. Trust in healthcare workers was generally higher among those exposed to Mobile Kunji. Similarly, exposed women were significantly more likely to “completely agree” with the information given by their FLW compared to those who were unexposed. The duration of interaction with a FLW was almost 10 minutes longer per visit for the women who were exposed to Mobile Kunji (mean 21 minutes vs 13 minutes, P<0.001), and a significantly higher percentage of women reported discussing the information they received with someone else, often including other family members. | lack of access and literacy | These results can help inform global understanding of how best to use mHealth tools, for whom, and in what contexts. | All three evaluations were survey-based and relied upon self-reported exposure to the interventions. However, the level of exposure and impact among those who were exposed to Mobile Kunji may have been limited by the relatively short, two-year time period for implementation spanned by this survey. The surveys had limited sample sizes for those exposed to certain messages, and thus, detecting meaningful differences in the recall of some specific messages was limited. The results may have been affected by social desirability and response biases. An important overarching limitation of this study is that the results of these surveys cannot delineate whether Mobile Kunji was used alongside other interventions given the context of a complex program implemented through various delivery platforms. Additionally, there may have been unaccounted for clustering of health behaviours among communities and selection bias when FLWs chose those beneficiaries for whom they would utilise mHealth tools. Finally, because intensive support and facilitation were provided for FLWs during the implementation period, generalisability may be limited given the challenges for sustainability and scalability of interventions bolstered by such extensive support. |
| Chattopadhyay et al (37) | 2020 | Qualitative (cross-sectional study) | Not available | Primary Care Physicians | 12 | To test the feasibility of using a set of free web‑based services in digitization, preservation, and retrieval of prescription on a smartphone by primary care physicians. | The intervention was found to be an easy method of prescription digitization and retrieval. All participants liked the system as it is a free service. Ease of uploading data with some clicks on the smartphone also partially eliminates the requirement of a data entry operator. All the components – creating from, data entry, and retrieval can be done on a smartphone. A total of 9 participants opined its advantage of being a smartphone‑based system. This eliminates carrying a personal computer to all the chambers. Rest 3 participants think that a laptop computer with a camera may also be tried if data is entered by physicians’ assistants. The lucid presentation of the tutorial helped the participant, which was praised by 6 participants. The rest 6 participants, though agreed that the presentation was lucid but opined that a video tutorial could help them more. However, one participant requested to add a segment on how to remove a question. | Low storage space on Google Drive was pointed as an issue. Six participants would digitize only important prescriptions to save space. Data privacy concerns were raised as the medical records are confidential data. Two participants thought the time spent in uploading the prescription to be an important inhibiting factor. However, the rest of the participants were ready to invest the time. | Free web‑based and smartphone applications can be used by a primary care physician for personal storage and retrieval of prescriptions. The simple tutorial presented in this article would help many primary care physicians in resource-limited settings. | nil mentioned |
| Harding et al (38) | 2020 | Mixed methods (Quantitative longitudinal study and Qualitative cross-sectional study) | Not available | caregivers | 149 | To design a mobile phone application to enable or improve communication between family caregivers, community caregivers, and palliative care teams; to evaluate its acceptability, processes, and mechanisms of action; and to propose refinements. | The intervention led to an improvement of symptom assessment, better prioritization of patients, and improvement in communication. It facilitated patient review during clinical team meetings and helped in recognition of trends in each patient’s outcomes. It led to strengthening of the voice of the patient and introduction of accountability in community care giving. The intervention can save potential future cost through prioritization and improved timeliness of interventions. All participants described positive experiences of using the app. It led to better understanding of patients’ symptoms, concerns, and facilitated outcomes of care in ‘‘real time’’ with regular ongoing assessment. This information enabled individualized care and was easy to use and understand. It enabled integrated patient-reported outcome assessment. People-centred outcome measurement empowered family and community caregivers to ask the most relevant questions and thereby improve the morale of patients and caregivers. The availability of outcome data enabled staff to prioritize patients. | A key challenge was the ability to learn everything needed within the time limitations of training, although over time users provided peer support to use the app efficiently. In India, there was concern that home visits would be replaced by the app, and also concerns regarding data privacy. Reliance on internet connectivity, which is unreliable in some places, disappointment by patients who expected to receive treatment immediately, requirement that the caregivers be literate, and lack of confirmation for caregivers that their upload was received were identified as barriers. | The app thereby would contribute to implementation of the 2014 WHA Resolution on Palliative Care and to universal health coverage. | The findings lack generalizability due to small sample size and selection bias in the pilot sites (i.e., those willing to pilot the app) may mean that other palliative care teams may find this app less useful. This bias may also have prevented from identifying different challenges to use between stakeholder groups. |
| Modi et al (39) | 2020 | Quantitative (Cluster randomized trial) | Gujarat | Not available | 561 | To assess the incremental cost per life-years saved as a result of the ImTeCHO intervention as compared to routine maternal, neonatal, and child health care programs. | More than three-quarters (76%) of the cost was directed toward annual recurrent implementation costs, comprising personnel, training, software development, annual maintenance, supportive supervision, and monitoring costs. There was a reduction of 16% infant deaths per-protocol in the study area. This resulted in an increase in 735 life years, with a life expectancy of 68.35 years. The implementation of ImTeCHO resulted in saving 11 infant deaths per 1000 live births in the study area at an annual incremental cost of US $54,360 per 1000 live births. Overall, ImTeCHO is a costeffective intervention from a program perspective at an incremental cost of US $74 per life years saved or US $5057 per death averted . | - | ImTeCHO, as a mobile phone app in the hands of health workers, has potential to bridge an important gap in the delivery of existing public health programs through ensuring data entry at the point of service delivery by the health provider through a handheld device, thereby ensuring better quality of data. | A limitation of the analysis is that it did not assess the health care input cost or time spent by health workers in training, supportive supervision by medical officers, and other supervisors from the health system. As the program operated in a realistic environment, it was assumed that the health care input cost will be absorbed as part of the health budget for replication of the program. |
| Shah et al (40) | 2019 | Mixed-Methods (cross-sectional study and interviews) | Gujarat | ASHAs | 15 | To assess the uptake, feasibility and effectiveness of an mHealth intervention in improving the performance of village-based frontline workers, called accredited social health activists (ASHAs), to increase the coverage of maternal, newborn and child health services in rural India. | The coverage of maternal, newborn and child health services largely improved during the antenatal, postpartum and early infancy periods due to the intervention; however, there was no significant difference in the intranatal services. All ASHAs demonstrated sufficient competency to use the ImTeCHO application. None of the ASHAs stopped using the ImTeCHO mobile phone application throughout the project. There was a significant improvement in careseeking from ASHAs for postnatal maternal care (OR 16.0, CI 1.54–166.05) and postnatal neonatal care. Most of the ASHAs agreed that the ImTeCHO application supported them in their work, and did not add components to the ones they were expected to collect before the use of ImTeCHO. The ImTeCHO application made home visitations considerably systematic for the ASHAs and improved their functioning. They found the content of the videos to be culturally acceptable, and the brief videos (2–3 minutes long) easy to use and time saving. They felt that the credibility of the information was increased because it was provided by the video in the voice of a doctor. None of the beneficiaries displayed or expressed discomfort with the presence of the mobile phone. | However, some of the ASHAs felt that their workload increased with the mobile application. The ASHAs found the ImTeCHO’s childcare schedule (children older than 2 months) to be more intensive than what they would like, especially because this was a non-incentivized task. | - | Lack of baseline data, a relatively small sample size, and the fact that effectiveness observed in this study might not be only attributed to ImTeCHO considering the managerial inputs from SEWA Rural were identified as limitations. |
| Suryavanshi et al (41) | 2020 | Mixed-Methods (Cross-sectional and in-depth interviews) | Maharashtra | out reach workers (ORWs) and patients. | 15 ORWs and 15 patients | To guide scale-up and optimize programmatic implementation, a mixed-methods study conducted was conducted for evaluation of the feasibility and acceptability of this intervention. | The mobile health platform helped ORWs effectively do their job. All of the ORWs reported satisfaction with the mHealth application, despite some initial apprehension in using the tablets. The message prompts were reported to be critical in helping them collect data correctly and reminding them of the information to be conveyed on key PMTCT components. The ORWs felt the videos were useful to the clients and the information was presented in an easy to understand format. Nearly all ORWs mentioned that the videos helped increase the client’s understanding of their condition and best practices for themselves and their children. Having videos in multiple languages was key in enhancing comprehension. The ORWs indicated that the videos reduced the time needed to talk about these issues. The behavioral training increased self-awareness and improved relational skills. The personal empowerment training component, which was a central part of overall behavioral intervention appeared to boost ORWs confidence and helped them improve their communication with their patients. Improved initiation and adherence to ART was noticed. More than 50% mentioned that tablet is feasible and acceptable to show videos and to capture data. Tablets have big screens making it easier to show the videos and likely to be understood by the patients. Videos allowed patients to understand the concepts more clearly and they felt that this could be easily rolled out at the national level. ORWs felt it would not be difficult if proper training is given to them and data is captured in the local language. Some ORWs reported this intervention resulted in a major change in their working style and they found data capture was more systematic and easier. . Women understood the content of PMTCT videos. The remaining five that had access to phones reported that it helped them remember the exact date of their and their babies visit. All five patients mentioned how the SMS component of the intervention improved their adherence as it is used to convey reminders to them that was easy for them. Almost all patients reported the strong support they received from ORWs and how they provided information on how to take care of themselves and their children. | There were minimal issues related to network connectivity and sending out forms. The videos were a primary component of the mHealth support, however, their utility was limited to a single showing as most clients expressed a desire not to see them repeatedly. Patients appeared more comfortable to view the videos at the ART center rather than within their home. Lack of compliance - ORWs felt that sometimes that was due to their social situation (e.g. husband doesn’t agree) or lack of financial resources. A similar number reported issues related to the health system, such as the negative behavior of clinic staff, long distances to the clinic, lack of an accompanying person, shortage of drugs, limited counseling by hospital staff, problems in transferring patients to link ART centers (which are closer to patient’s residence) before 18 months (baby’s) influenced ART adherence. Videos helped address misinformation on infant feeding. Of the sample of 15 women, two-thirds [10] reported challenges in access; two had no phones, one was illiterate so did not see any messages and seven of the patients reported that they did not have their phone but gave their husbands phone numbers. Only two said that their husband used to tell them regularly about reminder messages about the visits/ missed visits. | The intervention can be optimized and scaled for India. | - |
| Carmichael et al (42) | 2019 | Quantitative (Cluster randomized trial) | Bihar | Front Line Workers(FLWs) - ASHAs and AWWs and beneficiaries | 1100 Front Line Workers(FLWs) and 3000 beneficiaries | This paper evaluates the impact of a novel mHealth tool that was implemented in Bihar, one of India’s poorest and most populous states which relies heavily on FLWs to provide community-level reproductive, maternal, newborn and child health and nutrition (RMNCHN)-related services | About 85% of ASHAs and AWWs indicated that their ICT-CCS phone was charged and working all or most of the time and three-fourths of FLWs indicated that they had no problem in using the phone. Results for measures related to job confidence suggested that overall confidence and job performance was higher among FLWs from intervention than control villages but was only significant for ASHAs . | Fourteen percent of FLWs said their phone was broken at some time . | The ICT-CCS tool shows promise for facilitating FLW effectiveness in improving RMNCHN behaviors. | A limitation of this study is that some information was not available at both evaluation time points (eg, certain variables collected from FLWs, and maternal reports of frequency of home visits for newborns). In addition, information on actual health outcomes was not available, such as maternal or infant morbidities or mortality; collection of this type of information was not deemed feasible. |
| Modi et al (43) | 2019 | Quantitative (Cluster randomized trial) | Gujarat | ASHAs, type A and B respondents | 578 ASHAs and 3,023 type A respondents and 3,470 type B respondents | To evaluate the effectiveness of a mobilephone–and web-based application, Innovative Mobile-phone Technology for Community Health Operations (ImTeCHO), as a job aid to the government’s Accredited Social Health Activists (ASHAs) and Primary Health Center (PHC) staff to improve coverage of MNCH services in rural tribal communities of Gujarat, India. | Throughout of the study period, uptake and adherence to the ImTeCHO mobile phone application was satisfactory among ASHAs. Job performance, coverage of services and care seeking improved in the intervention group. Additionally, the mHealth strategies provided support to the ASHAs and encouraged them to adhere to protocols. The targeted client communication in the form of short video clips were found to be effective. The uptake of the ImTeCHO intervention was satisfactory among ASHAs as reflected in high login and task completion rate, whereas it was lower than expected among the PHC staff. | The PHC staff and PHC medical officers’ use of ImTeCHO web interface was less than expected. Throughout the study period, 17 mobile phones (285 ASHAs) were lost, stolen, or irreparably damaged. 96 software issues were escalated to and addressed by Argusoft India Ltd over 12 months. As expected, ImTeCHO intervention was not found to be effective towards increasing coverage of MNCH health services in which the ASHA was not the primary provider and relied on other cadres or infrastructure of the health system (such as full ANC, vaccination, or institutional delivery). | The findings support the scale-up of mobile-phone-technology–based interventions as a job aid to frontline health workers to improve health outcomes. | The outcomes were reported by the mothers, and there was a risk of recall bias. There was a higher proportion of women from the scheduled tribe and from the lowest standard of living index in the intervention arm. The total duration of the intervention was 12 months, which might be considered short. |
| Muke et al (44) | 2019 | Qualitative (cross-sectional study) | Madhya Pradesh | ASHAs | 32 | To explore the acceptability and feasibility of using digital technology for training community health workers to deliver evidence-based brief psychological treatment for depression in rural India. | Learning on the digital platform appeared to be a positive experience for several ASHAs. This was reflected when they talked about the value of learning using digital technology as this can give them freedom to learn and re-learn different aspects of the course content at their own pace. The digital training program improved the knowledge of ASHAsand helped them save time and costs that are typically spent on travelling to attend in-person training. The ASHAs also described the convenience of the digital training program, because they would no longer need to spend time away from their family for several days to attend inperson trainings. In each round of prototype testing, the ASHAs accessed the digital training platform and learned to use the platform with ease when they were provided additional instruction about the different features of the device and how to access the content on the device. e. In the evaluation of the second prototype, ASHAs appeared to find the modified language easy to understand and indicated, “it was their own language” | Limited familiarity with using digital devices among ASHAs emerged as one challenge. Several technical issues occurred during the prototype testing, and these were mentioned by ASHAs. These included interrupted and slow internet, slow speed of the devices, and poorly functioning touchscreen. ASHAs mentioned low sound quality as a concern. Some ASHAs also suggested making the course available offline to remove some technical challenges. | These findings can inform use of technology as a tool for developing the clinical skills of community health workers for treating depression in low-resource settings. | The small sample size and limited learning content was a limitation with this study, and thus the findings need to be interpreted cautiously and may not generalize to the larger ASHA workforce from diverse regions across India. The study setting was also a limitation because the prototype testing was conducted in community health centers. This classroom style setting may have been supportive for helping ASHAs learn the content. |
| Khan et al (45) | 2019 | Quantitative (cohort study) | Uttar Pradesh, Bihar | Female trainees | 55 | To explore the feasibility of the Acute Care Providers Project (ACPP) to remotely train community members to be health care providers in 2 sites: Haiti and India. | All trainees passed the procedural performance checklist. Graduates of the program are currently providing basic acute services in both nations | - | The ACPP offers a scalable, replicable asynchronous curriculum to train lay individuals to provide basic health care in rural communities. | - |
| Jindal et al (46) | 2019 | Research based interventions | Tripura | Not available | Not available | To articulate the key strategies used for scaling up a research-based intervention, mPower Heart electronic Clinical Decision Support System (e-CDSS), for state-wide implementation at health facilities in Tripura. | By involving all the concerned health care providers of health systems and defining their responsibilities, it became easier to implement the intervention, as everyone took ownership of his or her task. Nurses knew that the safety of equipment and using mPower Heart e-CDSS was their responsibility, so they asked for all the required help without any hesitation. Formation of a technical-cum-coordination unit helped in not only overcoming the barriers immediately, but also expanding the scope of intervention. Initially, the WhatsApp group only had the NCD nurses, but subsequently, medical officers, district NCD coordinators, SPO, and Mission Director, National Health Mission Tripura adjoined the group. This group provided a platform for notifying all the issues related to NCD activities to senior health officials who provided support to rectify these issues at the earliest. The intervention promoted task shifting and provision of evidence-based care to patients with NCDs. As the nurses were the primary users of the mPower Heart e-CDSS, content and duration of the training was developed to meet the needs of the nurses. All the nurses found the training helpful. Through refresher and on-site training, the project team built the confidence of the health care providers in the new intervention. Also, features such as the graphical display of the patient’s clinical parameters not only helped the doctor in making an informed decision about the patient’s management, but was also appreciated by patients. Nurses felt more confident while providing care to the patients after they started using the technology. Features such as searching patient’s previous records and reminder folder helped the nurses in performing their duties more efficiently. | Nonavailability of manpower (nurse or doctor) and the requirement of additional space or funds were key barriers. Of 40 nurses who attended the training, 16 nurses were using the tablet computer for the first time. Project team faced many challenges while providing an enabling environment, for example, resistance from the registration desk and doctors while implementing new workflow recommended for the mPower Heart e-CDSS and also arranging space for the NCD clinics. | - | - |
| Birur et al (47) | 2022 | Qualitative (cross-sectional) | not mentioned | CHWs and study participants | 2 CHW and 3445 study participants | To assess the use of mHealth by community health workers (CHWs) in the identification of oral mucosal lesions. | The positive oral lesions included 334 (84.8%) true positive oral lesions, false positive in 71 (2.3%) individuals, and false negative in 60 (15.2%) individuals identified by CHWs. There was 96% agreement between CHW and remote specialist in identification of oral lesions with a κ score of 0.62, which showed substantial agreement. A statistically significant association was found between CHW and remote specialist in biopsy-confirmed lesions (χ2-value = 50.33; P value < 0.001). The diagnostic accuracy of a remote specialist was 96.4%. There was 97% agreement between remote specialist and onsite specialist in identification of oral lesions with a κ = 0.94, which had almost perfect agreement. A statistically significant association was found between remote specialist and onsite specialist in identification of oral lesion. | The technical challenge in rural India is poor connectivity and low bandwidth, which poses a challenge in uploading files of large size. Hence the protocol was to capture the altered mucosa and a normal mucosa rather than taking images of entire oral cavity. However, if multiple suspicious lesions were found, all the subsites of oral mucosa were captured. | This module facilitates remote consultation and enables health care services far from clinical setting at remote places. | This study did not assess the agreement between two CHWs as individual CHW performed screening on different subgroups. Thirty-one cases were missing in Open MRS among 3414 uploads, which could be due to voluminous screening and an incorrect entry of alphanumeric IDs; and 36 images were of poor quality due to poor retraction and instability to focus on lesions because of subject movement and/or phone movement, zooming with autoflash. |
| Abdel-All et al (48) | 2019 | Qualitative (Cross-sectional) | Not mentioned | ASHA | 20 ASHAs | This paper describes the development of a DCE for low-literacy community health workers (CHWs) in low-resourced setting on an Android platform | The research team conducting the survey reported that the DCE was well received by the ASHAs and that they did not find it difficult to understand the choice sets presented to them. The ASHAs did not take much time to familiarize themselves with the computer tablets. The observers noticed that the younger ASHAs used the tablets with more ease compared to the older ASHAs. Most ASHAs had never used a computer tablet prior to the study, but almost all of them had access to a smart mobile phone. The Android-based answers were in line with the paperbased tool answers. Data collection time was notably shorter using the computer tablets, (10 to 15 min) compared to (20 to 25 min) for completing the paper based DCE. In addition, data collection time was notably quicker, since no additional data entry or cleaning was required. | not mentioned | It is feasible to use technology to develop and implement DCEs among participants with basic education in resource poor settings | The study did not investigate the capability of the low-literate community health workers to handle the complete DCE experiment without the research team guidance to explain the nature of the experiment. |
| Naslund et al (49) | 2019 | Review | Andhra Pradesh and Maharashtra | hcp s including non specialists | review - 2 Indian studies | To summarize examples and discuss future opportunities to use digital technology for supporting the development of a trained, effective, and sustainable mental health workforce. | Atmiyata champions and mitras were provided smartphones to access videos for ongoing training tools to learn about mental health concepts and to refresh their knowledge and skills. The program also includes Android mobile applications, with one version for the Atmiyata champions and a separate version for the general public and community members. The mobile app contains basic e-learning videos on mental health for the Atmiyata champions with questions built into the app following each training video to test knowledge of mental health topics.  The Systematic Medical Appraisal Referral and Treatment (SMART) Mental Health project involved task-sharing and use of mobile technology to support primary care health workers in providing evidence-based mental health care in remote villages in Andhra Pradesh, India. In this project, community health workers referred to as Accredited Social Health Activists (ASHAs) and primary care doctors were trained to screen, diagnose and manage common mental disorders using an electronic decision support system. At 3-months follow up, there was a reduction in depression measured using the PHQ-9 and a reduction in anxiety measured using the 7-item Generalized Anxiety Disorder (GAD-7) questionnaire. A key finding was that individuals increased their use of mental health services following screening and referral to the primary care doctor. The SMART program supported with mobile technology was considered both feasible and acceptable for use in this rural setting. | - | Ongoing work will be critical to explore new opportunities for digital technology to drive innovative models of care delivery necessary to enhance the impact and reach of non-specialist health workers, to demonstrate the effectiveness and cost effectiveness of these efforts, and to determine the scalability and sustained delivery of digital interventions in low-resource settings. | The search of the literature was not systematic as it was a narrative review expanding on four recent relevant reviews to identify the selection of highly promising examples summarized here. |
| Peiris et al (50) | 2019 | Quantitative (Randomised Controlled Trial) | Andhra Pradesh | Ashas ,physicians | 240 ASHAs, 18 doctors and 8642 paticipants | A stepped-wedge, cluster randomised controlled trial of a community health worker managed mobile health intervention for people assessed at high cardiovascular disease risk in rural India. The primary outcome was the proportion meeting systolic blood pressure (SBP) targets (<140mmHg). | ASHAs and PHC doctors were trained to assess CVD risk using a local (Telugu) and English language clinical decision support system (CDSS) application on a 7-inch Android tablet device. Each ASHA was provided with the necessary equipment. Overall there were no significant differences in any of the primary or secondary outcomes, aside from a small increase in self-reported physical activity. However, there was a significant improvement in use of BP medications (54.3% intervention vs 47.9% control. The mHealth platform was highly effective in linking village based assessments to doctor level care and supported systematic follow-up care in the majority of patients who needed such care. | Not mentioned | The findings clearly demonstrates the potential to leverage the ASHA workforce across India and to expand their role beyond maternal and child health role to non-communicable disease management and prevention. | Only around 50% of the outcome evaluation cohort were exposed to the intervention and many people for whom outcome data were not collected, were exposed to the intervention. A second issue relates to seasonal fluctuation in BP levels and the impact this may have had with a stepped wedge design. A third issue is related to a higher than expected improvement in BP treatment rates in the control period (a 5.6% absolute improvement). This could be related to known phenomena of control groups experiencing improvements when participating in trials but may also be due to background changes to usual practice. |
| Dandge et al (51) | 2019 | Quantitative (pre-post demonstration) | Telengana | Non physician health workers,physicians,ashas,anms | 3 ashas,2 anms,2 supervisor,1 physician | To determine the feasibility and effectiveness of an intervention anchored on mHealth and task sharing strategy of involving non-physician health workers (NPHW) on population level detection, treatment and control of hypertension and diabetes in India. | Overall, 54% of the individuals with hypertension achieved BP control status based on the end of study visit measurements. Based on the end of study visit HbA1c measurements, blood sugar was controlled in 34.0% of all individuals with diabetes. The relatively high participation rate in screening and follow-up demonstrate the acceptability of the intervention strategy at the community level. Identification of NPHWs from the study villages itself may have improved the acceptability of the intervention. The technology enabled and trained NPHW shared the responsibility of screening, risk management, and maintenance of the electronic health records of hypertension and diabetes patients. | Study NPHWs were above average in their capabilities, motivation and performance in the field. Additionally, the intensive in-office and long duration of on-field supervised training are quite demanding for an average ASHA. It is therefore, a challenge to maintain the same quality and standard in scale-up operations of the developed intervention strategy. | This research demonstrates the feasibility and local acceptability of a mHealth intervention strategy anchored on NPHWs guided by physicians for detection, treatment and regular follow-up of individuals with hypertension and diabetes in a community setting in India. | Being a demonstration project on a small scale without a comparison group, the results have limited generalizability. |
| Bhatt et al (52) | 2018 | Mixed-Methods (pre-test/post-test study and focus group discussions) | Mandhya Pradesh,Chattisgarh,Tamil Nadu | 8686 people,community health workers (CHWs - 10 from RUHSA, 8 from Mungeli and 7 from Padhar), supported by nurses (2 in Mungeli and Padhar, 5 in RUHSA) | 34 HCW and 8686 patients screened | To share lessons on use of mobile technology for cancer screening in rural India. | At each of the three sites, CHWs and nurses attended 3-4 hour sessions on the use of the mHealth tool and were trained using a range of methods including demonstrations, role-playing, group activities and practice exercises. Each workshop provided the staff with training materials, including a comprehensive guide for programme coordinators and a step-by-step tool guide for the CHWs and nurses. A pre-test/post-test training assessment was conducted at each site and demonstrated that, at the conclusion of the training, the trained staff were comfortable with the use of the feature phone. Mean follow up of the 34 prototype users (25 CHWs, 9 nurses) was 14 months from the time they received the feature phone and began using it. CHWs and nurses at each site continued, over the course of our evaluation, to actively using the mHealth prototype to report screening, test results, and treatment. Most participants felt the prototype was simple and easy to use; they typically indicated that the prototype promoted efficiency in a variety of ways, including reduced reporting time, less data errors, and enablement of faster case management and follow up in real-time. Younger CHWs and programme coordinators however, expressed a preference for smartphones over feature phones. Most participants reported that the mHealth prototype had a psychological impact effect on them – typically they felt that being provided with a phone, and the associated training, led to an increase in self-confidence, self-esteem, and motivation. Female staff, in particular, typically reported a greater sense of confidence in their own abilities. They also reported positive comments from their patients, who often viewed the mobile tools as a commitment of the hospital to their care and well-being. Health workers reported that the mobile tools made reporting much easier and quicker as compared to paper-based systems which were more cumbersome to carry out in the field and time-consuming to maintain. | Significant barriers to uptake of screening, including social and cultural factors and financial constraints were reported. The CHWs and nurses were disheartened by the impact of the mHealth intervention on take-up of follow-up investigations amongst test-positive individuals. | Education and promotion of cancer awareness are vital accompaniments to future mHealth strategies targeting cancer screening and early diagnosis in low-income countries. | Nil reported |
| Shah et al (53) | 2018 | Quantitative (nested cross-sectional study within a cluster randomised controlled trial) | Gujarat | 45 ashas | 45ASHAS | To evaluate the effectiveness of an mHealth intervention in improving knowledge and skills of accredited social health activists in improving maternal, newborn and child health care in India. | The proportion of ASHAs who identified five pregnancy complications was significantly higher in the intervention compared to the control group. In the intervention clusters, 75% (47 of 63) of ASHAs had knowledge of at least five signs of newborn complications compared to 57% (35 of 61) in the control areas. Both ASHA groups knew about the advantages of kangaroo mother care (KMC), but knowledge regarding the impact of KMC on weight gain was significantly higher among the intervention group. Both groups exhibited similar knowledge about pneumonia symptoms and treatment, of giving oral rehydration solution (ORS) and zinc for diarrhoea, stopping exclusive breastfeeding once weaning is initiated, supplementary feeding practices in infants aged six months to one year. Correct knowledge about giving fruits such as bananas to a child was higher in the intervention compared to the control group. The proportion of ASHAs who washed their hands before checking newborns was significantly higher in the intervention group. ASHAs in the intervention group had significantly better counselling skills regarding supporting the whole body of the newborn and keeping the chin close to the breast while breastfeeding as compared to the control group. The percentage of ASHAs having appropriate skills for preventing hypothermia by following the correct steps for wrapping the newborn (87% vs 72%), and the correct method of recording temperature (66% vs 36%) was higher in the intervention group compared to the control group. | ImTeCHO, the mHealth tool, required an Internet connection at least once daily for login and to check work logs. | Mobile phone technology should be explored further to provide ongoing training to ASHAs. | The intervention did not include all potential mHealth strategies for improving the knowledge and skills of ASHAs. |
| Pant Pai et al (54) | 2018 | Cross-sectional Study | Rural south India | 15 hcps who followed 35 pregnant women each | 15 HCW and 510 pregnant women. | To evaluate the AideSmart! strategy for feasibility, acceptability, preference and impact on maternal care. | The intervention helped in engaging with participants, with evidence-based information in English and Tamil. Feasibility of the Aide intervention strategy was documented at 91% (466/510). Preference for POCT based screening strategy was high at 73% (359/492). 92% (453/491) of participants rated high satisfaction with the strategy. Enhanced communication aided by AideSmart! helped patients promote an active follow-up without financial incentives. Continuous monitoring kept stakeholders engaged at all times which impacted the quality of health service. | - | The strategy could be reverse-innovated to any context to maximise its health impact. | Convenience sampling has a potential for selection bias and observational designs are limited by observed and unobserved confounding bias. |
| Birur et al (55) | 2019 | Quantitative (cohort study) | Not mentioned | population of 3440 | Primary care dentists,frontline workers,oral cancer specialists | This paper summarizes the mobile-Health model, in which Frontline Healthcare providers (FHP) in rural areas were empowered for early detection and connected to specialist through mHealth. | The work flow involved the use of a mobile phone with oral cancer screening software, Sana. 61% of images obtained by FHPs were deemed interpretable; and 45% of these were confirmed to be OPMDs. In the opportunistic cohort, all images captured (100%) were interpretable and all were confirmed as OPMDs. At the community level, trained FHPs are capable of utilizing smart-phone-based technology for primary and secondary prevention of oral cancer. It was also suggested that further training of the FHP with newer technology that overcomes human error needs to be conducted. This was also effective in creating awareness in the screened population at minimal cost. Mobile phones equipped with cameras for documentation as well as the decision supported algorithm, Poi Mapper, were issued to these FHPs. FHPs selected were residents in the community where the screenings were conducted and fluent in that community’s dominant language. The images obtained at workplace settings were better than those obtained from door-to-door screening. | Uploads were challenging in many locations due to poor connectivity. Post-training dropout of FHPs after program initiation caused unexpected delays in running the program. It was a challenge to identify new FHPs and train them in a timely fashion. Reasons for not participating varied from the absence of male members of the family, social stigmas, ‘lack of time’ in workplace settings to their ‘perception of being healthy’ and lack of awareness. Screening within houses added the challenges of lack of clinic-like environment and poor lighting. Our mHealth initiatives were conducted to evaluate the feasibility and effectiveness of mobile application-based diagnosis. Some difficulty was encountered when subjects were not aware of their date of birth, or phone number, or when they lost their ID cards. | - | - |
| Sharma et al (56) | 2018 | Mixed-Methods (Cross-sectional) | - | Nurses | 238 patients | To demonstrate the feasibility and reliability of cervical cancer screening by health workers/nurses using VIA (Visual Inspection under Acetic acid) in rural setting, and assessing the role of smartphone-imaging for continuous training of nurses. | The cervical images for VIA screened women were reviewed by expert. Expert rated 32/180 (17.8%) as positive and 106 (58.9%) negative. Feedback from the nurse and supervisors revealed smooth processes on clinic days without any disruption in the hospital’s routine activities. No untoward incident was reported during the program implementation. The sessions reported satisfactorily on parameters of infrastructure, recording, and human resource studied during each of the sessions. Thereby, suggesting feasibility of implementation. | Due to lost images, poor image quality, and inability to visualize squamo-columnar junction completely, the reviewer was not able to comment on few images. Referral linkage, however, remained challenging and below satisfactory level. | Appropriately trained nurses can reliably conduct screening. Real-time expert feedback might improve reporting. Rigorous awareness activities and on-site treatment can reduce drop-outs. | - |
| Ilozumba et al (57) | 2018 | Mixed-Methods (cross-sectional and interviews) | Jharkhand | Community health worker | Quantitative study - 740 women and 57 CHWs who utilized the intervention)  Qualitative methods - 12 interviews and 4 group discussions with CHWs and 20 interviews and 5 group discussions with pregnant and lactating women and 15 interviews and 2 group discussions with men were conducted. | To assess Mobile for Mothers (MfM), a community health workers (CHW) utilized maternal mHealth intervention and to identify intervention-related and contextual factors, which influence the observed outcomes of a CHW, utilized mHealth intervention. | The provision of support emerged as a facilitator for continued appropriate utilization of the mobile phone by CHWs. Overall CHWs expressed satisfaction with support received. In the survey, 68.4% indicated that MfM had made them more knowledgeable on recommended maternal health practices. 94.7% of CHWs in the survey indicated that MfM helped them work better and 80.7% reported increased confidence. CHWs shared that the mobile phone improved their ability to perform tasks by enlightening them on issues which they were previously ignorant about, such as the number of required ANC visits. They explained that the intervention also improved their ability to explain concepts (such as the need for STI testing) to their community members. MfM also assisted with their recall and delivery of essential information. ANMs reported that they perceived that MfM increased CHWs’ confidence, improved case-management, improved CHWs’ performance (i.e. number of visits and awareness) and made the CHWs’ work easier k. The local government officer mentioned that he received positive reactions from the CHWs about the program, such that community members were now more interested in listening to the information CHWs presented. In the quantitative surveys, 80.7% of CHWs indicated that they gained respect from women and community members because of MfM utilization. 87.7% also indicated that MfM had led to an increase in the village women trusting them and following their directions. However, 13.3% of CHWs younger than 30 reported that women did not trust them or follow their instructions. None of the older CHWs reported problems with distrust of community members. | Language was a frequently mentioned challenge. CHWs older than 30 found it easier to understand the Hindi utilized in the MfM application compared to those younger than 30. However, younger CHWs found using the mobile phone in general to be easier than older CHWs. Some CHWs shared that they experienced problems with the mobile phone itself and the cellular network. Network and internet service disruptions were system level challenges, which affected the CHWs ability to submit collected data. The CHW who admitted to having problems with MfM had the lowest education level recorded–grade 5. A few CHWs (6%) reported that MfM made their work harder; they were all in the younger age bracket. 7.14% of CHWs in the lower education group reported that MfM made their work more difficult. | Mobile health applications are promising interventions for improving the performance of CHWs and health-seeking behavior of pregnant women. | A limitation of this study was that the primary researcher and other research assistants were not local to the research environment. Attempts were made to limit this by engaging a local translator and involving local experts in instrument design and data interpretation. In addition, simple expressions in Hindi and Kortha can have nuanced meanings, which are difficult to capture in English. Also qualitative and qualitative data collection occurred concurrently, so while qualitative interviews provide additional explanations for a number of quantitative results, some points were not explored in the qualitative interviews. |
| Patterson et al (58) | 2017 | Quantitative (cross-sectional) | Chattisgarh | Non physician health worker (NPHW) | 15 NPHWs and 96 patients | To compare the use of an app on a tablet computer to diagnose epileptic episodes NPHWs with diagnosis by local physicians and a neurologist. | The participating NPHWs were given 11 h of training delivered in Hindi which covered the nature of epilepsy, causes, treatments, first aid, education of patients, neurocysticercosis, its effects and social aspects. The app disagreed with the neurologist on eight out of 96 cases (8%) with physicians disagreeing on seven (7%). Of the patients less than age 10 where the app had not been used in the previous studies it agreed with 17 of 19 patients all of whom had epilepsy according to the neurologist. The remaining two had uncertain scores. The physician agreed with all 19. . The neurologist rated 19 as epilepsy and three as non-epilepsy. The physicians judged the diagnosis as uncertain in two and made five misdiagnoses. The NPHW/app had four uncertain diagnoses but only three misdiagnoses. | - | These results suggest that taskshifting epilepsy diagnosis and management from physicians to NPHWs, who are enabled with appropriate technology, can be an effective and safe way of reducing the epilepsy treatment gap. | - |
| Garner et al (59) | 2017 | Quantitative (cross-sectional) | Karnataka | Nurses and physicians | 97 nurses and 17 physicians. | To determine smartphone access and use including future opportunities for mHealth and potential ethical implications among health care professionals practicing at a health care facility in Bengaluru, India. | While 100% of respondents had a mobile phone, 75.3% (N = 73) reported their cell phone was a smart phone and completed the remainder of the survey. Among respondents who had a mobile phone but did not have a smart phone (N = 24), 100% were nurses. Among physician respondents, 52.9% (N = 9) had access to WiFi at work compared with nurses at 37.5% (N = 21). | The participants were had ethical concerns related to mobile phone use.Other concerns included potential confidentiality breach, misuse/ misconstruing of health information, cybercrime, and increase in patient anxiety due to phone use. | Credible, evidence‐based, affordable mobile applications are needed to provide a platform for continuing health education to health professionals and patients in India and limited resource settings | The convenience sample used in this study limits the generalizability of the study results. Additionally, the study initially aimed to target health care professionals which may have included pharmacists, physical therapists, occupational therapists, and others, but the procedure for distributing the survey within the health care facility resulted in all respondents being either nurses or physicians. |
| Prinja et al (60) | 2017 | Qualitative (pre- and post-quasi-experimental design) | Uttar Pradesh | ASHA workers | 259 | To raise the quality of counselling by community health volunteers resulting in improved uptake of maternal, neonatal and child health services (MNCH), an m-health application was introduced under a project named ‘Reducing Maternal and Newborn Deaths (ReMiND)’ in district Kaushambi in India. The aim of the study was to report the impact of this project on coverage of key MNCH services. | The application had tailored content with locally relevant audio and visual prompts to equip ASHAs with multi-media job aids to support client assessment, counselling, early identification, treatment and/or rapid referral of pregnancy, postpartum and new-born complications. The intervention resulted in a statistically significant increase in coverage IFA supplementation (12.58%), identification and self-reporting of illnesses/complication during pregnancy (13.11%) and after delivery (19.6%) in the intervention area vs. the control area. The coverage of ≥3 ANC visits, ≥2 tetanus toxoid, full ANC and ambulance usage also increased in the intervention area by 10.3%, 4.28%, 1.1% and 2.06%, respectively; however, the change was statistically insignificant. | - | The Government of Uttar Pradesh and the Government of India could consider replication of such an intervention in the entire state or country. | Allocation of intervention was not randomised. Also, there may be some individual-level unobserved heterogeneity which cannot be removed even after matching. Another limitation of the matching method was the limited number of variables used for matching. |
| Naslund et al (61) | 2017 | Review | Multiple settings in India | Health care providers | Narrative Review | The study reviewed evidence on the use of mobile, online, and other remote technologies for treatment and prevention of mental disorders in low-income and middle-income countries. | Web based screening tools designed to be delivered on mobile phones by non-medical health workers were effective for diagnosing depression or common psychiatric disorders in clinical settings in India. The review reported digital interventions to be feasible and reliable and consulting psychiatrists and patients reported them to be satisfactory. | - | Digital technologies represent tools that clinicians, researchers, and policy makers must embrace to advance efforts to support and treat individuals with mental health and substance-misuse disorders in lowincome and middle-income countries. | - |
| Maulik et al (62) | 2017 | Mixed-Methods (pre-post evaluation) | Andhra Pradesh | Health care providers | 23 health care providers and 5007 individuals | The project had two key objectives: 1. The development of a multifaceted intervention using training, task shifting, and mobile–based decision support to increase the screening and referral of individuals with CMD in one area of rural Andhra Pradesh. 2. To evaluate the feasibility and acceptability of the intervention amongst community members, health workers, and other stakeholders, and document preliminary evidence and lessons learned about the intervention for future study and scale up. | A stigma reduction campaign was conducted at the outset to increase knowledge about mental health in the community and reduce stigma related to mental health. This was perceived as a key step to ensure that the services were availed and the importance of CMD was understood. During the intervention, the ASHAs re–screened all eligible adults and identified 238 individuals (4.75%) as ‘screen positive’ (72.7% female). There were only 38 individuals who were commonly identified as screen positive at both the baseline survey by interviewers and screening done by ASHAs at the beginning of the intervention. Thirty of the individuals identified as screen positive by ASHAs visited a doctor, with 19 visiting camps and 11 visiting the PHCs. Of these, 24 (80%) were female. Eighteen of these 30 individuals were diagnosed with a confirmed mental health problem by the mhGAP– IG tool. The increase in mental health service use in this population was from 0.8% at the beginning of intervention to 12.6% at the end of intervention. After adjusting for clustering, there was a significant reduction in score of depression by 3.6 (SE 0.6, P<0.0001) and a reduction in anxiety score by 1.3 (SE 0.4, P=0.004), at the end of intervention. The results from the anti–stigma campaign show that during the project period, the communities’ knowledge, attitude and behavior related to mental health had consistently shown an improved lower score, especially for the attitude and behavior related questions The authors found the intervention of training, task shifting, and referral model to be feasible, acceptable to community and health care providers. |  | - | As this is a pre–post design with no controls, hence the results need to be interpreted cautiously. Due to the small numbers, the authors had not adjusted for the clustering effect of villages for this analysis. The 3–month intervention period was identified to be short by the authors as it allowed only 30 screen positive individuals to access services. Lastly, information about adequacy and effectiveness of treatment was not available. |
| Balakrishnan et al (63) | 2016 | Quantitative (cross sectional) | Bihar | Accredited social worker, ASHA, Anganwadi workers, Nurses , Midwives, Lady health supervisors | 550 | To assess the effectiveness of the Continuum of Care Services (CCS) mHealth platform in terms of strengthening the delivery of maternal and child health (MCH) services in a district in Bihar, a resource-poor state in India. | Training was provided to the frontline workers through an incremental learning process. Additional supportive supervision was made available during the intervention at the field level. Between the period of 1st July 2012 and 10th March 2015 a total of 19,880 pregnancies and 19,888 children were registered and provided the continuum of care services. A total of 3,09,733 home visits were provided for these women and children. It was seen that registration of pregnancy, early registration, complete 3 antenatal visits, receiving at least 90 iron and folic acid tablets, institutional delivery, early initiation of breastfeeding and post natal home visits were all higher in the implementation area compared to rest of Bihar during the same time. It was also seen that these indicators were better in the same district compared to the previous year. |  | By virtue of its impact on quality, efficiency and equity of service delivery, health care manpower efficiency and governance, the mHealth inclusion at service provision level can be one of the potential strategy to strengthen the health system. | The data from the mHealth district was compared with data from rest of Bihar as well as historical data of the previous year. This could lead to potential confounding effect in analysis. Since raw data were not accessible from the maternal and child tracking system and only summarized data was used in the analysis, fixed or random effects modeling could not be performed to assess the effects of this limitation. This paper does not report the actual reasons for why certain outcome measures did not improve substantially following the intervention i.e., iron and folic acid uptake. |
| Modi et al (64) | 2016 | Quantitative (cross-sectional) | Gujarat | ASHA workers and their beneficiariies | 844 families | To determine the proportion of pregnancies, deliveries, and infant deaths (events) being registered through the ImTeCHO application against actual number of events in a random sample of villages. | For the pregnancies, deliveries, and infant deaths found in household survey, the sensitivity of the ImTeCHO application with the household survey was 97% (95% CI 0.85- 1.00), 99% (95% CI 0.93-0.99), and 100% (95% CI 0.46-1.00), respectively. Near-complete registration is ensured via numerous safeguards incorporated into the program that includes linking performance-based incentives with digital record, demand generation, regular feedback, and point of care registration. |  | The use of mobile-phone technology and strategies applied during the ImTeCHO implementation should be upscaled to supplement efforts to improve the completeness of registration. | One of the limitations of the study was its small sample size. Also, SEWA Rural has been active in the study area for more than a decade, which might have positive influence on the results. |
| Smith et al (65) | 2015 | Qualitative (semistructured, individual interviews) | Bihar | ASHA workers and physicians | 15 | To assess the potential for using mHealth in cardiovascular disease (CVD) management in Kerala by exploring: (1) experiences and challenges of current CVD management; (2) current mobile phone use; (3) expectations of and barriers to mobile phone use in CVD management | The participants reported that communication networks offered by mobile phones could potentially benefit all stakeholders involved by improving accessibility to health information. Health workers could improve their own clinical knowledge, as well as access decision-making information whenever and wherever they may require it. The intervention would remove the geographical and time restraints that are often encountered in busy, remote health centres. Data could be shared quickly and efficiently through mobile phones. This enabled remote monitoring of disease and treatment adjustment. Thirteen of fifteen participants reported the potential benefit of receiving reminders about important aspects of disease management, such as appointments, medication and healthy lifestyle changes, in order to improve disease management. Although two ASHA participants noted that cumulative use of mobile phones is costly, patients focused on the potential cost-saving opportunities mobile phones could offer. ASHAs recognised the potential for mobile phone use to improve their work efficacy. | Participants acknowledged that one must know how to use the different functions of mobile phones in order to reap any potential benefits. Physicians were the main advocates against the use of mobile phones. With busy work schedules, they were concerned that increased use of mobile phones will become a disturbance to their work and personal lives and they felt that unregulated phone use may appear unprofessional and pose a threat to patient care. Increased mobile phone use may result in fewer patients attending appointments and hinder the need for physical patient examination. Four health workers mentioned that the effects of radiation act as a barrier to mobile phone use. They were concerned about mobile phone overuse and the unintentional harm it may cause to the body. | Successful mHealth design, which takes barriers into account, may complement current practice and optimise use of limited resources. | The researcher acknowledges that data saturation is a disputable concept and novel themes may have emerged from further interviews. |
| Gera et al (66) | 2015 | Quantitative (cross-sectional) | Rajasthan and Uttar Pradesh | Frontline health workers and block level health workers |  | To evaluate the performance of MCTS and identified implementation challenges in areas in Rajasthan and Uttar Pradesh (UP). | The overall performance results were 34 % for sampled pregnant women, and 33 % for sampled children in the Rajasthan study areas. In the UP study areas, the results were 18 % for sampled pregnant women, and 25 % for sampled children. Low data completeness rates were the primary factor leading to these poor performance numbers. While the Rajasthan sample had the higher MCTS beneficiary registration rates out of the two assessed states, the completeness of these profiles was 64 % for both sampled pregnant women and children. Accuracy rates in Rajasthan and UP for data on sampled pregnant women were at 86 %, and 79 % respectively. Accuracy rates for data on sampled children were at 92 % for UP and 71 % for Rajasthan. | The root causes of these data quality weaknesses lay in suboptimal field level data collection, consolidation and transfer processes, inconsistent training levels for health staff and a lack of clear monitoring and supervision guidelines. In all surveyed areas, there was an absence of standardization in the data tools, and data processes. In UP there was a shortage of MCTS-trained staff, with the exception of data entry personnel, in the assessment areas. One reason for incomplete MCTS portal data is the incompleteness of the primary data source. In UP, there was a lack of linguistic standardization in the names of data fields between the MCTS register (primary data source) and the portal. For example, the birth dose of the Hepatitis B vaccine was recorded as “Hep B1” in the register, while it was labelled “Hep B0” in the portal. These discrepancies compromised data quality. Irregular electricity supply, inconsistent internet connectivity and the slow speed of the MCTS web portal were some of the challenges faced by block-level facilities, which act as the primary MCTS data entry points. In Rajasthan, interviews highlighted issues of work burden for data entry personnel, as MCTS data entry was an additional charge imposed on existing staff with other responsibilities. | There is an urgent need to create data processes and supervision guidelines that complement existing workflows and service delivery priorities. Health staff should be trained to implement these guidelines. MCTS outputs, such as service delivery planning tools, should replace existing tools once data quality improves. | These findings may not be representative of the MCTS’ performance in India as a whole. |
| Modi et al (67) | 2015 | Qualitative (cross sectional) | Gujarat | ASHA workers | 45 | This article describes the process of development and formative evaluation of a complex mHealth intervention (ImTeCHO) to increase the coverage of proven MNCH services in rural India by improving the performance of ASHAs. | A total of 1,100 pregnant women and 1,422 children were registered by the ASHAs. All ASHAs demonstrated enough competencies to get certified to use ImTeCHO. None of the ASHAs stopped using the mobile phone application during the pilot. There was no reported instance of any beneficiary or ASHA refusing to participate. The average login rate during the study period was 88%; however, the login rate for medical officers was only 17%. During the pilot phase, 10,774 tasks were generated, of which 7,710 (71%) were completed by the ASHAs. During the interviews, all ASHAs found the ImTeCHO and its components quite acceptable, feasible, and useful. Overall, all ASHAs found delivery of the intervention largely acceptable and implementable. The process indicators, available in real-time with the help of mobilephone technology, were an innovative tool that proved quite useful for monitoring delivery and adherence to the intervention. Also, the ANMs and medical officer took on more and more of the responsibility for providing patient care. To conclude, ImTeCHO and its delivery were found to be acceptable, feasible, and useful within the primary health care system and the ASHA program. | Some of the issues that arose during the pilot are listed here: 1) Some beneficiaries found the videos monotonous after a certain amount of repetition. 2) Scheduling of home visits was appreciated; however, ASHAs expressed the need to add scheduling for services to be provided during VHND as well. 3) None of the ASHAs reported any additional instances of proactive supervision from PHCs through the use of ImTeCHO. 4) Limited phone memory occasionally created technology-related issues, with the increasing requirement to store data on the mobile phone. 5) The PHC staff found the ImTeCHO incentive management system rigid and less user-friendly. 6) Higher-level officials expressed the need to add features to facilitate and assess the progress made in enrolling new pregnancies and delivery outcomes. It became clear that the mobile phone application lent a certain degree of formality to the visit. Two ASHAs experienced difficulty in receiving and transferring data because of lack of GPRS in their villages for prolonged period of time. At the outset, there were quite a few instances of ‘false positive’ diagnoses due to ASHAs inadvertently entering the wrong information in the application; however, the proportion of false positive diagnoses sharply declined over the study period to less than 5% at the end. |  |  |
| Thukral et al (68) | 2014 | Mixed-Methods (pre-post tests, Likert's scale and focus group discussion) |  | Nursing students | 27 | To evaluate the efficacy of interactive mobile device application ‘Apps on sick newborn care’ as a training tool, in improving the knowledge and skill scores of postgraduate nursing students. | Training resulted in significant improvement in the post-MCQ score (19.7 3.6 vs. 12.5 2.2; P < 0.001) and the composite OSCE score (63.6 7.1 vs. 32.8 7.3; P < 0.001). Similar observation was also noted in the individual OSCE scores. The participants expressed overall satisfaction from the course as evident by their scores on the Likert’s scale. The training contributed to their level of confidence and proficiency. They felt that the training was very useful for their professional activities. All their doubts were cleared and they felt that they would be able to implement the techniques learnt into practice. They found the study material interactive and easy to learn. They also felt that the algorithmic approach and the related videos were very helpful. All the participants knew the use of computer Internet before attending the workshop and recommended similar activity for their colleagues when asked. The main strengths of Apps were its ability to provide instant support to a remotely placed caregiver, without hassles of high-tech connectivity or operational cost. | Some of the potential obstacles in implementing Apps were uncertainty regarding the availability of the device, lack of awareness of Apps, resistance to change amongst senior colleagues, leadership concerns, likelihood of overdependence causing disappointment in case of technology failure. However, some of them wanted more individual time dedicated to the workstations. | Such applications have potential to train health-care professionals. Apps have a high potentiality to invade into our class rooms as a ‘supplement’ if not a ‘substitute’ in the near future. | The study sample the authors drew was of MSc nursing students, who may not share the same kind of competencies and attitudes of general practitioners working at the district level. The evaluation was based on the immediate outcome in terms of knowledge, skill and attitude, rather than long-term impact. The quality of trainers and the motivational level of our participants could have contributed to the success, rather than mere efficacy of the instrument. |
| Jarosławski et al (69) | 2014 | Qualitative (interview based study) | Pan India | Designers, implementators, technology holders for ehealth programs | 30 | To understand the kinds of eHealth programmes being offered in India today, the challenges they face and the nature of their financing. | The ASHAs have been provided mobiles that are usually Java-based. The mobiles increase their stature in the community, and the message on the mobile is also taken more seriously than the spoken word. It improves the quality of care, including follow-up and emergency care, and the number of referrals. Monitoring the productivity of rural health workers, who normally work without supervision, is an important aspect of these programmes. A separate initiative involves instructional videos that are preloaded on a micro-SD card for a mobile phone, that is purchased in small shops. There are key messages on 14 health issues, such as hand washing, oral rehydration therapy and exclusive breast feeding, that are derived from a document that is endorsed by organizations of the United Nations. These serve to educate the ASHAs and beneficiaries directly as well. Some of the ASHAs are highly motivated, contributing to the success of the programmes. | For outreach workers using mobiles, data collection is not straightforward, given widespread illiteracy or semi-literacy. Although the software is sometimes designed to work on both Java and Android phones, it usually has to work on lower cost mobiles. Even if some data is collected electronically, the remainder has to be done by paper, lowering the incentive to shift to electronic records. There are several technological challenges in telemedicine. First, those who design the mobile applications may do so without much understanding of the ground reality, making them sub-optimal. Second, as mentioned above, there is a trend to constantly upgrade software while the existing one is still functional. Instead of implementing a project with the current software, money is spent on developing and piloting a new version. Low end mobiles are widespread, but cheap smartphones such as the open-source Android devices would be better suited for people with poor literacy, and for applications that employ a global positioning system (GPS) tracking function. However if one upgrades to a smartphone, one needs an application that may no longer run on a simpler phone. Third, the novelty of the technology has required extended learning and optimization periods for both manufacturers and implementers, thereby slowing or decreasing its spread, especially in the early years. Fourth, the electric supply and network coverage can be poor, which is sometimes circumvented by the store-and-forward modality. If the government subsidised it, a satellite connection could be used where internet connectivity is absent, but no new satellite-based connections are being established. Fifth, companies that were pioneers in this area were forced to come up with their own technical solutions. Thus, hospitals may use proprietary software that is not Dicom compliant, and therefore not transferable across hospitals with different patient flows, employment structures and so on. Since IT and medical professionals don’t really understand each other, developing appropriate software is not straightforward. The shortage of skills is evident in a variety of settings, as follows. (i) Although supposedly educated up to middle school, some ASHAs are illiterate and can barely answer a call. Therefore learning how to use the phone can be a challenge. Their professional training may also be poor, leading to low quality counselling of beneficiaries. (ii) Technicians who work in the telemedicine centres are not skilled in computer use and need to be trained. This can lead to high training expenditure and staff shortages. (iii) The NGOs and research groups in health and population studies, and in the government, may find technology challenging. There may be reluctance to learn something new, and even if provided a ready product they will not necessarily make full use of it. Users’ acceptance of both technology and mode of healthcare delivery is important, and convincing them may take considerable effort. One of the biggest challenges facing most of the programmes concerns funding and sustainability. | It is unlikely that eHealth will have widespread and sustainable impact without government involvement, especially in rural areas. Nevertheless, programmes run solely by the government are unlikely to be the most effective. | The authors were unable to record the interviews, and thus if there were nuances that would have been picked up in the subsequent analysis of recorded interviews, mheyay have missed them. Although the authors interviewed a wide set of organizations, the number for most of the categories was low. |
| Rajasekaran et al (70) | 2013 | Quantitative (cross-sectional) |  | Doctors, Nurses and Health inspectors | 280 | Mobile technology helps to improve continuing medical education; this includes all aspects of public health care as well as keeping one’s knowledge up-to-date. The program of continuing medical and health education is intertwined with mobile health technology, which forms an imperative component of national strategies in health. Continuing mobile medical education (CMME) programs are designed to ensure that all medical and health-care professionals stay up-to-date with the knowledge required through mobile JXTA to appraise modernized strategies so as to achieve national goals of health-care information distribution. | Users underwent training and took online exams; they obtained high scores half of the time. Users who completed the training did so in 54 minutes or less and tested higher on the final assessment tests than the others of the firm. Of the 250 eligible employees, 6% launched the content at least once and 56 people completed the courses. Overall, the mobile learners obtained a 56% higher knowledge gain completion rate in 30% less time than the control group. A total of 70 employees responded to a survey indicating more than 75% praised the benefits of convenience, time management, and training through mobile JXTA. | - | Compared with traditional learning system, enhanced study improves cloud-based mobile medical education technology.. | - |
| Goel et al (71) | 2013 | Review | Pan India | doctors,nurses,midlevel workers |  | The aim of this paper was to review published and unpublished literature, field projects, and pilot studies on mHealth usage in overcoming shortage of human health resources in developing countries. | Health workers used simple mobile phones for data transfer, and negligible errors were reported in the process. This process helped in completing the survey in a record time of 20 days, as use of mobile phones significantly reduced transport time and separate time required for data entry and transfer. A pilot project Real-Time Biosurveillance Program was implemented by Sarvodaya and LIRNEasia to investigate the conditions for effective deployment of wireless technologies in disease surveillance. This project proved effective in the early detection of clusters of population affected by chicken pox, acute diarrheal disease, respiratory tract infection, dengue, and viral fever in Kurunegala district, Tamil Nadu, India. Jalaaka Project in India facilitated education and spreading awareness about HIV among the community and high-risk groups. mHealth has also been used for educating community in projects focused on family planning, adolescent health, and antenatal care. Community health workers in Bihar, India, used mobile phone application CommCare to disseminate information on common adolescent health issues such as menstrual hygiene, sexually transmitted diseases, and family planning methods among adolescent girls and women. The outcome of the project was encouraging in that it had a wider outreach among adolescents, even as health workforce was scarce. Mother and Child Tracking System under National Rural Health Mission in India was introduced to ensure delivery of comprehensive maternal and child health services through the use of mobile phones. Work plans and reminders for antenatal and postnatal checkups were sent via mobile phones to respective health workers. This eliminated the need for paper work to draft the work plan and manual delivery. Repeated reminders through phone calls and SMS ensured better and timely health care delivery to the beneficiaries. | - | mHealth care can bring about a revolution in health care in India and other developing nations. | - |
