## Supplementary material for "Barriers and Facilitators for the Use of Telehealth by Healthcare Providers (HCP) in India - A Scoping Review": S2 Table

S2 Table: Study characteristics for studies on telemedicine as mentioned by the authors.

| Study identification* | Year of Publication | Study design and setting | States Studied | Population studied | Sample size | Primary objectives | Positive findings/Facilitators | Barriers | Implications for healthcare delivery/ universal health coverage | Limitations of the study |
| --- | --- | --- | --- | --- | --- | --- | --- | --- | --- | --- |
| Verma et al (87) | 2022 | Quantitative (Cross-sectional study) | Not available | Patients availing hepatology services | 245 | To study the feasibility, outcomes, and safety of telehepatology in delivering quality care amid the pandemic. | Patients were likely to recommend telehepatology services to their near ones in the prevailing situation. Overall, 208 (99.0%) and 178 (84.8%) of patients reported teleconsultation as convenient and comfortable, respectively. During the pandemic, 176 (83.8%) preferred teleconsultations, whereas 133 (63.3%) would prefer physical OPD after the pandemic. Overall, 179 (85.2%) patients were satisfied with the teleconsultation, and 104 (49.5%) patients reported improvement in their overall health. Diagnosis was achieved in 197 (94%) patients, and 173 (82.4%) patients complied with the treatment with an overall success rate of 46.2%. Patients found telehepatology to be convenient and comfortable, with good confidence rate, diagnosis rate, and acceptable understanding (85.7%). It was also reported to be economical with respect to both time and money spent due to the savings on travel and accommodation. | The productivity rate of physical OPD was higher as compared with tele-OPD. The time spent (in minutes) per consult was more in teleconsultation than physical OPD. Treating physicians deemed 88% patients suitable for tele-evaluation alone; unfortunately, nearly 20% made an unscheduled appointment due to improper understanding or persistence or worsening of symptoms. Time taken per patient was lower in physical OPD because of more effective face-to-face communication, judicious use of clinical acumen, and better building of the doctor–patient relationship. Connectivity, data exchange, and language issues, the potential conventional barriers to teleconsultation,(27) were reported by less than 10% patients, whereas more than 70% did not report barriers. Reimbursement, one of the main limiting factors for telemedicine in the Western and European countries, was considered an impediment by less than 1% of our patients, likely due to the free cost government health services in our country and posttreatment reimbursement by local state governments. The primary connectivity issue faced by patients was the inability to register, sometimes due to busy telephone lines because of a relatively narrow time period of 2 hours for registration. | Telehepatology could possibly be integrated with the conventional healthcare delivery system. | Limitations of the study include relative immaturity of telehepatology setup, which may have led to some inaccurate assessment of parameters like level of understanding, connectivity rate, improvement rate, and success rate. There was a lack of data for patient centered outcomes, safety, and success rates from physical OPD consults. |
| Angral et al (92) | 2021 | Review | Not available | Not available | Scoping review with 16 articles | To assess tele-otology and tele-audiometry practices in India. | The intervention takes more time and can be conducted by non-medical persons. It is a safe and reliable method of screening out the hearing-impaired individuals from the general population. So, no significant difference was noted in PTA and DPOAE performed via in-person and tele-hearing methods. The feasibility of tele-ABR in mobile van with satellite connectivity was compared with ABR recordings made face to face in 24 new-born individuals. No significant difference was seen in these two modes in the peak V latencies at three intensity levels. Real time tele ABR testing is a feasible component for new-born hearing screening with assistance of VHWs. Tele ABR group presented 11% improved follow-up rate. The use of remote diagnostic tele-audiology in rural population has the potential of long-term cost saving in developing nations. Parents experience was reported good and found it to be better than the in-person method. Access to tele-hearing testing it has the potential of reducing rural travel time considerably, as the parents had mentioned less than 30 min travel time for seeking health services. Parents preferred tele van mode of hearing testing because of easy accessibility through video conferencing with a bigger TV screen and stable satellite connectivity. Parents were satisfied about the counselling, testing process and accessibility because of the logistic factor like reasonable travel time, accompanying local VHW, cost free testing and technical factors like good video quality. | Technical issues related to internet connectivity resulted in video time lag and difficulty in screening. Difficulties faced while implementation of hearing screening programme in India were audiologist’s shortage, lack of infrastructure, difficulties in providing services to rural population and poor follow-up rates in distant tertiary centres. Poor video quality and inaccessibility of the audiologist were the reasons mentioned by those who were not satisfied. Other barriers included - Instructions in English, requirement of a quiet room, and the fact that android application uses pure tones which are less reliable than speech audiometry. | Web based hearing assessement is particularly applicable for rural population specially during covid pandemic. | - |
| Nair et al (96) | 2021 | Quantitative (Cross-sectional study) | Tamil Nadu | Patients living with epilepsy | 141 | To assess the feasibility, satisfaction, and effectiveness of video teleconsultation using mobile phones for managing persons with epilepsy (PWEs) on follow-up at a tertiary care center in the southern part of India. | A successful video connection was achieved in 95 (28.3%) PWEs, and only audio consultation in 46 (13.7%). All registrations were completed on the same day. After teleconsultation, prescriptions were provided to all 95 PWEs, and for 18 (18.9%) of them, new medications could be added. The physician felt that the video quality was good in 68. (71.6%), average in 22 (23.2%) and bad in five (5.3%), and audio quality was good in 62 (65.3%), average in 24 (25.3%), and bad in 9 (9.5%) connections. During the assessment of satisfaction of video teleconsultation, more than 90% of participants either ‘agreed’ or ‘strongly agreed’ on 12 out of 14 questions such as – they could easily see, hear, talk, communicate, get enough attention, fulfill their healthcare needs, and save time. They also opined that they would like to receive teleconsultation again in the future and were overall satisfied with teleconsultation. Forty-six (48.4%) participants either disagreed or strongly disagreed that assistance is needed for teleconsultation. | Lack of smartphones in 75 (39%) and inability to contact participants in 71 (36%) due to various reasons such as out of coverage area, phone being switched off, and not responding to calls and lack of connectivity were identified as barriers. Only 19 (20%) participants could connect successfully in the first attempt itself, whereas remaining people required two (47.4%) or three (32.6%) attempts. The main reason for more than one attempt and extra time for consultation was the difficulty in teaching the participants how to enable the video facility in their smartphones. | This real-time model has the advantage that it does not require any mobile application to be downloaded and installed. | The limitation of our study is the cross-sectional nature with its inherent drawbacks. The study did not have a control group to compare. |
| Naik et al (95) | 2021 | Quantitative (Cross-sectional study) | Karnataka | Patients with psychiatric disorders | 1049 teleconsultations | To assess the utility of telephonic aftercare services (including liaising with primary healthcare providers) rendered to persons with psychiatric disorders during the lockdown period of COVID 19 pandemic in India. | Seventy one patients expressed their preference for teleconsultation and 29 opted for in-person consultation at NIMHANS. All these patients reported higher satisfaction and acceptance of clinician-initiated teleconsultation services and did not find this approach to be intrusive. These patients opted for teleconsultation if regular supply of their medicines from DMHP or local pharmacy is ensured. | Personal satisfaction was low in patients who used teleconsultation. Low digital illiteracy, healthcare system related reasons, and lack of in person contact with doctor were identified as limitations. | - | - |
| Ragesh et al (86) | 2020 | Quantitative (Cross-sectional study) | Karnataka | Mothers discharged from a Mother Baby Unit | 76 mothers. | To assess the feasibility, acceptability, limitations, and usage patterns of a helpline service for mothers discharged from a mother-baby psychiatry unit. | Of the 76, 57.9% (n = 44) had used the phone helpline. When asked about the usefulness of the helpline, of the 44 users, 93% (41) said that it was a useful service, 91% (40) said they got a helpful response, 95% (42) said they would recommend this service to others. The psychiatric social worker who was in charge of answering the helpline and held the phone, reported feeling comfortable in handling the calls. | Language and communication difficulty, problems in getting in touch, not knowing the helpline number, and nonavailability of a phone might be a limiting factors. Difference in language, difficulty in handling matters related to interpersonal relationships over the phone, difficulty in persuading the family members and the patient about the need for inpatient care if there was a suicidal risk or relapse/exacerbation of symptoms and difficulty in contacting the local psychiatrist in case of psychiatric emergencies were identified as barriers. | This model could be adapted in other mother-baby psychiatry units in low and middle-income countries. | The main limitation of the study is that the authors could not get in touch with all the mothers to whom the helpline number was provided. |
| Biswas et al (91) | 2020 | Quantitative (Prospective cross-section analysis) | Delhi | Patients availing telemedicine services | 314 patients | To assess how palliative medicine physicians could follow up on cancer patients and barriers they faced, discuss their results, and evaluate their treatment response with the help of telemedicine. | The majority of the patients availed telephone calls and text messages (n = 167, 53.18%), whereas video consultations were required for 84 (26.75%) patients. Teleconsultation removed barriers like restriction of movement across the state borders and lack of transport availability (in 124 patients), terminal patients (in 88 patients), and fear of getting infected (in 71 patients) 56 patients were very satisfied and 152 patients were satisfied with the service. However, 59 patients remained partially satisfied and 47 patients were unsatisfied. | Unavailability of multidisciplinary advises over a single call was a barrier. 42 patients believed that face-to-face consultations may be more useful for them. | Telemedicine is the future of health-care delivery systems. Palliative Care deals with immunocompromised debilitated cancer patients and telemedicine is immensely helpful to provide holistic integrated care to these patients who are unable to visit hospitals regularly. | The authors were unable to assess the psychological aspects of the patients and caregivers. Patients who were advised for another PC unit visit, emergency department of nearby hospitals for symptom management, or provided with the contact details of other departments could not be followed up. |
| Ravindran et al (77) | 2020 | Quantitative (cohort study) |  | Patients | 184 | To describe the preliminary experience in providing psychosocial support amid the COVID -19 pandemic from a Tertiary Care Centre in India. | Over 90% callers were satisfied with the provision of the service. Callers reported that the mental health professionals were able to address their psychological distress, were able to help with linking to local resources, thereby addressing their concerns, and most of them reported that they would call back the helpline if in crisis. Hand-holding by the more expereinced mental health professionals was extremely helpful to boost their confidence in handling callers whose concerns may bring about negative emotions in the volunteer. | Nearly 50% of the calls were not connected to mental health professionals possibly due to limited number of lines. A few individuals however expressed discontent of this helpline service being unable to directly help in logistical concerns such as travel related and monetary concerns. Being unable to see a distressed individual face-to-face as it is in an usual psychiatric practice, was challenging as mental health professionals were unable to guage facial expressions, and body language and had to rather rely on their skills to pick up the same over a phone call. The volunteers coming from varied backgrounds and expereince in mental health, was a challenge to make the service as uniform as possible. Individuals with acute psychiatric emergencies posed difficulty to address over a phone call. | This intervention could play a major role to reach out to the masses in all phases of this biological disaster. | - |
| Panda et al (84) | 2020 | Quantitative (cohort study) | Uttarakhand | Children with epilepsy | 278 telephone consultations performed for 153 children | To explore the feasibility and efficacy of advanced telecommunication measures to provide an allinclusive and precise teleconsultation for children with epilepsy. | No untoward events occurred in the seven children as determined on follow up telephone calls. A total of 152 significant clinical events were identified during telephonic consultation in 113 children. Out of the total 278 telephone calls of average 7 min duration, only 5 calls were interrupted by technical glitches/network error and 147 (96 %)caregivers were satisfied with medical advice provided over the telephone. | Twelve caregivers were able to comply while the remaining seven caregivers did not have well-equipped smartphones to have a good quality video recording of seizures. Further clinical information through picture message/video recording could not be obtained in them and the treatment was prescribed based on the best possible clinical judgment of the investigator. 27 caregivers were unable to understand English prescriptions. | This intervention might be the only feasible option to cater to quality health care to children with epilepsy all over the country, clinicians and public health officials need to work in this direction to make its widespread use. | - |
| Gupta et al (75) | 2020 | Quantitative (cohort study) | Delhi | Community Health Workers, Patients | 5 CHW and 3000 patients. | To assess the feasibility of empowering trained health workers equipped with ENTraview, a store-and-forward telemedicine device that integrates a camera- enabled smart phone with an otoscope. | Of the 3,000 patients screened, health workers identified 1,619 (54%) patients requiring examination by an ENT specialist or an audiologist, who were then referred to the base hospital. | Poor interpersonal and communication skills in the local language of the community to be screened were identified as barriers. | The presented model of screening patients with hearing disorders has the potential to change the way the community and national hearing programs are run in the developing world. | - |
| Rout et al (74) | 2019 | Quantitative (cohort study) | Maharashtra | anti-retroviral treatment (ART) centers | 6 ART centers | To estimate the unit cost of ART services for pediatric HIV patients and examine the efficiency in the use of resource and treatment compliance resulting from telemedicine initiatives in pediatric HIV compared to usual ART services. | The results clearly demonstrate that the PCOE-linkage resulted in a cost savings of Rs 1610/ visit in the PCOE-linked center as compared to non-linked centers. The linking of per-pediatric patient cost with treatment compliance showed a 5 percentage point improvement in lost to follow-up in the linked centers compared to non-linked centers against a back-drop of a reduction in per-pediatric patient cost of INR 557. As mentioned in the table the per-patient cost in the linked ART centers was INR 4513 as compared to INR 5069 in the PCoE-non-linked centers. The findings clearly suggested that the timeliness of the visit increased in the linked ART centers compared to their counter parts. However, as the initiative matured over time, the effect of the telemedicine support from PCOE was tangible as the mean/median time-interval dropped in the linked ART centers as compared to their non-linked counterparts. | It was observed that—the median/mean time-interval was slightly better in the linked ART centers compared to nonlinked centers, however, the proportion of visits without delays started declining. This suggested that the advantages gained could not be sustained during the last phase of the analysis. This could be due to several reasons: systems “fatigue” an important reason among them. | These advantages demonstrated through this initiative could be encouraged and scaled-up in similar settings. | The study did not use more commonly used outcomes of “cost effectiveness” studies—Disability Adjusted Life Years (DALYs) and death averted, because of limited availability of data in a mid-term evaluation, the objective of which was to generate evidence whether the intervention is in right direction and is achieving the intended results. The sample size was small, 3 centers in both the arms, designed so, keeping in view the time and resource constraint in this mid-term evaluation framework of the project. |
| Ramkumar et al (89) | 2019 | Quantitative (Prospective cohort study) | Tamil Nadu | Village Health Workers | 6 Village health worker screened 1335 children in group A and 1480 in group b | To explore a community-based, pediatric hearing screening program in villages, integrating two models of diagnostic ABR testing; one using a tele-medicine approach and the other a traditional in-person testing at a tertiary care hospital. | Training was conducted in the local language (Tamil) and included PowerPoint presentations, hand-outs and videos. The post-training evaluations showed considerable improvement in the scores obtained by VHWs on questions related to importance of age of identification of hearing loss and effective methods for screening children for hearing loss. The average coverage rate was 77% (Group A: 65%; Group B: 90%). Time taken for screening ranged from 10 min to 60 min. The median follow-up rate for 2nd screening was 85%. In-person ABR was recommended for four children referred in the 2nd screening and three followed up, resulting in 75% follow-up rate. Tele-ABR was recommended for 20 children referred in the 2nd screening and 17 followed-up. In addition, two children who passed the screening but subsequently developed ear infections were also asked to follow up for tele-ABR and both followed up, resulting in 86% follow-up rate. | - | In this community-based hearing screening program, tele-ABR improved follow-up rate when compared to in-person ABR. Can help in establishment of large scale hearing screening programs in India. | - |
| Patel et al (82) | 2018 | Quantitative (Randomised Control Trial) | Maharashtra | Mother-infant pairs | 1036 | To assess effectiveness of cell phones for personalized lactation consultation to improve breastfeeding practices | The rates of exclusive breastfeeding were sustained above 95% at all visits in the cell phone group but dropped from 81% at 6 weeks to 48.5% at 6 months in the control group. Each woman who received the intervention was six times more likely to exclusively breastfeed her infant for six months in comparison to those women who received standard healthcare services. Infant illness was reported in 77/1031; 7.5% (control: 45/513; 8.8% vs. intervention: 32/518; 6.2%) of cases of mothers not exclusively breastfeeding, whereas, maternal illness was reported in only 8/1031; 0.8% (control: 5/513; 1.0% vs. intervention: 3/518; 0.6%) of cases. Rates of initiation of breastfeeding within an hour of birth were significantly higher in the intervention compared to the control . The rates of infant hospitalization (neonatal intensive care unit admissions) were significantly lower in the intervention at visit 3 (12.5% v/s 6.8% p < 0.01).. The mean weight of babies at delivery was similar in both groups, but infants in the intervention group weighed significantly more than those in the control group at each subsequent visit. In the intervention, 92.3% of the women were completely satisfied with breastfeeding counselling provided by the lactation counsellors over cell phones. It was reported by 93% women from the intervention that the information received by them was helpful. In the control, only 36% of women were completely satisfied with the breastfeeding counselling provided by the health care provider and 31% felt that all the information they received regarding breastfeeding was helpful. | The average total cost incurred by all the subjects in the study from third trimester to 1 week and 6 months after delivery was Rs.4687. The point estimate of incremental cost-effectiveness ratio showed that it was costlier [5603; 95%CI (5587, 5619)] to receive cell phone counselling. Other implementation challenges such as switched off phones, discharged phones, rejected calls or unanswered calls, calls received by someone other than the enrolled women and loss of cell phones were also encountered during the study. | This intervention shows immense potential for scale up by incorporation in both, public and private health systems. | The limitation of the study was that it was an unblinded pilot study of only four clusters. Another limiting factor was that the intervention was not designed to assess the effectiveness of different frequencies of contact with the women, on exclusive breastfeeding. |
| Khanna et al (85) | 2018 | Quantitative (Retrospective cross-sectional study) | Karnataka | Patients who were provided with tele-neuro rehabilitation services | 37 | To study the socioclinical parameters, feasibility, and utility of telemedicine services in India. | Overall, telemedicine was found technically and operationally feasible in a developing country like India. Availability of technicians ensured further support in the operation and resource feasibility of the model. This model eventually reduces health cost burden and reduces health and time gap to reach experts in the field. Telemedicine makes universalization of health services accessible to patients in their vicinity. It also helps in providing consultation services, education and training to the patients’ families, and follow-up and monitoring of teleservices. | Lack of coordination in cross-consultation in telemedicine services within the institute was identified as a barrier. Poor follow-up was observed in the study. There are multiple challenges and potential barriers to telemedicine practice, the primary concerns being privacy and confidentiality while providing teleservice. | The findings suggest that the services are feasible, effective, and less resource intensive in delivering quality telemedicine care in India. With effective collaboration with district hospitals’ clinicians/staff, specialists at the tertiary/quaternary care center and telemedicine department can help in treatment and rehabilitation of needy population and universalize health into all sectors. | However, the study has limitations such as small sample size and outpatient consultation only. Another limitation is small number of district hospitals in the entire state utilizing tele-rehabilitation services. |
| Manjunatha et al (78) | 2018 | Qualitative (cohort study) | Karnataka | Primary Care Doctors | - | Goal of this paper was to provide an overview of all these (five) modules - orientation module, basic module, advanced module [Tele-psychiatric ‘On-Consultation Training’ (Tele-OCT)], videoconference-based continuing skill development module, and collaborative video consultation modules with its various stages of implementation. | Available videoconference-based telemedicine facility at Maddur General Hospital was used to simulate OCT sessions in subsequent extension clinics with randomly selected patients from the general outpatient pool. This led to a smooth transition from IP-OCT to Tele-OCT, which addressed the challenge of travel of trainer psychiatrists. In this process, a couple of sessions could demonstrate acceptance among Primary Care Doctors (PCDs) and the technical feasibility of Tele-OCT. The advantages of Tele-OCT over IP-OCT are its perceived cost-effectiveness in terms of travel time and money spent. Tele-OCT is a friendly training program from both PCDs' and psychiatrists perspectives. The acceptability of Tele-OCT was found to be high. | The inherent disadvantage of OCT is it is labour intensive. Furthermore, it is expensive in terms of the salary of the telepsychiatrist but worth it in terms of perceived public health benefits from this one-time spending money on a telepsychiatrist. Considering the huge number of PCDs in the country, it is a daunting task to have an adequate number of trainer telepsychiatrists. In tele-OCT sessions, PCDs may not be comfortable in front of their patients. Most PCDs express apprehension about sessions before they come; once the session begins, they feel comfortable and involve themselves actively thereafter. | Once validated thoroughly, Primary Care Psychiatry Program' (PCPP) has potential for pan-India expansion. | - |
| Ganapathy et al (93) | 2018 | Quantitative (cohort study) | Himachal Pradesh | - | 753 teleconsults | To describe the profile of patients seen using tele-emergency. | Presenting a clinical problem, in an emergency situation, through video conferencing, management information systems, reporting, and troubleshooting Internet connectivity problems, was taught. Appointment scheduling, live chatting, and interfacing with telediagnostic medical equipment was also possible. Personal interaction by telemedicine coordinators on both sides ensured that traditional human touch continued. A survey of 140 users of Tele-emergency service (TES) indicated that considerable effort, time, physical discomfort, and emotional stress were reduced, besides not incurring out-of-pocket expenses of about US$20,000. Nontangible benefits such as satisfaction and happinessthata caringgovernmenthasfacilitatedquality accessible healthcare round the clock and free of cost cannot be quantified. The total costs incurred for setting up the TES, including the initial capital expenditure, the recurring operational expenditure (salaries, connectivity charges, consumable charges), and the program management charges (all for the TES component), were computed. This was divided by the number of patients who utilized the TES. The cost per patient in the first 145 weeks was US$208. However, if one takes into account the average yearly depreciation on the capital expenditure, average annual cost escalation on operational expenditure, and the expected percentage increase in annual patient footfalls, when distributed over 5 years, the cost could come down to US$120 in the fourth year with a 5-year average of US$143. The local staff were virtually trained and soon became adept at using the kit in an emergency setting. This resulted in better clinical management. | Foremost was obtaining a reliable history from the remote patient or the caregiver. Most patients spoke the local dialect. To the clinician at the other end, English was a foreign language. As digital imaging was not available, hard copy images had to be photographed and sent electronically. This had limitations. The doctor at the remote end, having to interact with different teleconsultants, was unable to establish specific rapport. Lack of continuous reliable power is a concern. Diesel for generators was not always available. The only radiology technician was also not available after 4 pm or on holidays. Due to initial software glitches time of presentation was unavailable in 51 patients. It was surprising that contrary to expectations, there were only 8 patients who utilized the service between 9 pm to 9 am, although the services were available. Lack of awareness, lack of transport, and the very harsh weather conditions at this time could have been contributory factors. | Preliminary analysis confirms that delivering TES in inhospitable terrains in a Public Private Partnership mode is doable and is welcomed by the community. | - |
| Thakar et al (76) | 2018 | Quantitative (cohort study) | West Bengal | TM for elective post–neurosurgical care patients | 1200 patients | In this study, the authors evaluate the cost-effectiveness of telemedicine (TM) consultations for followup care of a large population of patients who underwent neurosurgical procedures. | The sum of the prorated costs relating to capital and personnel and total costs pertaining to travel, investigations, and lost productivity in the TM-care and routine-care groups over 52 months were INR 2,417,965 and INR 909,509, respectively. The mean per episode cost was INR 2338 for TM care versus INR 5479 for routine care. The overall TM utility was 89%, whereas that of the provider center consultations was lower at 80%. The overall effectiveness of the TM-care group was 917.4 and that of routine care was 132.8. The cost-effectiveness ratios were INR 2635 and INR 6848 for the TM-care and routine-care groups, respectively. TM care dominated routine care in the analysis. |  | TM for follow-up care for elective neurosurgical care patients is a potentially viable, cost-effective option if the provider center sets up a TM center close to a well-served remote region, and if patients are well educated about the scope and utility of the TM services. | The heterogeneous clinical profile of the patients in the 2 groups could have been a source of bias. The utility scores were based on a sample population, and this could influence the results if there was greater variance between scenarios. The use of the first follow-up episodes rather than a longitudinal approach to the analysis could potentially affect results. There could also be potential sources of over- and underestimation of costs across both groups for factors such as travel and lodging. |
| Acharya et al (83) | 2016 | Quantitative (cross-sectional study) | Telangana | Doctors and Patients | 71 patients and 51 doctors | The aim of this questionnaire study was to evaluate the effects of telemedicine on patients and medical specialists. | All the patients in this study had used the Apollo telemedicne service near their hometown either through e‑mail, telephonic conversation, or video conferencing with a specialist. The duration of treatment given through telemedicine ranges from one visit to as long as a year depending on the nature of cares given to the patient. All the doctors had telemedicine experience; 55% of the participants had 10–20 consultations per day, 22% had more than 20 consultations per day. Sixty‑five percent of participants devoted 1–2 h per day of their time for consultation using telemedicine. All the doctors were satisfied with the treatment given through TSC. Ninety‑four percent of the respondents answered that they got desirable results on the diagnosis of patient’s condition. Recall percent of patients in this study was 71%. About 94% of providers were open to promoting telemedicine. About 90% of the respondents said their appointment was scheduled according to their convenience. About 82% of the participants were satisfied with the treatment given through the medium of telemedicine and that they would recommend this medium to their relative and friends. Fifty‑four percent of the patients acknowledged that they did not have a problem in understanding the usage of telemedicine. About 89% of the patients stated that the treatment was both feasible and convenient; 7% felt treatment through telemedicine was costly for them but convenient, and 4% of the participants felt it was economical but not convenient. About 90% of the specialists reported that telemedicine was beneficial for them, and 61% patient’s inflow increased since the commencement of telemedicine practice as compared to 35% who reported no change. | Issues faced by patients with the use of telemedicine were 24% on technical issues, and 18% were not satisfied with the treatment provided of which the major issue was not comfortable to face the camera and lack of personal face to face contact with the doctor. During the consultation, the doctors faced issues on the technical subject (47%), time scheduling (39%), and few of them (14%) had problems with communication lapse. None of the respondents had problems with the staff. | In the near future, telemedicine can be considered as an alternate to face to face patient care. | The limitations of the study were that the responses of the questionnaire were prone to respondent bias; the results were limited to one group in each phase of the study. |
| Mohanan et al (80) | 2016 | Quantitative (cluster Randomised Control Trial) | Bihar | Children under 5 years of age. | 36,315 children living in 21,646 households in 2011 and 31,635 children living in 21,367 households in 2014. | This article evaluates the World Health Partners (WHP) Sky program, a large-scale social franchising and telemedicine program in Bihar, India. | The WHP-Sky program had no overall significant effects on the provision of appropriate treatment for either childhood diarrhea or pneumonia, conditional on seeking care. Specifically, there was no significant effect of implementation of the program on the likelihood that a child with diarrhea received treatment with either zinc or zinc in combination with oral rehydration solution. Similarly, there was no significant effect of program implementation on the likelihood that a child with pneumonia received a five-day course of antibiotics. We also evaluated the WHP-Sky program’s effects on secondary outcomes and found no evidence of any program impact. We also found no significant effect of the WHP-Sky program on population health outcomes. Estimated program effects on the prevalence of diarrhea (a 1.5-percentage-point change) and the prevalence of pneumonia (a 0.1-percentage-point) were statistically indistinguishable from zero. | - | The findings highlight the importance of conducting rigorous impact evaluations of new and potentially innovative health care delivery programs before investing in scaling them up. | Although the evaluation was originally designed as a randomized controlled trial, program implementation in practice deviated substantially from the original implementation plan, which rendered the original study design unusable. Second, it was not feasible to conduct complete diagnostic evaluations of health outcomes in the large sample. Instead, the authors relied on survey based measures of recall of childhood health outcomes and health care use in the previous fifteen days to minimize loss of recall, and also used the Child Health Epidemiology Research Group (CHERG) method of using video demonstration of pneumonia symptoms to improve the specificity of measurement of pneumonia. Third, as mentioned above, there was disagreement between the authors' field observations of the locations of the WHP-Sky program providers and the locations reported by WHP |
| Lindquist et al (73) | 2016 | Quantitative (cross sectional study) | Gujarat | Emergency Medicine Technicians(EMT) | 108 | To identify obstacles in the cellular communication process among GVK Emergency Management and Research Institute (GVK EMRI) EMTs in Gujarat, India. | - | Overall, ninety-seven (89.8%) EMTs responded that the most common reason they did not initiate a call with the call center physician was because of insufficient time. In assessing phone call initiation by EMTs, forty-six (42%) EMTs reported that they were unable to initiate a call to the physician one or more times during a typical workweek due to their hands being occupied performing direct patient care. Of the 46 EMTs who reported they were unable to call the call center physician one or more times a week due to their hands being occupied, thirty-nine (85%) reported that the reason was insufficient time, and seven (16%) reported it was too difficult to call. In evaluating phone call connection, fifty-eight (54%) EMTs reported they were unable to reach the call center physician, despite attempts, one or more times each week. Of those 58 EMTs, nineteen (32.8%) reported they were unable to reach the call center physician because the call was dropped, while four (6.8%) reported no one was available to answer the call. EMTs most commonly selected ‘other’ (n = 35; 60.3%) as the reason the phone call connection failed, but did not provide further explanation. . Finally, in evaluating barriers to completion of medical direction, thirty-eight (35%) EMTs reported that at least once a month they were unable to finish the call with the physician, and receive final recommendations, once initiated due to the patient requiring assistance. | Best practices, which may include the use of hands-free technology, need to be established. In India, further work is needed to improve phone communication as a means of supporting more comprehensive prehospital care. | The self-reported survey data is subject to recall bias. Almost 27% of EMTs reported not calling for medical direction due to patient needs at least 2-3 times per week. However, according to call center data, respondent EMTs averaged 19.5 patients/week (SD 9.6) with 19 calls to the physician/week (SD 7.4) during the month of February. Notably, there are wide standard deviations in this data, which may account for the distribution of responses seen in this study. Furthermore, sampling bias may be present since the population studied was a convenience sample of EMTs working on a single day. Similarly, undercoverage bias is likely since the survey was conducted during the day and did not include EMTs working at night who may have unique challenges. |
| Ganapathy et al (94) | 2016 | Quantitative (cohort study) | Himachal Pradesh | - | - | This included providing access to quality multispecialty health services virtually, through the existing Government Health System, reducing cost and travel, providing 24/7 emergency services, providing government doctors in isolated rural areas access to specialist health information, services, and support, and providing authenticated validated health information to the community promoting health literacy, thus encouraging preventive and promotive healthcare seeking behavior. | An experienced ER specialist from Apollo Main Hospital Chennai telementored the CMO/duty nurse at Kaza and Keylong to provide initial medical support to stabilize patients. One hundred seventy-one out of 2,213 teleconsults were emergencies. Emergency medicine, medical disposables, hardware/software, emergency mobile cart, and videoconferencing systems for direct real-time interactions between the casualty at HP and ER specialists at Chennai were made available. The total cost of the telehealth project for 15 months, with its major societal impact, was $350,000 (USD). A detailed survey of 105 users of the telehealth services indicated that in addition to saving considerable effort, time, physical discomfort, and emotional stress, $13,040 (USD) would have been spent in travel alone for obtaining perhaps suboptimum healthcare. provided teleconsults so far, the community would already have saved $217,000 (USD). The nontangible benefits are literally priceless and cannot be quantified, including the happiness that a caring government has facilitated quality accessible healthcare with no cost to the end user. In a first of a kind, pilot initiative telecervical cancer screening has commenced . A nurse trained semivirtually performs a cervical speculum examination and takes a smear after coating with acetic acid. These images are captured and sent to a senior consultant gynecologist in Chennai who reviews the images and counsels the patient. Smears are couriered to Chennai where the slides are evaluated by a trained cytopathologist. Out of the first six patients for whom this was done, clinically four were normal and two were diagnosed to have cervical erosion. Telelaboratory - The local staff were virtually trained and soon became adept at using the kit. In 7 weeks, random blood sugar was checked in 434, Hb in 325, PCV in 307 had TSH, Troponin I in 8, besides urine routine in 28, TSH 6, and lipid profile in 2, resulting in better clinical management. Objective assessment of user satisfaction is under way. An interim interview of 659 patients revealed that 75% were delighted and only 9% dissatisfied with the telehealth services provided. | Mobilizing equipment within time constraints, facing melting snow and landslides was the first obstacle. Bandwidth issues (technical and commercial) were mitigated by seeking redress at the highest level. Dedicated, customized Very Small Aperture Terminals were provided by BSNL7 —(India’s largest government network provider). Major change management issues were faced with the local staff, who initially perceived telehealth as a threat. Limited infrastructure, multiple dialects, poor health-seeking behavior, and total unawareness of telehealth compounded the challenges. Technology provided virtual specialists on a screen—but making available drugs prescribed and tests requested was extremely difficult. Making sophisticated urban teleconsultants constantly use generics for limited medicines available and avoiding multiple and sophisticated investigations were also difficult. Trained coordinators made house-to-house visits to create awareness within the community, | Evaluation confirms that delivering remote healthcare in inhospitable terrains in a PPP mode is effective. | - |
| Zayapragassarazan et al (72) | 2016 | Quantitative (Cross-sectional study) | Puducherry | Healthcare Professionals | 120 | The main objective of this study was to assess the awareness, knowledge, attitude and skills of telemedicine among the health professionals working in the teaching hospitals of Puducherry Region of India. | The highest and lowest mean value for awareness about telemedicine was recorded among the faculty members teaching postgraduate courses (Mean 20.35 and SD 3.76) and among faculty members teaching undergraduate courses (Mean 16.84 and SD 4.29) respectively. The highest and lowest mean value for knowledge of telemedicine was recorded among the male respondents (Mean 10.43 and SD 2.78) and among the respondents whose age group was between 51-60 years (Mean 8.2 and SD 3.11) respectively. The highest and lowest mean value for attitude towards telemedicine was recorded among the respondents whose age group is between 51-60 years (Mean 35.61 and SD 3.12) and among the para-clinical respondents (Mean 33.21 and SD 3.76) respectively. The highest and lowest mean value for skills of telemedicine was recorded among the para-clinical respondents (Mean 25.14 and SD 3.58) and among the respondents whose age group is between 41-50 years (Mean 22.1 and SD 4.56) respectively. The awareness level shows that 12% of the respondents have low level of awareness, 25% have average level of awareness and 63% have high level of awareness. The knowledge level shows that 24% of the respondents have low level or below average level of knowledge of telemedicine, 35% have average or moderate level of knowledge and 41% have high or above average level of knowledge of telemedicine. With respect to the attitude level 30% of the respondents have low or below moderate level of attitude towards telemedicine, 31% possess moderate level of attitude and 39% possess high attitude level towards telemedicine. With respect to the skill of telemedicine 56% don’t have adequate skills of telemedicine, 25% possess moderate skills of telemedicine and only 19% have adequate skills of telemedicine. Only 60% had expressed interest in adopting this new technology for their future career. Respondents who were less than 45 years of age had shown more interest and 91% of the respondents had expressed interest in undergoing training programmes and acquire hands-on experience on telemedicine. | Content analysis of responses to open-ended questions and direct observations made by the researchers revealed lack of organizing skills, technical skills including computing and management skills in organizing telemedicine sessions, management of telemedicine unit and handling telemedicine equipments was the common problems faced by the telemedicine users. Further it was also noted that there is no adequate financial and infrastructural support from the administration for using telemedicine. One of the remarkable comments among the telemedicine users is their concerns about ethical issues with respect to handling of patients, patient data and privacy. | It is the need of the hour for better dissemination of information about the state of the art of research and development in telemedicine and to intensify training workshops for health professionals and improve telemedicine facilities to reach the unreached. | This study was administered among the faculty members working in selected medical and para medical colleges in the Puducherry region and the results cannot be attributed to the whole health professional population. The study does not take into consideration the cultural issues towards telemedicine for discussing the results. The study was administered to the faculty whose contact details are available in the researchers department and hence the results cannot be attributed to the whole faculty community of the concerned colleges. |
| Patro et al (79) | 2015 | Quantitative (Cross-sectional) |  |  | 206 patients screened by 2 doctors | The study was aimed at estimating the diagnostic agreement of common dermatological conditions between a Priamry Care Provider (PCP) and a teledermatologist. | The overall degree of agreement between the diagnoses made by a PCP and the final diagnoses by the dermatologist was 56%. Good agreement (κ > 0.7) was seen in the diagnosis of pyoderma (0.76), moderate agreement (<0.4< κ <0.7) was found in scabies (0.56) and dermatophyte infection (0.49). However, κ agreement was poor (κ < 0.4) in case of eczema (0.37) and psoriasis (0.27). PCPs correctly diagnosed only 2 of the 14 cases of psoriasis but 7 out of 8 cases of impetigo and 16 out 18 cases of scabies were diagnosed correctly. However, 11 cases of eczema had been misclassified as scabies.All five cases of papular urticaria had been misidentified as scabies whereas PMLE had been misdiagnosed as dermatophyte infection in 6 out of 7 instances. The PCP could correctly diagnose pityriasis versicolor in 6 out of 7 cases. | Since the diagnosis by the specialist dermatologist was based on digital images of patients seen by the PCP, taking proper images for better visualization becomes extremely important. | Teledermatology can supplement specialist dermatology service in remote areas. | The major limitation in this study stems from the fact that the diagnosis made by the dermatologist is based on digital images and patient’s history and not actually examining the patient which could have been the ideal situation. |
| Raman et al (88) | 2014 | Quantitative (Cross-sectional) |  |  | 3522 people with diabetes mellitus underwent ophthalmologist-based diabetic retinopathy screening and 4456 people with diabetes underwent ophthalmologist-led (telescreening) diabetic retinopathy screening. | To determine the accuracy of telescreening compared to the traditional camp-based screenings. To compare the prevalence of diabetic retinopathy between an ophthalmologist-based and an ophthalmologist-led model on two different samples of people self-reporting with diabetes in rural South India. | A total of 519 people (14.7%) were diagnosed to have diabetic retinopathy in the ophthalmologist-based model, and 853 people (19.1%) in the ophthalmologist-led model (p<0.0001). More sight-threatening retinopathies were found in the ophthalmologist-led model than in the ophthalmologist-based model (6.3% vs 5%). Mode of transmission in our study was satellite-based. Satellite transmission of data eliminates the need for having local internet connectivity and the need for infrastructure at the village site. | - | Because it obviates the need for travel by an ophthalmologist, telescreening is a good method for diabetic retinopathy screening in rural areas of India. | - |
| Meher et al (90) | 2013 | Mixed-Methods |  |  | Of 143 respondents, 61 were Interns and Residents (42.6%) and the rest 82 (57.3%) were faculty. | To understand legal issues pertaining to telemedicine | 39.9% of the total respondents were somewhat concerned about the legal issues associated with telemedicine, followed by 21.7% who were strongly concerned about the legal issues. It is observed that 29.4% of the respondents somewhat agreed that they may get sued due to the malpractice through telemedicine , followed by 25.2% who have no opinion on this, among total 13.3% somewhat disagreeing to it, while 11.9% strongly agreeing. 38.5% respondents have no opinion on legal issues, followed by 28% of the respondents who were not comfortable in giving teleconsultation due to legal issues, while 15.4% somewhat disagree, and 12.6% strongly agree that they are not comfortable in teleconsultation. 39.2% strongly agree that policies should be made on legal issues. | - | This would help the doctors to be careful during the consultation process and initiate telemedicine consultation accordingly. Legal back up (to save mistakes). All legal issues associated with teleconsultation should be classified & policies to be made. | - |
| Keeppanasserril et al (81) | 2011 | Quantitative (Cohort) |  |  | 4 interns and 60 patients | To find out whether newly graduated dentists, under remote guidance from specialists, can fabricate over-dentures that are functional and improve the oral health related quality of life. | FAD scores of the two groups of the dentures fabricated at both the sites didn’t exhibit statistically relevant difference. Comparison between the OHIP -EDENT scores before and 3 months after treatment revealed that subjects in both groups exhibited significant improvement in all the domains. Overall, 11 out of 19 subdomains showed improvement. No statistically significant difference was observed between the FAD scores of the two groups of patients. Both groups also showed significant improvements in almost all domains of OHRQOL as measured by OHIP -EDENT. | Training of the general dentists, hardware and software problems, poor network, and patient compliance were few of the problems encountered during the study. A high degree of unreliability in connectivity and the need for on-site information-technology support often presented hurdles and led to several appointments at site 1 being extended or even rescheduled. Transportation issues leading to missed appointments, poor oral hygiene maintenance, and the not-so-rare lack of family support for the long treatment sequence also presented problems. | This strategy has the potential to improve access to care and elevate the level of dentistry available to rural population when referral to specialists in not feasible.. | Limitations is the choice of study design. Number of subjects in the study had to be limited due to financial, technical and logical constraints. No attempt was made to match the two groups of patients with each other, which reduces external validity of the results. Absence of an economic evaluation is another drawback of the study from policy and sustainability point of view. |
