## Supplementary material for "Barriers and Facilitators for the Use of Telehealth by Healthcare Providers (HCP) in India - A Scoping Review": S3 Table

S3 Table: Study characteristics for studies on tele-education as mentioned by the authors.

| Study identification* | Year of Publication | Study design and setting | States Studied | Population studied | Sample size | Primary objectives | Positive findings/Facilitators | Barriers | Implications for healthcare delivery/ universal health coverage | Limitations of the study |
| --- | --- | --- | --- | --- | --- | --- | --- | --- | --- | --- |
| Priya et al (106) | 2021 | Quantitative (Cohort study) | Not available | Nursing and allied health professionals | 347 | To develop and validate the effectiveness of a web-based learning module on oral health promotion among nursing and allied health professionals. | The web-based learning module effectively improved the knowledge of the participants. | - | Web based training could be used to remotely train human resources for health. | Incomplete data of the participants and evaluation of only the short-term effectiveness of the web-based programme on oral health promotion were the mentioned limitations of the study. |
| Philip et al (97) | 2021 | Quantitative (Cohort study) | Punjab | Doctors | 114 | To train for mental health in PHC using tele services. | Through the training, 37% improvements in knowledge scores of the participants on case-based scenarios were noted. Improvements in attitudes and practices towards MHCs were also noted. These demonstrate high participant engagement and satisfaction. | - | Capacity building through tele training. | - |
| Gautam et al (108) | 2021 | Quantitative (pre-post study) | Uttar Pradesh | Medical Officers(Doctors), Community Health Officers(Nurses) | 896 | To assess the impact of online training and to overcome the limitations of e‑learning. | Online training is beneficial for many candidates as improvement in average marks was seen. The Medical Officers and Community Health Officers participated enthusiastically in the training programme. | Few candidates were unable to keep up with the pace of the training module and it was difficult to keep track of all the trainees. | E-training could be used to effectively train human resources for health. | 222 Community Health Officers did not give the post-test. |
| Orsolini et al (121) | 2021 | Quantitative (Cross-sectional) | - | Medical students, psychiatry trainees and early career psychiatrists | 60 | To evaluate the level of training, knowledge, experience, and perception regarding the topic of digital psychiatry in a sample constituted by medical students, psychiatry trainees, and early career psychiatrists. | Most participants believe that digital psychiatry may be indicated mainly for follow-up visits of already known and pharmacologically stable patients and it is effective as face-to-face interventions. | Lack of training on new digital tools and digital interventions in psychiatry | Digital intevention offers a costeffective solution in those circumstances in which the current mental health services and infrastructure are not able to properly accommodate the patients' needs | Findings not generalizable to the entire country because of a small and regional sample size. |
| Rao et al (117) | 2021 | Quantitative (Cross-sectional) | - | Health care providers | 2706 | To analyze the planning process and implementation of fast-track online/in-person training during the Coronavirus disease (COVID-19) pandemic, and evaluate its effectiveness in building a rapid, skilled, and massive workforce | Good bandwidth connectivity was essential for uninterrupted and good quality video and audio, which was ensured by information and technology (IT). The availability of appropriate infrastructures such as training spaces and audio-visual systems to run multiple parallel sessions, as well as manpower resources to train the participants, facilitated the rapid generation of a workforce during the lockdown period. | - | A training action plan for disease outbreaks would be a useful resource to tackle such medical emergencies affecting substantial populations in future. | - |
| Shiva et al (98) | 2021 | Quantitative (Cross-sectional) | - | Health care providers | 41 | To assess the effectiveness of virtual courses. | A total of 90.2 % fulfilled attendance criterion. The average total feedback score from all 8 sessions was 9.54/10. | Technical problems faced due to internet connectivity issues posed a challenge. | Virtual certificate course can be provided to other professionals globally and may also serve as an example for other centres to develop similar courses. | - |
| Hariprasad et al (118) | 2018 | Quantitative (pre-post test study) | Karnataka | Health care providers | 27 | To present a low-cost training model that is highly suitable for resource-deficient settings, such as those found in India, through Extension for Community Health Outcome (ECHO), a knowledge-sharing tool, to enable high-quality training of HCPs. | At the start of the training program, a baseline evaluation of knowledge was conducted using the predesigned questionnaire. The train - ing started with sharing knowledge about cancer screening (why, when, and how) and the pur - pose and objectives of cancer screening in the primary health care setting. These topics were discussed along with hands-on demonstration. On the final day, all of the topics discussed were reiterated. The greatest knowledge increase was seen in sterilization methods (220%), followed by cervical cancer information (194.7%) and breast cancer information (107.7%). We compared the change in the knowledge level between the post-training questionnaire and the questionnaire given after 6 months of intervention (Table 3). In this comparison, the greatest increase in knowledge was seen in the areas of breast cancer (8.9%), cervical cancer (6.7%), oral cancer (4.2%), and sterilization methods (4.2%). ECHO is a low-cost tool that can be easily accessed on a desktop or laptop computer, tablet, or smartphone with broadband or wireless Internet connectivity, and most PHCs are equipped with a desktop computer. ECHO can be set up by adding a Web camera, microphone, and speakers at nominal cost. Many HCPs also carry smartphones with them, through which they will be able to download the software required for ECHO, which is free of cost. These training sessions were conducted over ≤ 1 hour when the HCPs came to the PHC to attend their weekly meeting. Most importantly, ECHO eliminates travel time of the HCPs—time that can be used in screening the population. It also helps in reiteration of the study material and to update the cohort of any recent research development, which HCPs can then apply in the field to enhance the quality of cancer screening without having to wait for the next in-person training session. Using a widely spoken language, in this case Kannada, as the medium for teaching and conducting the training plays a huge role in the HCPs’ understanding of the information being taught. | In-person training is essential before HCPs implement a population-based cancer screening program because their knowledge about cancer and screening tests is minimal. | ECHO is an affordable and effective model to train HCPs in cancer screening in a resource-constrained setting. | Survey response rates were low. |
| Bhattarai et al (123) | 2021 | Quantitative (Cross-sectional) | - | Physicians and surgeons | 130 | To study the perceptions, practice and preferences of medical residents and professionals about webinar‑based teaching. | Webinar-based teaching was suggested to be continued in various ways after COVID‑19 period by 87% participants. Among them, 65.6% were of opinion that webinar should be continued after pandemic in equal proportion or as an adjuvant to conventional class. Only 2% opined that it should be never used after pandemic. Four out of five participants (80.8%) said that webinar recording is useful while 71.5% said that pre‑ and post‑webinar questionnaire should be practiced. Majority (91%) participants found the webinars were useful. | Webinar hindered works in one fourth doctors. Ongoing webinar were found to be less interactive by 33% while 33.8% labelled most ongoing webinars as ‘waste of time’. Sixteen (12.3%) participants argued that webinars have increased stress in residents and professionals whereas 44.6% opined that it helpful and should be done more frequently. | Teaching through virtual mode (e.g. webinar) is a valuable tool for medical education especially during the need of social distancing. | Findings of the survey were not generalizable due to poor sampling technqiue. |
| Manjunatha et al (109) | 2021 | Quantitative (retrospective study) | Karnataka | Primary care physicians | 73 | To establish performance indicators of Tele‑OCT for its effective implementation. | A total of 146 Tele‑OCT sessions were fixed during the study period, but 116 (79.5%) sessions were successfully conducted. Almost 80% of Tele‑OCT sessions are successfully conducted, denoting a higher level of feasibility. The remaining 20.5% of Tele‑OCT sessions were canceled. | Thirty sessions were canceled; the reasons are abrupt deputation of PCDs for urgent duty (n‑19), due to public strike (n‑6), no patients at outpatients (n‑2), and poor internet connectivity especially on rainy days (n‑3). | Tele‑OCT appears to be a path‑breaking training model for PCDs to integrate psychiatric care in their general practice. | Patient profiles of each PCD are specifically not analyzed to understand whether each PCD is trained in all kind of psychiatric disorders. |
| Ibrahim et al (101) | 2021 | Quantitative | Chattisgarh | Rural medical officers and primary care doctors | 501 | To give a brief overview of the Community Mental Healthcare Tele-Mentoring Program (CHaMP) initiative. | This approach empowers the primary care doctors to effectively deliver mental health services to the underserved population along with having support and supervision from specialists. This program involves very less traveling and saves much time for both the trainees and trainers. | - | Such training models to healthcare workers at the grass root level (in comparison to direct consultation to patients/clients) seem to be a more efficacious model for service. | - |
| Doherty et al (122) | 2021 | Quantitative (Cross-sectional) | - | Physicians, nurses, medical officers, pharmacists, project cordinator | 18 | To describe the experiences of Implementing a Teleteaching and Mentorship Program (Project ECHO) | The main factors that facilitated participation include convenient session timing and duration, ease of accessing the sessions online, and reminder e-mails. Participants were very motivated to attend further training, and all stated that they would recommend ECHO PPC to others. | The most significant obstacles of participation included Internet connection problems and busy work schedules. | These adaptations may improve the efficacy of Project ECHO and others using virtual learning programs in resource-limited settings. | - |
| Kumar et al (105) | 2020 | Quantitative (cross-sectional) | - | Dental students | 170 | To assess the impact and acceptance of e‑learning for dental education. | 58.7%, 58.9%, and 64.7% of slow, moderate, and advanced learners said that e‑learning methodology helped very much to improve their knowledge. 63% of slow learners, 71.4% of moderate learners, and 69.1% of advanced learners said that they have noticed an overall improvement in clinical due to e‑learning technique and especially in identifying clinical features, diagnosis, and treatment planning. When e‑learning methodology compared with traditional methods of learning: 34.8% said e‑learning is better and 47.8% said e‑learning is very much useful among slow learners. Most of the students reported that (69% of slow and moderate learners, 60% of advanced learners) e‑learning methodology has overall improvement in theoretical and clinical training. e‑learning methods improved the overall performance of students appearing university examinations by 97.8% in slow learners, 98.2% in moderate learners, and 97.1% among advanced learners. | Initially, the attitude and awareness of students to learn through online education was challenging. Poor education and motivation of all the teachers toward the online education system was also a barrier. | Traditional teaching approach could be shifted to e‑learning methodology. | - |
| Barik et al (112) | 2020 | Quantitative (Cross-sectional) | - |  | 227 | To present the viewpoint of the orthopedic residents to the paradigm shift in clinical care as well as the academic activities to to a form of a web-based learning process and simulation-based training. | Most of the residents found overall learning through web platforms (44.2%) to be easier than before. Although there were diffculties in facing a complete online interface-based viva voce (45.8%), multiple-choice questions (MCQ) or Objective Structured Clinical and practical examination (OSCE/ OSPE) did not pose any problem for most of the residents (47.8% and 45.6% respectively). | The majority of residents felt that participating in an online case presentation (55.3%) and maintaining the attention of the audience (63.8%) during any online presentation were relatively difficult than offline activities. | This study can help the universities as well as program chairs to develop a robust program that can outlive this pandemic. The web-based learning process might prove to be useful and can be incorporated into the resident training program in the long term. | A major limitation of this study is that the fndings are based on the responses of orthopedic residents from only one country, which may not be extrapolated to other countries all over the world. |
| Pahuja et al (102) | 2020 | Quantitative (Cross-sectional) | Uttarakhand | Primary care physicians | 11 | To highlight the effectiveness of an optional opioid training module for a PCD. | In this paper, authors demonstrated the effectiveness of digitally driven CVCs between a telepsychiatrist and remotely practicing untrained PCD in an OST clinic for 11 difficult OUD patients. This denotes that if PCDs are properly trained and guided when required using video technology, they may act as a long-term viable solution to reduce treatment gap, morbidity, and mortality associated with OUDs. | - | With this kind of telemedicine-based guidance from a telepsychiatrist, PCDs can be trained to manage OST clinic in remote PHCs, especially in the areas with high-prevalence of OUDs. | - |
| Lakshminarayanan et al (104) | 2020 | Qualitative | Karnataka, Maharashtra, Chattisgarh, Bihar | Field workers | - | To describe the experience of remote training of persons with no previous knowledge or experience of mental health to recognize mental health problems and deliver psychosocial services at the grassroot level and discuss the impact and acceptability of the training. | The participants had little knowledge relating to mental health and mental illness. The average score of the participants in the pre-training assessment was close to 20 %, which, in the post-training assessment went up to 75 %. Experience of the participants was reported to be “very satisfactory”. They emphasized that, despite their initial apprehension and disappointment at technical glitches, they were happy that they were able to communicate and learn from their own offices. They confirmed that they were more confident about the subject after the training. The results of the training and the overall experience encourage the need to use digital platforms to address gaps in mental health training and service delivery. | There were challenges such as unfamiliarity, poor technological competencies, and issues of connectivity. | The authors of this paper do not advocate digital training as an alternative to face-to-face hands-on training. Based on the trial, the authors recommend it as a viable alternative in situations with a limited budget and other constraints | Costs involved in digital training vary widely. The current training program used simple mobile phones, tablets, and basic computers. |
| Babu et al (113) | 2020 | Qualitative | - | Health care providers | 471 | To provide a systematic and structured curriculum that elaborates on the methods, various courses conducted on the ECHO platform, challenges faced during the program, and strategies used to overcome them. | The training will be conducted in their regional language with the experts who speak and understand their local language, to create awareness in cancer screening and sensitize the CHW community in facts and myths of cancer screening. | In the event where State authorities were engaged in recruiting the medical officers, it was observed that the adherence to the training program was poor. This could be attributed to a lack of motivation, lack of awareness about the program, and unwillingness of the participants. Technical issues ranged from internet connectivity or inability to install or use the software used for the course, while academic reasons comprised of missing a session and not knowing how to get back on track. | Training offered using a hybrid model as mentioned above might be the most effective and help a country like India to achieve WHO’s goal of eliminating cervical cancer by the year 2030 and reduce the burden of preventable cancers. | - |
| Dhanasekaran et al (100) | 2019 | Quantitative (pre post-test study) | - | Health care providers | 32 | To disseminate the experience on the effectiveness of this hybrid model in training health care providers in cancer prevention. | The online participants had significantly (60%) more knowledge in cervical cancer screening compared with their counterparts. The intervention group participants had performed 26% better than their counterparts. Good feedback was received from most participants - useful, informative, learning experience. | Internet connectivity was a critical factor in deciding the participation in some remote places. Only a limited number of participants from the online group attended the hands-on training due to the cost involved in traveling and accommodation. | This intervention can be a potent tool for the government for efficient training of HCPs in cancer screening. | - |
| Nethan et al (116) | 2019 | Quantitative (pre post-test study) | Pan India | Health care providers | 48 | To assess training effectiveness using ECHO model | The program evaluation results illustrated a significant knowledge gain among the participants regarding oral cancer screening and tobacco cessation. Both the trainees and also the trainers could attend the meetings from the comfort of their respective locations. This training module also proved apt for HCPs, especially in the urban settings, having busy schedules, as the sessions were conducted only once a week, lasting for a duration of about 1 h each. | post evaluation was filled by lesser number of participants possibly because of their busyschedule or apprehension of being assessed | this provides a conveniant,cost effective,large scale,best practise tele mentoring tool | The limitation of this study is the lack of longitudinal follow-up of the spokes to know their adaptation of the learnings into their practice.post evaluation nos are less. |
| Sagi et al (111) | 2018 | Quantitative (pre-post test design) | Bihar | Primary care physicians | 38 | To develope an innovative telementoring model and to look at its feasibility as well as acceptability among remote PCPs on drug addiction management. | All the PCPs were familiar with its use for social networking. Hence, it was used for sharing of nonpatient-related information, discussion, reminders, and sometimes didactic presentations. The tele-ECHO clinics were conducted in Hindi, the local language of the participants. Among 38 PCPs, a cumulative of 89.47% (n = 34) had completed three e-learning assignments. On post hoc analysis, the knowledge scores were significantly more during 1-month (3.00 ± 0.86, P < 0.001) and 3-month (3.16 ± 0.90, P < 0.001) assessments compared to the baseline (1.77 ± 1.02). About 32.25% (n = 10) reported improved confidence level and 54.83% (n = 17) reported the same confidence level in managing a case of SUD. About 22.58% (n = 7) reported improved satisfaction level from baseline, 51.61% (n = 16) reported the same satisfaction levels above 5, and 16.12% (n = 5) reported the same satisfaction level below 5 about the management of patients with SUDs. | On an average there were 3-4 breakages in the internet connection per participant in each tele-ECHO clinic. About 25.92% (n = 7) of participants required >5 attempts to complete the assignments in part because of language barriers (English) and technological issues. | Telementoring model in e-health care has tremendous future potential using technology as a part of learning. Preliminary evidence suggests that this model is a feasible model to enhance the training and mentoring of existent human resources to address major public health problems such as drug addiction. | This was a small unicentric study with limited participants from a single state. Only a single-focused area of de-addiction was undertaken and therefore the findings are not generalizable. |
| Mehrotra et al (107) | 2018 | Quantitative (Cross-sectional) | Chattisgarh | Mental health counsellors | 12 | To ascertain the effectiveness of Project ECHO, a Hub and Spokes tele-mentoring model to bridge the urban-rural divide in mental health and addiction care in the context of a developing country like India. | Over the 12 fortnight tele-ECHO clinics, 6 participants had an attendance of over 80% and all participants maintained an attendance of over 60%. There were no dropouts. There was a difference in the mean scores from the pre-test (M = 11.25 ± 2.98) to the final post-test scores (M = 12.91 ± 1.73)(p < 0-01). All participants passed all assessments. There was an statistically significant increase in self-confidence. There was a high rate of participant satisfaction. | - | The results indicate promise of the NIMHANS ECHO tele-mentoring model as one with potential for capacity-building in mental health and addiction for remote and rural areas by leveraging technology. | The small numbers limit the generalisation of the findings. The authors did not monitor the longterm effectiveness of the training. |
| Balasubramaniam et al (99) | 2017 | Quantitative (pre-post intervention study) | Bihar | Auxillary nurse midwives | 85 participants in the control arm and 51 participants in the study arm. | To assess the efficacy of a training model blended with virtual training as compared to conventional classroom teaching in auxillary nurse meidwives in Bihar. | The students exposed to blended learning scored 32.57 points (p = <0.001) more than their counterparts, who received only conventional teaching. In the post-intervention cohort, 55% students (N = 28) passed as compared to none in the pre-intervention cohort. Mean scores for all six practices also increased significantly from pre-to post-intervention assessments. Competency levels differed significantly (p < 0.001) between the two cohorts: none of the 85 students in the pre-intervention cohort achieved the cut-off score of 75%, compared with 55% (28 of 51 students) of the post-intervention cohort. | - | The approach is especially useful for educating students who attend training institutions that have limited faculty and infrastructure. Based on the study findings, we hypothesize that in the future, the blended learning model will be able to prepare more competent and confident auxiliary nurse midwives than conventional training. | Only objective structured knowledge was assessed and not real life clinical competency. The authors assumed that the students of the two cohorts assessed before and after the intervention were different, because it was impractical to assess the same students before and after the implementation of the intervention. The authors also assumed that students in consecutive academic classes at the same institution would have similar in personal characteristics and would be exposed to the same standard of conventional classroom learning. |
| Bansal et al (124) | 2015 | Quantitative (Cross-sectional) | NA | Physicians | 17 | To test the feasibility of using a novel point of care echocardiography training program for improving physicians’ imaging skills during preanesthetic cardiac evaluations performed in a community camp organized for treating cataract blindness. | Of the 968 scans performed, 861 (88.9%) were graded to have excellent, good, or fair image quality and another 46 (4.8%) as technically difficult but adequate. The remaining 61 (6.3%) had poor or limited image quality, but only 12 (1.2%) were nondiagnostic, precluding any meaningful interpretation. Importantly, there was no difference in the overall image quality across different skill levels. A comparison of onsite and remote interpretations revealed that the remote physician could recognize major echocardiographic abnormalities with 58.7% sensitivity and 97.0% specificity (overall k = 0.62, P < .001), though severity was underestimated by one grade in 11 studies (11.2%) and overestimated in another 11 (2.0%). There were no significant differences in the accuracy of findings reported by onsite versus remotely trained physicians. The majority of the remote physicians (12 of 17 [70.6%]) expressed complete or at least reasonable satisfaction with their overall experience during the entire activity For both groups, the training resulted in significant improvements in selfperceived competence in all components of POC echocardiography. | not mentioned | Web-based integration of remote, expert interpretation of stored images, allows the delivery of echocardiographic expertise to remote communities, which could be of great help in optimizing cardiovascular health outcomes in these communities. | First, for logistic reasons, we could not use elaborate, objective tools for quantifying the impact of training on different components of competence among PhyS in using POC echocardiography and to determine the influence of baseline skill and mode of training on the imaging outcomes. Second, although the initial intense training was only for 6 hours, subsequent echocardiographic scanning during the camp provided continued learning opportunity to the PhyS and must have contributed to further improvements in their imaging abilities. Third, in the community camp, the PhyS performed echocardiographic scanning under supervision by ExpS, which precluded Figure 5 (A) Overall experience of PhyS with the remote, Web-based training. (B) Overall impact of training on the self-perceived competence of the physicians in different components of POC echocardiography. All scores were derived using a five-point modified Likert-type scale, as described in the text. Journal of the American Society of Echocardiography Volume 28 Number 1 Bansal et al 83 assessment of their ability to independently perform POC echocardiography. |
| Mahadevan et al (119) | 2012 | Quantitative (Cross-sectional) | Karnataka and West Bengol | Post graduate trainees in critical care, anesthesia and superspeciality candidates in cardiac anaesthesia | 1000 | To examine the feasibility, advantages, and disadvantages of this method of e-learning sessions. | The participants expressed satisfaction over the teaching, and the attendance was 84.4%; of those, about 48.4% of participants expressed good and 37% excellent impressions of the program. | The disadvantages include need of commitment from both sides, costly multipoint equipment, technical difficulties, need of planning and scheduling, and need for technical support. | Telemedicine offers solutions for emergency medical assistance, long-distance consultation, administration and logistics, supervision and quality assurance, and education and training for healthcare professionals and providers. | - |
| Chang et al (110) | 2013 | Quantitative (pre post-test study design) | Maharashtra | Health care providers- physicians ,residents or students | 115 | To evaluate a distance learning course on HIV management for clinical care providers in India. | Most participants indicated that they had access to a computer (81%) or the internet (71%) “all the time” or “daily.” Participant scores rose significantly from a mean proportion answering questions correctly of 78.4% on the pretest to 87.5% on the posttest. After completing the course, responding participants were more likely to be confident in several aspects of patient management including starting an initial antiretroviral (ARV) regimen, understanding ARV toxicities, encouraging patient adherence, diagnosing immune reconstitution syndrome, and monitoring patients on ARV medications (all Ps ≤ .05). All participants strongly agreed or agreed that they would recommend this course to others, and most of them (96%) strongly agreed or agreed that they would take a course in this format again. | - | - | The study has a small sample of participants and reports on a single distance learning experience. Additionally, the response rate was not complete, which may have introduced biases. |
| Thukral et al (115) | 2012 | Quantitative (pre post-test study) |  | Nurses | 98 | To evaluate the efficacy of internet-based distance learning in conjunction with local hands-on skill enhancement in improving knowledge and skills of essential newborn care among in-service nursing health professionals. | There was significant increase in knowledge. There was a significant improvement in the post-test OSCE scores of all participant groups except one. In addition, it increased their confidence for day to day skill in work and resolved their doubts to a reasonable extent. They were satisfied with the tutoring skills of their online tutor. All of the respondents said that they would like to participate again in a similar venture in the near future, and would also recommend the programme to their colleagues. | Twenty-two (22%) participants were not computer literate and did not know the use of internet before the enrolment in the course. | Online training and teaching in essential newborn care is feasible and acceptable for in-service nursing professionals and serves as a useful tool for professional development of their practical skills and knowledge. | The limitations of the study include the limited utility of this programme in areas with no facility of information technology. Slow streaming for video viewing was identified as a barrier. |
| Ramanathan et al (114) | 2011 | Mixed-Methods (quantitative pre- and post-course knowledge assessments, and qualitative focus groups and in-depth interviews) |  | Physicians and nurses. | 42 | To examine the effectiveness of a pilot internet-based continuing medical education course in increasing knowledge of pediatric HIV diagnosis and treatment among providers in Pune. The study also explored perceived factors limiting the effectiveness of the pilot course. | Mean post-test scores for the global knowledge assessment and two of the five course modules increased significantly from pre-test scores. The weekly videoconference sessions with the course faculty were well received: attendance exceeded 90%, with 81% of participants reporting that the sessions were helpful to their learning. Overall, 77% of the participants reported the lectures to be of high quality and 88% felt that the material improved their ability to treat HIV. | Participants reported that issues related to Internet access limited them from taking full advantage of the audio and video components of the course. One-fifth of the participants had access to internet only a few times a week. Although all participants were proficient in English, they stated that noisy audio connections, differences in accents and the relatively fast rate of speech of the Johns Hopkins faculty limited their comprehension at times. | This course resulted in a modest increase in pediatric HIV knowledge among Pune healthcare providers. Identification of perceived factors limiting the effectiveness of the course will provide guidance for improving future Internet-based courses. | A small sample size and purposie sample potentially resulting in a selection bias were identified as limitations. An additional limitation of this study is that the focus groups and interviews were led by investigators who, while not involved directly with course development and participant evaluation, were nonetheless affiliated with the course. As a result, participants may have been reluctant to provide criticism. |
| Jain et al (120) | 2010 | Quantitative (randomized controlled trial) |  |  | 48 | To compare gain in knowledge and skills of neonatal resuscitation using tele-education instruction vs conventional classroom teaching. | Pre-training mean knowledge scores were higher in intervention group. However, skills scores were comparable in the two groups . Training resulted in similar gain in knowledge scores and skills scores in both the groups. Trainees’ satisfaction with content, delivery, personal gain and teachers’ ability was comparable in the two groups. Instructors’ satisfaction with teaching experience in the two groups was comparable. | The two groups differed only slightly on the issues of ‘active participation’ and ‘delivery of subject’, both being higher in the conventional teaching group. Minor technical difficulties were encountered during telemedicine sessions. There were three episodes of interruptions due to signal failure. | Although classroom method remains an ideal choice for capacity development, teaching and learning by teleeducation offers a rapid, pragmatic and reasonably effective method of large-scale training of nurses in neonatal resuscitation. | - |
| Mahapatra et al (103) | 2009 | Review |  |  | - | In this study, the authors tried to bring out critical issues in successful implementation of distance medical education using telemedicine technology based on the analysis of activities carried out at SGPGIMS over the past decade and reviewed nationwide efforts in this direction including the policy initiative of government based on the recommendation of the National Knowledge Commission (NKC). | Problems in management of difficult and complicated cases faced by the doctors in Orissa state medical colleges were clarified and guidelines for management were provided. Interesting and illustrative cases seen at SGPGIMS were shown to the postgraduate students and doctors of Orissa, for their knowledge update. Free satellite bandwidth was provided by ISRO for telemedicine projects under the government policy of societal benefit of indigenous space technology. | Though the routine tele-educational sessions with available satellite bandwidth of 300–384 Kbps were adequate, there was a constraint of getting higher bandwidth for carrying out advanced teleeducational applications such as live surgery exchange. | The future holds the key for the development of a countrywide medical college network grid using high bandwidth fiber backbone which, once in place, will facilitate seamless knowledge exchange among the stakeholders. | - |

* manuscript citation number given in brackets
