## Supplementary material for "Barriers and Facilitators for the Use of Telehealth by Healthcare Providers (HCP) in India - A Scoping Review": S4 Table

**S4 Table:  HRH Framework**

| **As defined in HRH Action Framework (https://www.capacityproject.org/framework/)** | | | | | |
| --- | --- | --- | --- | --- | --- |
| **HRM Systems** | **Policy** | **Finance** | **Education** | **Partnership** | **Leadership** |
| Personnel systems: workforce planning (including staffing norms), recruitment, hiring and deployment | Professional standards, licensing and accreditation | Setting levels of salaries and allowances | Pre-service education tied to health needs | Mechanisms and processes for multi-stakeholder cooperation (inter-ministerial committees, health worker advisory groups, observatories, donor coordination groups). | Support HRH champions and advocates |
| Work environment and conditions: employee relations, workplace safety, gender equity, job satisfaction and career development | Authorized scopes of practice for health cadres | Budgeting and projections for HRH intervention resource requirements including salaries, allowances, education, incentive packages, etc. | In-service training (e.g., distance and blended, continuing education) | Public-private sector agreements | Capacity for leadership and management at all levels |
| HR information system integration of data sources to ensure timely availability of accurate data required for planning, training, appraising and supporting the workforce | Political, social and financial decisions and choices that impact HRH | Increasing fiscal space and mobilizing financial resources (e.g., government, Global Fund, PEPFAR, donors) | Capacity of training institutions | Community involvement in care, treatment and governance of health services. | Capacity to lead multi-sector and sector-wide collaboration |
| Performance management: performance appraisal, supervision and productivity. | law and rules for civil service and other employers. | Data on HRH expenditures | Training of community health workers and non-formal care providers. |  | Strengthening professional associations to provide leadership amongst their constituencies. |
| The HRH Action Framework website has been developed as an initiative of the Global Health Workforce Alliance (GHWA) and represents a collaborative effort between the U.S. Agency for International Development (USAID) and the World Health Organization (WHO). | | | | | |
| The HRH Action Framework does not represent an official view of the World Health Organization. Similarly, the information provided on this website is not official U.S. Government information and does not represent the views or positions of USAID or the U.S. Government. | | | | | |
